# Evaluation of polygenic scoring methods in five biobanks reveals greater variability between biobanks than between methods and highlights benefits of ensemble learning

**DOI:** 10.1101/2023.11.20.23298215

**Authors:** Remo Monti, Lisa Eick, Georgi Hudjashov, Kristi Läll, Stavroula Kanoni, Brooke N. Wolford, Benjamin Wingfield, Oliver Pain, Sophie Wharrie, Bradley Jermy, Aoife McMahon, Tuomo Hartonen, Henrike Heyne, Nina Mars, Genes & Health Research Team, Kristian Hveem, Michael Inouye, David A. van Heel, Reedik Mägi, Pekka Marttinen, Samuli Ripatti, Andrea Ganna, Christoph Lippert

## Abstract

Methods to estimate polygenic scores (PGS) from genome-wide association studies are increasingly utilized. However, independent method evaluation is lacking, and method comparisons are often limited. Here, we evaluate polygenic scores derived using seven methods in five biobank studies (totaling about 1.2 million participants) across 16 diseases and quantitative traits, building on a reference-standardized framework. We conducted meta-analyses to quantify the effects of method choice, hyperparameter tuning, method ensembling and target biobank on PGS performance. We found that no single method consistently outperformed all others. PGS effect sizes were more variable between biobanks than between methods within biobanks when methods were well-tuned. Differences between methods were largest for the two investigated autoimmune diseases, seropositive rheumatoid arthritis and type 1 diabetes. For most methods, cross-validation was more reliable for tuning hyperparameters than automatic tuning (without the use of target data). For a given target phenotype, elastic net models combining PGS across methods (ensemble PGS) tuned in the UK Biobank provided consistent, high, and cross-biobank transferable performance, increasing PGS effect sizes (β-coefficients) by a median of 5.0% relative to LDpred2 and MegaPRS (the two best performing single methods when tuned with cross-validation). Our interactively browsable online-results (https://methodscomparison.intervenegeneticscores.org/) and open-source workflow prspipe (https://github.com/intervene-EU-H2020/prspipe) provide a rich resource and reference for the analysis of polygenic scoring methods across biobanks.

## Introduction

Polygenic scores (PGS), also referred to as polygenic risk scores (PRS), have become a major application of genome-wide association studies (GWAS). PGS are constructed by scoring individuals based on their genotype, adding up effects of many genetic variants genome-wide. They can improve existing disease risk models that rely on family history and established biomarkers^1–4^, and individuals in the upper tail of the PGS distribution have an elevated disease risk similar to that caused by rare damaging monogenic mutations for some diseases^5^. PGS have received attention in areas ranging from disease prevention to clinical trials, owing to their wide applicability to personalized medicine^6–9^.

Various methods to derive PGS weights from GWAS summary statistics (effect sizes and their correlation structure) have been developed. These methods are of particular interest as they do not rely on access to individual-level data, which is typically restricted. Furthermore, the largest GWAS are meta-analyses, for which direct access to all individual-level source data is not feasible.

The construction of a PGS from summary statistics can be divided into two main stages: A public stage, that relies only on publicly available data and tools, and a private stage that requires access to individual-level target data, i.e., genotypes and phenotypes. The public stage uses variant correlation (linkage disequilibrium; LD) from reference panels that are matched in ancestry to the GWAS sample to adjust the marginal effect size estimates of genetic variants and derive the per-variant PGS weights. These adjustments include frequentist shrinkage^10^, Bayesian approaches^11–15^, or other strategies like thresholding, which depend on one or more hyperparameters (e.g., p-value thresholds, heritability estimates, or shrinkage parameters).

Many methods allow automatically setting suitable parameters without the use of phenotype data (we refer to this generally as automatic tuning). Alternatively, target data can be used to empirically determine hyperparameters based on, for example, cross-validation (CV). The adjusted variant effect sizes (PGS weights) are used in the private stage to score individuals based on their genotypes using a linear additive model, i.e., to calculate their PGS.

PGS method authors usually claim superior performance to other methods. However, comparisons are often limited to a small number of methods, traits, or target datasets. Furthermore, the input summary statistics used in those comparisons may not reflect the properties of (messy) real-world data, especially those from meta-analyses. In practice, other factors also affect performance, e.g., ease of use and documentation.

The INTERVENE consortium^16^ seeks to develop risk scoring methods that integrate PGS with other health-related information. For this reason, we compared summary-statistics-based PGS methods. Building on the GenoPred suite introduced by Pain et al. that implements different PGS methods in a reference standardized framework^17^, we developed prspipe, a snakemake^18^ workflow that runs seven polygenic scoring methods. A full evaluation including hyperparameter tuning with cross-validation was performed in the UK Biobank^19^ (UKBB) and replicated in FinnGen^20^, Estonian Biobank^21^ (EBB), the Trøndelag Health Study^22^ (HUNT) and Genes & Health^23^ (GNH). In total, we meta-analyzed performances for ten binary disease traits and six quantitative traits in two replicated ancestry groups European (EUR) and South Asian (SAS). Replication in multiple biobanks allowed us to estimate how much PGS effect sizes vary within biobanks (between methods) and how this compares to the variation between biobanks.

We publish our workflow, summary data, and PGS weights, allowing others to replicate analyses e.g., for methods comparisons or developing new polygenic scores from summary statistics. The results of this analysis are made available in a browsable online resource at https://methodscomparison.intervenegeneticscores.org/.

## Results

### Prspipe workflow and experimental setup

We created a snakemake workflow prspipe to run different polygenic risk scoring methods based on GWAS summary statistics. Prspipe makes it possible to automate the within-biobank analyses from Pain et. al.^17^ based on the GenoPred suite of scripts (https://github.com/intervene-EU-H2020/GenoPred). Notable differences include an updated set of methods (p-value thresholding and clumping (pT+clump), lassosum^10^, PRScs^11^, LDpred2^13^, DBSLMM^14^, SBayesR^12^ (robust parameterization) and MegaPRS^15^), the use of LD reference panels provided by the methods’ authors, and software managed partially with containers. We used prspipe to derive PGS weights using both methods’ automatic settings (auto), and grids of hyperparameters (MegaPRS, LDpred2, PRScs, lassosum, and pT+clump)^17^. For the baseline method pT+clump, we considered the score with the most stringent p-value threshold (p < 1e-8, i.e., keeping only highly significant variants) as the automatically tuned score.

The workflow defines steps to set up PGS methods, download and process summary statistics from the NHGRI-EBI GWAS Catalog^24^, run PGS methods (i.e., the derivation of PGS weights), target genotype harmonization, ancestry matching based on the 1000 Genomes superpopulations^25^, and target polygenic scoring with plink2^26^. As PGS performance depends on the genetic similarity of the target and GWAS samples^27^, performance evaluation is stratified according to the matched superpopulation. Using CV, elastic net models combining scores from different methods are fit (ensemble PGS), and the best single PGS weights are selected for each method (hyperparameter tuning)^17^.

We applied this workflow to 14 sets of summary statistics from the GWAS Catalog to derive PGS and predict six continuous traits and 10 binary disease traits derived from harmonized ICD-code-based definitions^20^ (Methods, Table 1). Our main analyses focus on the two replicated ancestry-reference-matched superpopulations: EUR and SAS. The number of cases used for performance evaluation across biobanks ranged from 5,384 (T1D) to 81,487 (type 2 diabetes; T2D) for EUR, and 60 (RA, available in GNH only) to 8,696 (T2D) for SAS ancestry matched target data. The total sample size for performance evaluation for continuous traits ranged from 85,973 (urate, available in UKBB only) to 524,056 (height) for EUR target data, and 13,572 (urate) to 43,197 (height) for SAS target data (Table 2).

**Table 1:**
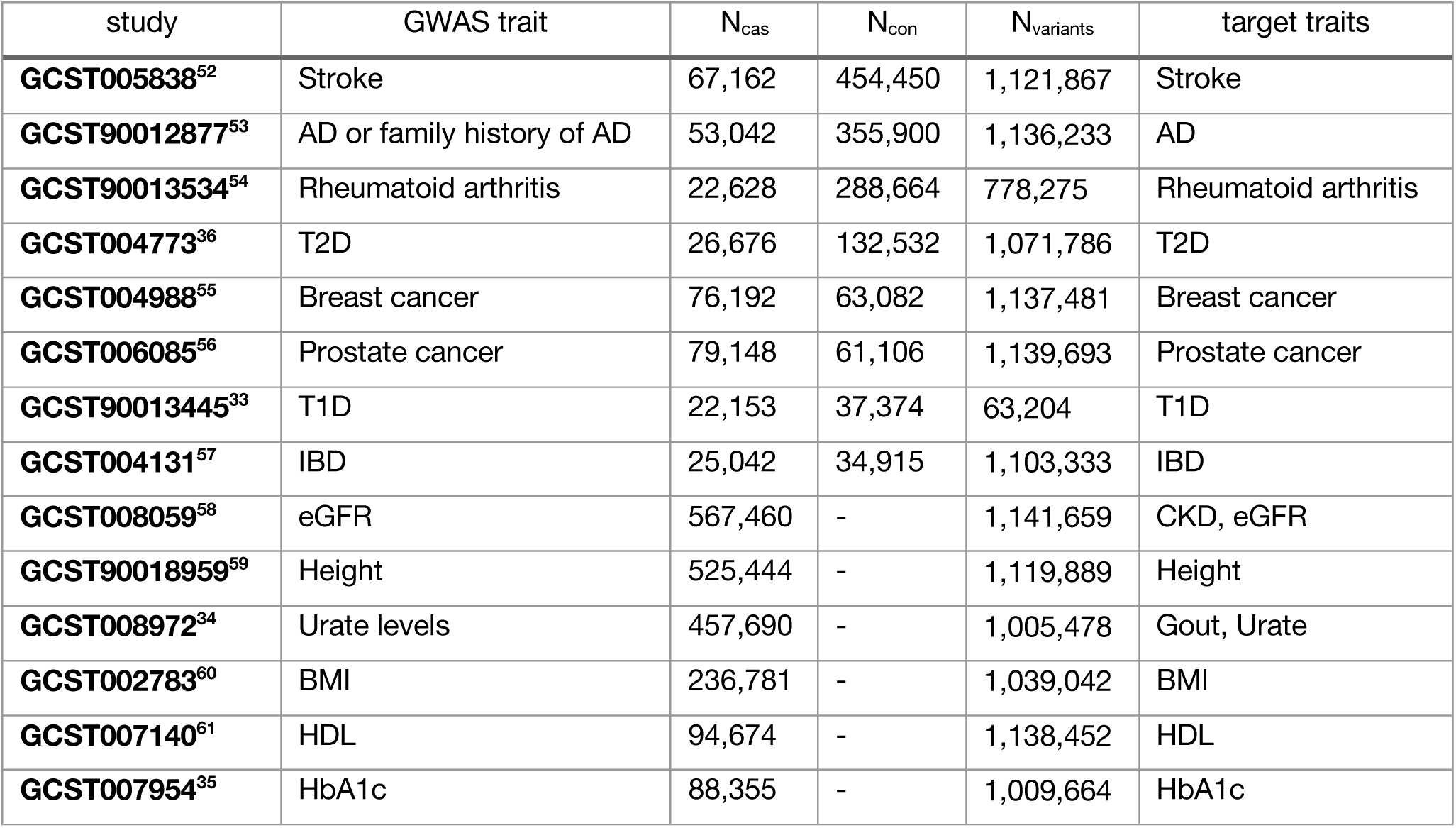
GWAS summary statistics used to derive PGS weights. Entries are ordered by the total sample size and type of trait (binary, continuous). From left to right: GWAS catalog study identifiers (study), the respective reported GWAS traits, number of cases (N_cas_) and controls (N_con_), the number of variants after intersection with HapMap3-1KG and quality control (N_variants_), and the evaluated target traits. Scores constructed from urate and eGFR summary statistics were also evaluated for gout and CKD, respectively. The GWAS for T1D considered only a small panel of variants of which 84% remained after intersection and QC.

**Table 2:** Target sample sizes across traits. For each trait and replicated ancestry group (EUR, SAS) the number of cases (binary disease traits) or sample size are shown, either combined (“total”, excluding UKBB training data) or separated by biobank. For the UKBB-EUR, data were split into train (80%, used to tune hyperparameters and ensemble PGS) and test sets (20%, used for evaluation and meta-analyses). UKBB EUR data were excluded for Alzheimer’s disease and height due to sample overlap and could therefore not be used for tuning (leaving 14 traits for a full evaluation). Dashes “-” indicate the phenotype was unavailable.

|  | EUR<br>total | SAS<br>total | EUR<br>ebb | EUR<br>finngen | EUR<br>hunt | EUR<br>ukbb(test) | EUR<br>ukbb(train) | SAS<br>gnh | SAS<br>ukbb |
| --- | --- | --- | --- | --- | --- | --- | --- | --- | --- |
| **AD** | 15,940 | - | 555 | 13,823 | 1,562 | - | - | - | - |
| **Arthritis** | 13,060 | 60 | 2,384 | 9,332 | 1,139 | 205 | 820 | 60 | - |
| **Breast**<br>**cancer** | 23,610 | 393 | 2,685 | 16,076 | 1,729 | 3,120 | 12,483 | 197 | 196 |
| **CKD** | 19,714 | 1,609 | 4,224 | 9,314 | 2,802 | 3,374 | 13,496 | 1,131 | 478 |
| **Gout** | 22,399 | 488 | 10,646 | 8,759 | 1,318 | 1,676 | 6,704 | 282 | 206 |
| **IBD** | 13,016 | 634 | 2,097 | 7,815 | 1,769 | 1,335 | 5,340 | 466 | 168 |
| **Prostate**<br>**cancer** | 20,492 | 205 | 2,227 | 13,606 | 2,242 | 2,417 | 9,671 | 95 | 110 |
| **Stroke** | 37,920 | 635 | 4,515 | 26,166 | 5,204 | 2,035 | 8,142 | 424 | 211 |
| **T1D** | 5,384 | 443 | 501 | 4,286 | 396 | 201 | 804 | 443 | - |
| **T2D** | 81,487 | 8,696 | 12,344 | 59,345 | 3,861 | 5,937 | 23,748 | 6,630 | 2,066 |
| **BMI** | 346,290 | 42,243 | 189,651 | - | 66,663 | 89,976 | 359,913 | 33,146 | 9,097 |
| **HDL** | 139,248 | 37,693 | 10,642 | - | 49,824 | 78,782 | 315,135 | 29,628 | 8,065 |
| **HbA1c** | 120,242 | 21,696 | - | - | 34,192 | 86,050 | 344,209 | 12,948 | 8,748 |
| **Height** | 524,056 | 43,197 | 190,013 | 267,343 | 66,700 | - | - | 34,089 | 9,108 |
| **Urate** | 85,973 | 13,572 | - | - | - | 85,973 | 343,904 | 4,730 | 8,842 |
| **eGFR** | 152,793 | 38,916 | - | - | 66,759 | 86,034 | 344,140 | 3,061 | 8,855 |

Using 80% of the UKBB EUR target data (training set), we selected the best performing weights for each method and fit ensemble PGS (full workflow). PGS weights were shared with other biobanks, in which we still performed target data harmonization, ancestry matching, and polygenic scoring steps needed for performance evaluation (Figure 1).

**Figure 1:**
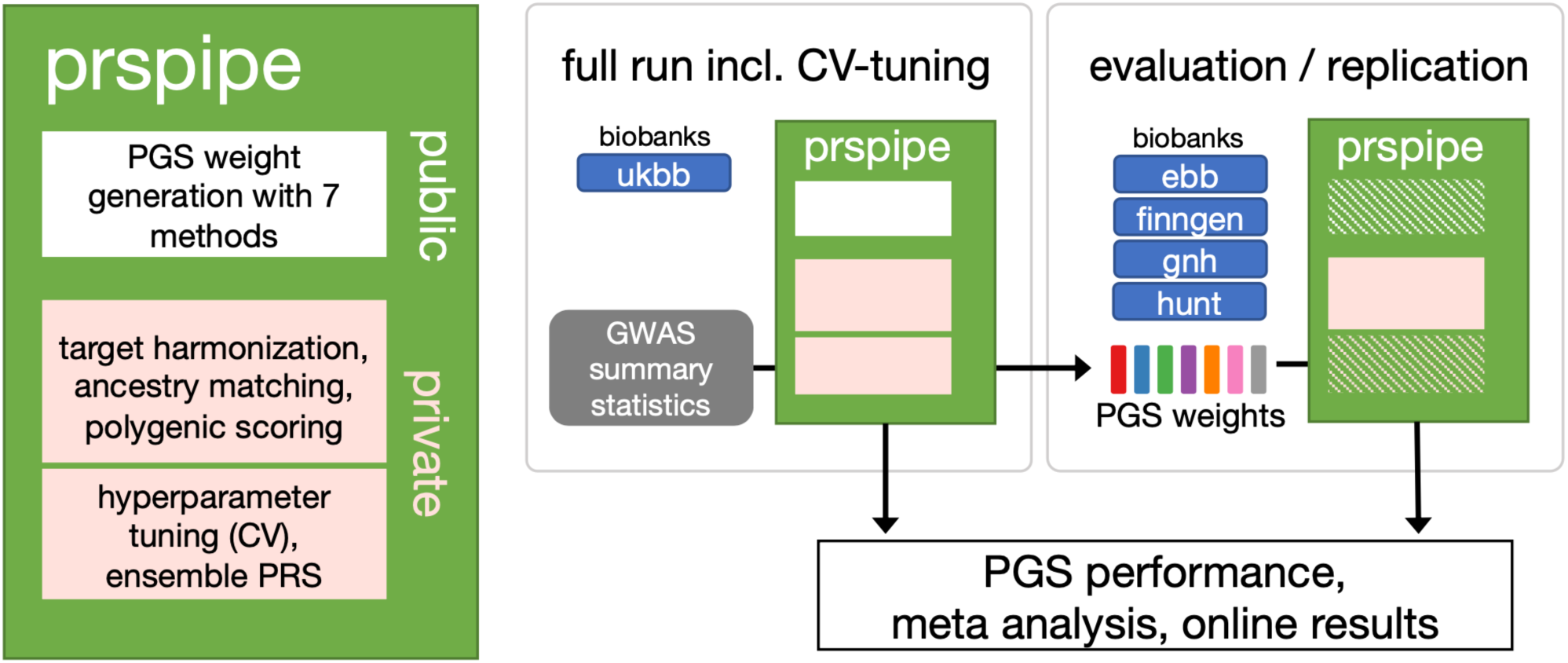
prspipe workflow and application. Prspipe is a snakemake workflow that automates within-biobank method comparisons introduced by Pain et al.^17^ The public stage uses only public data (e.g., summary statistics, ancestry reference, PGS software) to derive PGS weights using seven methods from GWAS summary statistics. The private stage requires access to target genotype and phenotype data and includes data harmonization, polygenic scoring, and PGS tuning using cross-validation (CV). We used prspipe to generate PGS weights and tune hyperparameters in the UKBB EUR data (full run). PGS weights were shared with other biobanks for evaluation/replication (skipping the public stage). Other biobanks were not used for hyperparameter tuning. Downstream analyses were conducted to determine PGS performance using a meta-analytical framework (not part of the workflow), and results were published as an online resource at https://methodscomparison.intervenegeneticscores.org.

### Browsable results, meta-analysis and ranking

As outlined in Figure 2 for T2D, we calculated PGS effect sizes for continuous and binary traits across all target biobanks and ancestries (Supplementary Tables 1-3, Supplementary Figures 1-5) and performed mixed model meta-analyses within ancestry groups to determine the best performing PGS (the one with the largest effect size) for each trait across biobanks (Supplementary Table 4-6). Additionally to scores produced by single methods, we evaluated the UKBB-tuned ensemble PGS in other biobanks after projecting them back to the variant-level (Methods). For each trait, we meta-analyzed β-coefficients (i.e., the change in the trait per PGS standard deviation, on the log-odds scale for binary traits, in standard deviations for continuous traits) of up to 13 PGS corresponding to different tuning types (auto or CV) for seven methods and the UKBB-EUR-tuned ensemble PGS (Supplementary Figure 6).

**Figure 2:**
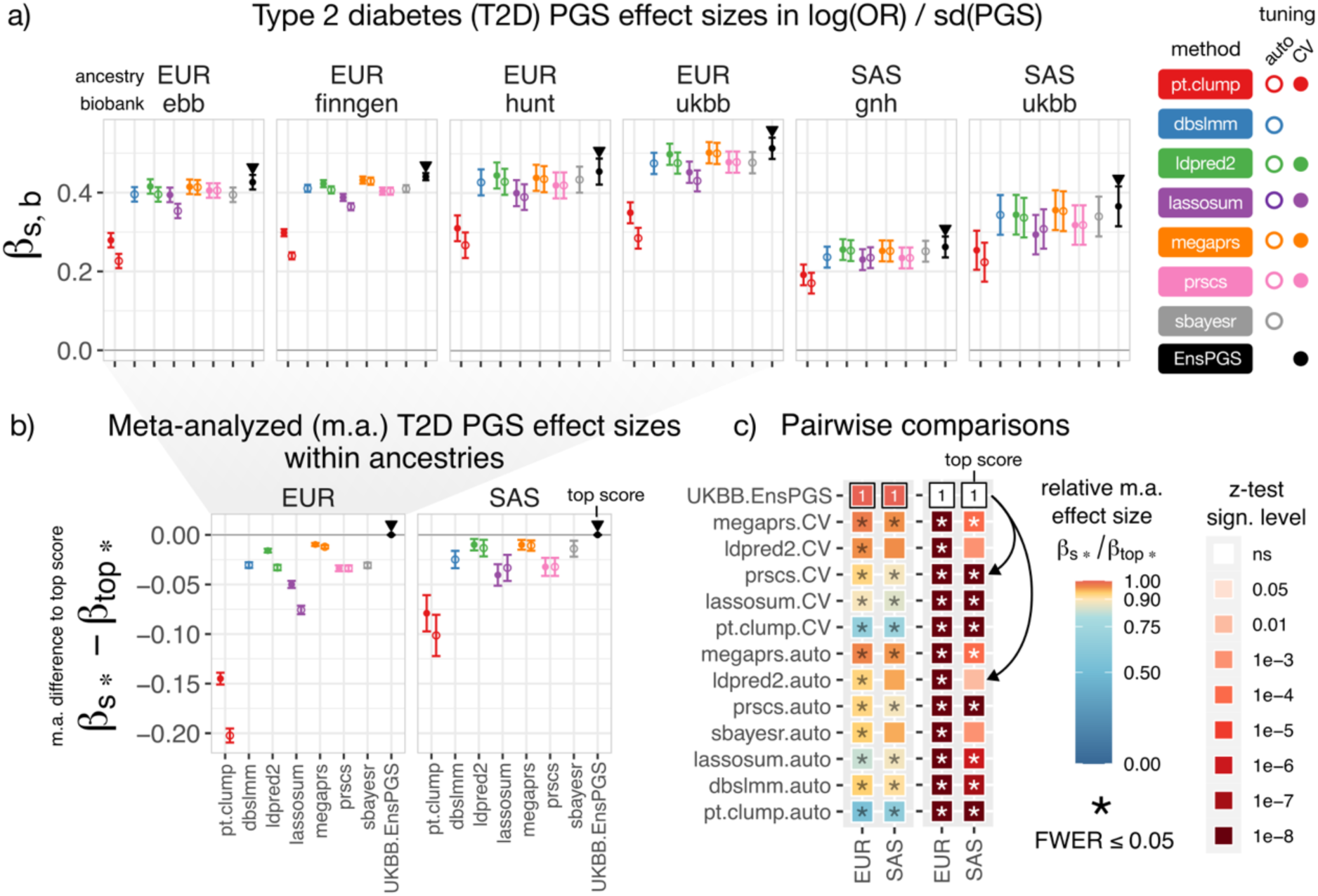
Meta-analysis workflow for methods comparison, example: type 2 diabetes. a) PGS effect sizes β_s,b_ (i.e., the change in log odds-ratio per PGS standard deviation measured for scores “s” across biobanks “b”, see Methods) with 95% confidence intervals for all PGS methods (x-axis) stratified by biobank, replicated ancestries (EUR, SAS) and tuning types (auto, CV) serve as the inputs for the meta-analysis (shown for example trait type 2 diabetes). We evaluated scores for seven methods shown on the right, as well as the UKBB-EUR-tuned ensemble PGS (EnsPGS). The largest effect size for each ancestry and biobank is marked with a triangle (given by the ensemble in all cases). β_s,b_ for all target data and traits are displayed in Supplementary Figure 1 and browsable online. b) PGS effect-sizes are meta-analyzed within ancestries across biobanks (yielding a single β_s*_ for each score “s”). Effect-size differences relative to the largest meta-analyzed effect size (β_top*_, given by the ensemble) and 95% confidence intervals are shown. All pairwise differences are available in Supplementary Table 5-6, and browsable online. c) Meta-analyzed effect sizes β_s*_ are compared, and significance testing is performed. Heatmaps show both the effect-size relative to the largest (β_s*_/ β_top*_, left) as well as corresponding two-sided z-test significance levels at which H_0_: β_s*_-β_top*_= 0 can be rejected (right). Significant differences at a FWER <= 0.05 are marked with an asterisk (*), accounting for all 351 tests performed across traits and ancestries. The score against which comparisons are performed with effect size β_top*_ is marked with a “1” and black border. Arrows indicate two example comparisons: against PRScs-CV (significant difference in SAS and EUR) and LDpred2-auto (significant only in EUR). Data for all PGS and traits are provided in Supplementary Figure 6

Other than the ensemble PGS, we found that CV-tuned PGS from LDpred2 and MegaPRS ranked highly across traits (Figure 3). The median relative increase in PGS effect size over CV-tuned pT+clump was 29.2% for CV-tuned LDpred2 (mean 30.9%±10.3sd, N=12, EUR) and 29.9% for CV-tuned MegaPRS (mean 31.2%±12.6 sd, N=12, EUR), showing overall comparable performance (the median relative difference between the two was 0.1% in favor of MegaPRS).

**Figure 3:**
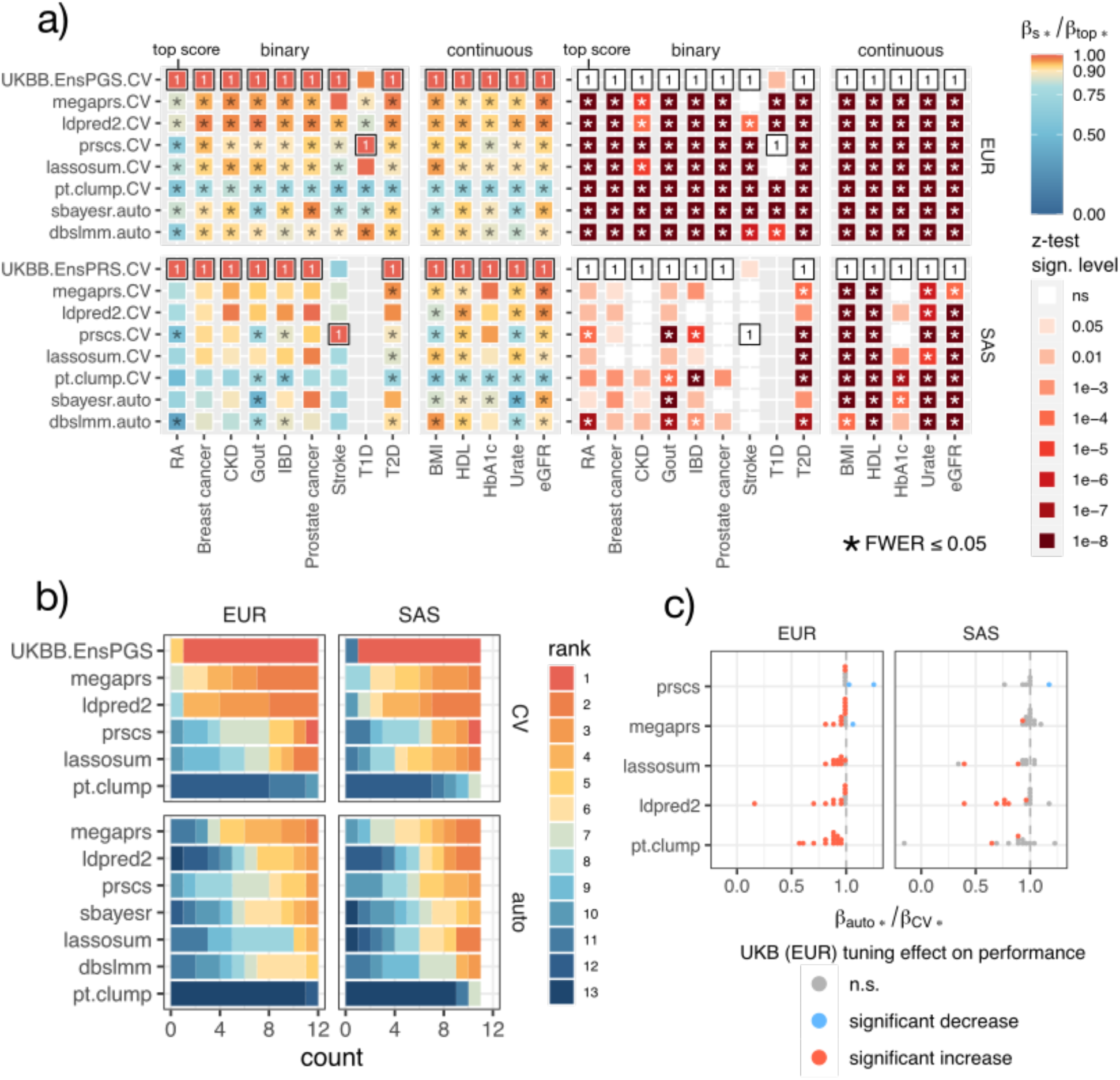
Relative meta-analyzed PGS effect sizes across 14 traits. a) For the 14 traits for which we tuned hyperparameters using CV (x-axis), we show heatmaps of meta-analyzed β-coefficients relative to the highest within traits (β_s*_/β_top*_, left) as well as significance levels for the two-sided z-test (H_0_: β_s*_- β_top*_= 0, right) stratified by ancestry (EUR, SAS). The top score with the largest effect size for each trait (β_top*_) is marked with a “1” and black box. Differences significant at FWER <= 0.05 are marked with asterisks (*), accounting for all 351 pairwise tests performed across traits, replicated ancestries and tuning types (auto, CV) (full data for all traits and sores are displayed in Supplementary Figure 6). b) Bar plot counting PGS ranks across traits (1 is the highest), stratified by ancestry, method, and tuning-type (auto, CV). Methods are ordered by the average rank-sum across tuning types (highest to lowest). c) For each method (y-axis), dot plots showing the relative meta-analyzed effect-size of the score derived using methods’ automatic settings against the CV-tuned scores (β_auto*_/β_CV*_). Colors denote sign and significance of the two-sided z-test (H_0_: β_CV*_-β_auto*_= 0) at FWER <= 0.05 after accounting for 114 tests across traits and ancestries (Supplementary Figure 12-13). Methods are ordered by the median difference.

Scores produced by automatic tuning appeared overall less reliable, especially for LDpred2 (see Discussion) and SBayesR (as previously described^17^). Although automatic tuning typically outperformed the baseline method pT+clump, we observed seemingly non-systematic cases of reduced relative performance (e.g., SBayesR for urate/gout, DBSLMM for Alzheimer’s disease, or LDpred2 for HbA1c or RA) (Supplementary Figure 11).

MegaPRS was the best automatically tuned method (median 23.3% relative increase over CV-tuned pT+clump, mean 27.4%±15.9 sd, EUR), yet PGS effect sizes were comparatively low for some continuous traits (e.g., BMI, HDL, or height, Supplementary Figure 11).

Effect size differences between the top PGS by single methods were mostly not significant (Supplementary Figure 7, FWER <= 0.05, two-sided z-test). We also provide these data on the level of individual biobanks (Supplementary Figure 9-10), revealing that the best single method was not necessarily consistent between biobanks.

### UKBB-tuned ensemble PGS outperforms other methods

The ensemble PGS ranked favorably for all traits in EUR- and SAS-matched target data (Figure 3) except for T1D in EUR (driven by lower performance in FinnGen) and stroke in SAS (the trait with the overall lowest performance). For EUR target data, effect sizes were significantly greater than those of all other PGS for 6/9 binary and 5/5 continuous traits (FWER <= 0.05, two-sided z-test) and the largest overall in 13/14 traits for which we fit ensemble PGS. These results stood out compared to those for single methods, which did not produce a consistent best method and for which differences were mostly not significant.

Compared to the best single methods, the median relative increase in effect size was 3.7% over CV-tuned LDpred2 and 4.5% over CV-tuned MegaPRS for binary disease traits (N=9). Median relative increases for continuous traits were larger (5.2% and 7.9%, respectively, N=5). When measured in terms of variance explained, relative differences were larger. We observed median relative increases of 7.4% and 9% for binary traits (liability scale) and 10.7% and 16.1% for continuous traits, respectively (Supplementary Figure 8, Supplementary Table 7). Similar trends were observed for SAS target data, with the ensemble PGS having the largest effect size in 12/13 traits, albeit its effect size was only significantly larger than all others for continuous traits urate, eGFR and HDL (FWER<=0.05, two-sided z-test). We report relative effect sizes of all methods relative to the ensemble PGS in Table 3.

**Table 3:** PGS meta-analyzed β coefficients relative to the ensemble PGS (β_s*_/ β_EnsPGS*_) For the 14 traits for which we tuned hyperparameters with CV, relative PGS effect sizes relative the ensemble PGS are shown (β_s*_/β_EnsPGS*_) stratified by PGS method, tuning type (CV/auto), ancestry (EUR, SAS) and type of trait (binary/continuous). The number of traits (N), medians, means and standard deviations (sd) are shown. Methods are ordered by the median EUR relative effect size within traits and tuning types.

| method | tuning type | trait | N (EUR) | N (SAS) | median (EUR) | median (SAS) | mean (EUR) | sd (EUR) | mean (SAS) | sd (SAS) |
| --- | --- | --- | --- | --- | --- | --- | --- | --- | --- | --- |
| **ldpred2** | CV | binary | 9 | 8 | 0.965 | 0.963 | 0.943 | 0.045 | 0.972 | 0.127 |
| megaprs | CV | binary | 9 | 8 | 0.957 | 0.934 | 0.947 | 0.041 | 0.958 | 0.124 |
| lassosum | CV | binary | 9 | 8 | 0.921 | 0.914 | 0.913 | 0.061 | 0.920 | 0.111 |
| prscs | CV | binary | 9 | 8 | 0.903 | 0.900 | 0.896 | 0.100 | 0.909 | 0.259 |
| pt.clump | CV | binary | 9 | 8 | 0.735 | 0.734 | 0.721 | 0.077 | 0.748 | 0.186 |
| **megaprs** | auto | binary | 9 | 8 | 0.948 | 0.950 | 0.933 | 0.050 | 0.964 | 0.104 |
| ldpred2 | auto | binary | 9 | 8 | 0.927 | 0.955 | 0.838 | 0.265 | 0.904 | 0.314 |
| prscs | auto | binary | 9 | 8 | 0.925 | 0.876 | 0.915 | 0.052 | 0.877 | 0.264 |
| sbayesr | auto | binary | 9 | 8 | 0.907 | 0.895 | 0.873 | 0.083 | 0.841 | 0.181 |
| dbslmm | auto | binary | 9 | 8 | 0.904 | 0.865 | 0.890 | 0.092 | 0.815 | 0.199 |
| lassosum | auto | binary | 9 | 8 | 0.891 | 0.870 | 0.861 | 0.103 | 0.753 | 0.262 |
| pt.clump | auto | binary | 9 | 8 | 0.629 | 0.627 | 0.607 | 0.098 | 0.527 | 0.302 |
| **ldpred2** | CV | continuous | 5 | 5 | 0.950 | 0.936 | 0.948 | 0.016 | 0.925 | 0.049 |
| megaprs | CV | continuous | 5 | 5 | 0.927 | 0.931 | 0.940 | 0.024 | 0.946 | 0.034 |
| prscs | CV | continuous | 5 | 5 | 0.923 | 0.926 | 0.909 | 0.031 | 0.875 | 0.090 |
| lassosum | CV | continuous | 5 | 5 | 0.906 | 0.920 | 0.914 | 0.026 | 0.916 | 0.021 |
| pt.clump | CV | continuous | 5 | 5 | 0.735 | 0.729 | 0.743 | 0.037 | 0.718 | 0.048 |
| **prscs** | auto | continuous | 5 | 5 | 0.923 | 0.928 | 0.907 | 0.028 | 0.903 | 0.076 |
| dbslmm | auto | continuous | 5 | 5 | 0.901 | 0.904 | 0.891 | 0.039 | 0.897 | 0.066 |
| sbayesr | auto | continuous | 5 | 5 | 0.887 | 0.862 | 0.868 | 0.075 | 0.806 | 0.184 |
| megaprs | auto | continuous | 5 | 5 | 0.883 | 0.907 | 0.885 | 0.067 | 0.922 | 0.055 |
| lassosum | auto | continuous | 5 | 5 | 0.873 | 0.890 | 0.877 | 0.021 | 0.901 | 0.054 |
| ldpred2 | auto | continuous | 5 | 5 | 0.851 | 0.778 | 0.823 | 0.121 | 0.781 | 0.114 |
| pt.clump | auto | continuous | 5 | 5 | 0.643 | 0.614 | 0.606 | 0.088 | 0.635 | 0.112 |

### CV-tuning increases PGS robustness

Hyperparameter tuning with cross-validation using the UKBB EUR data was often beneficial and rarely harmful when evaluated on EUR target data (Figure 3). CV-hyperparameter tuning strongly increased effect sizes in a subset of traits for specific methods, rather than providing large benefits across traits (Supplementary Figure 12-13). pT+clump benefited most from CV-tuning when evaluated on EUR target data, i.e., selecting p-value thresholds larger than the baseline 1e-8 was always beneficial (median 12.8% increase in effect size), followed by lassosum (median 6.2% increase) and LDpred2 (median 4.1% increase). MegaPRS and PRScs benefited the least (median 1.2% and 0.2% increase, respectively). For SAS target data, the median benefits were smaller, except for PRScs (Supplementary Table 8) and overall less consistent (Figure 3). Mean increases were larger for all methods except for PRScs in EUR target data, often dominated by few instances in which automatic tuning had comparatively low performance.

The performance increases seen by CV-tuning were by and large significant when evaluated in EUR target data (Figure 3, FWER <= 0.05, two-sided z-test), except for PRScs which only saw an improvement for two phenotypes. Significant negative effects of CV-tuning for EUR data were only observed for RA (PRScs and MegaPRS, driven by FinnGen) and CKD (PRScs, Supplementary Figure 12). For SAS target data, we observed fewer significant differences, and PRScs was the only method for which we observed a significant reduction in effect size (BMI, Supplementary Figure 13). A more detailed description of these comparisons is provided in the Supplementary Results.

### Tuned PGS performance varies more between biobanks than between methods within biobanks

We estimated PGS effect size heterogeneity between biobanks and how it compares to the heterogeneity between methods within biobanks using 3-level meta-analytic random effects models in EUR target data (Methods). These nested models have two random effect parameters: τ^2^_biobank_ and τ^2^_method_. τ^2^_biobank_ captures effect-size heterogeneity due to differences between biobanks, and τ^2^_method_ captures heterogeneity due to differences between methods within biobanks (we report their square roots τ_biobank_ and τ_method_, as they are on the same scale as the PGS effect sizes). Additionally, we estimated I^2^_biobank_ and I^2^_method_, which quantify the overall fraction of variance (between 0 and 1) in effect sizes attributable to biobank or choice of method within biobank, respectively.

We focused on scores selected via cross-validation in the UKBB-EUR sample (if available) and excluded scores from SBayesR that performed poorly in the UKBB-EUR 80% training data (RA, T1D, BMI, urate/gout). We did not consider the ensemble PGS or baseline method pT+clump, meaning that up to 6 scores were considered per trait. This setting was chosen to mimic the case in which multiple validated PGS from standard methods are available.

We found significant heterogeneity of PGS effect sizes in all 13 traits replicated in at least two biobanks (FWER<=0.05, Cochran’s *Q*-test, accounting for 13 tests). The target biobank had a larger influence on the PGS effect size than the choice of method within biobank across all traits (i.e., τ_method_ < τ_biobank_, Figure 4, Table 4). However, likelihood-based 95% confidence intervals for τ_biobank_ were large and sometimes included the estimate for τ_method_ (RA, stroke, T2D) and 0 (T1D, breast cancer). The variation in PGS effect sizes could to a large degree be explained by heterogeneity between biobanks (average I^2^_biobank_ = 82.9% ±14.3 sd, N = 13) and, to a lesser degree by heterogeneity between methods (average I^2^_method_ = 11.97% ±12.4 sd, N=13).

**Figure 4:**
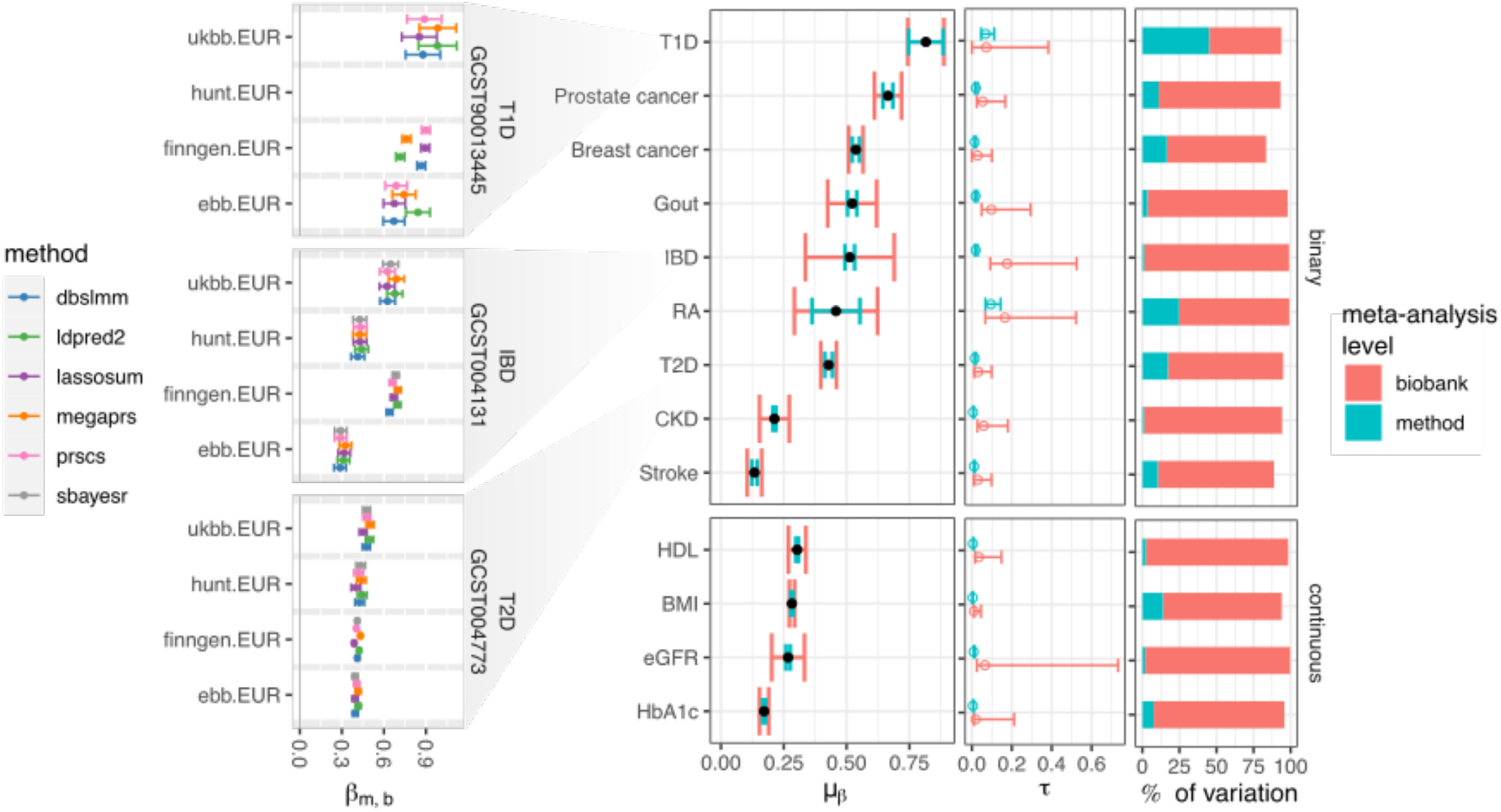
3-level meta-analysis of PGS effect sizes in EUR target data. For all 13 traits replicated in at least 2 biobanks in EUR ancestry target data and CV-tuned in UKBB, from left to right: 1) PGS effect sizes (β-coefficients, β_m,b_) with 95% confidence intervals for three example traits within biobanks (T1D: high variability between methods, IBD: high variability between biobanks, T2D: intermediate to low variability between methods and biobanks), 2) the meta-analyzed average effect sizes across biobanks and methods (μ_β_) with bars denoting the square roots of the variance components (τ), i.e., the standard deviations of the random intercepts for biobanks or methods, 3) τ-values with likelihood-based 95% confidence intervals and 4) I^2^ estimates, i.e., the fraction of variance of effect sizes explained by heterogeneity between biobanks or methods within biobanks. τ and I^2^ are colored according to the levels of the meta-analytic 3-level random effects model (Methods).

**Table 4:** 3-level meta-analytical random effects model results (EUR) Table corresponding to Figure 4. From left to right, the target trait, the number of biobanks with the trait (N_biobank_), the number of methods/scores considered (N_method_), the meta-analyzed average PGS effect size across methods/scores and biobanks (μ_β_) with standard deviation (sd), the standard deviation of the random intercepts specific to biobanks (τ_biobank_) including 95% likelihood-based confidence intervals (95% CI), the standard deviation of the random intercepts specific to methods within biobanks (τ_method_) including 95% CI, the fraction of total effect size variance due to heterogeneity between biobanks (I^2^_biobank_) in %, and the fraction of the total effect size variance due to heterogeneity between methods (I^2^_method_) in %. Endpoints are ordered by type (binary/continuous) and μ_β_. SBayesR was excluded for RA, T1D, gout, and BMI (*). Full results for EUR and SAS are given in Supplementary Table 9-12

| trait | N<sub>biobank</sub> | N<sub>method</sub> | $\mu_{\beta} \pm \text{sd}$ | $\tau_{\text{biobank}}$ (95% CI) | $\tau_{\text{method}}$ (95% CI) | I<sup>2</sup><sub>biobank</sub> (%) | I<sup>2</sup><sub>method</sub> (%) |
| --- | --- | --- | --- | --- | --- | --- | --- |
| **T1D** | 3 | 5* | 0.815 ± 0.05 | 0.072 (0-0.383) | 0.069(0.046-0.112) | 48.8 | 45 |
| **Prostate cancer** | 4 | 6 | 0.664 ± 0.029 | 0.054 (0.024-0.167) | 0.02(0.015-0.029) | 82.2 | 11 |
| **Breast cancer** | 4 | 6 | 0.537 ± 0.017 | 0.029 (0-0.1) | 0.014(0.012-0.02) | 67.1 | 16.5 |
| **Gout** | 4 | 5* | 0.522 ± 0.05 | 0.098 (0.049-0.294) | 0.018(0.014-0.028) | 94.7 | 3.3 |
| **IBD** | 4 | 6 | 0.513 ± 0.089 | 0.177 (0.092-0.525) | 0.019(0.015-0.028) | 97.8 | 1.1 |
| **RA** | 4 | 5* | 0.458 ± 0.087 | 0.165 (0.067-0.522) | 0.095(0.068-0.144) | 74.4 | 24.7 |
| **T2D** | 4 | 6 | 0.428 ± 0.017 | 0.031 (0.013-0.099) | 0.015(0.012-0.02) | 77.8 | 17.2 |
| **CKD** | 4 | 6 | 0.213 ± 0.031 | 0.059 (0.028-0.18) | 0.006(0.006-0.01) | 93.4 | 0.9 |
| **Stroke** | 4 | 6 | 0.133 ± 0.016 | 0.029 (0.01-0.099) | 0.01(0.01-0.016) | 78.5 | 10.4 |
| **HDL** | 3 | 6 | 0.303 ± 0.02 | 0.035 (0.016-0.148) | 0.005(0.005-0.009) | 96.2 | 2.3 |
| **BMI** | 3 | 5* | 0.282 ± 0.006 | 0.01 (0.01-0.046) | 0.004(0.004-0.004) | 80.2 | 13.8 |
| **eGFR** | 2 | 6 | 0.267 ± 0.046 | 0.065 (0.025-0.733) | 0.009(0.009-0.015) | 97.9 | 1.8 |
| **HbA1c** | 2 | 6 | 0.172 ± 0.013 | 0.018 (0.011-0.211) | 0.005(0.005-0.009) | 88.3 | 7.5 |

Effect sizes for inflammatory bowel disease and RA varied most between biobanks (τ_biobank_), also when adjusting for the average effect size (τ_biobank_/μ_β_). Effect sizes for BMI varied the least between biobanks, both absolutely (τ_biobank_) and relative to the average effect size (τ_biobank_/μ_β_). For binary traits, effect sizes for breast cancer varied the least across biobanks, both absolutely and relative to the average effect size.

Across traits, τ_method_ was correlated with the average effect size (Pearson correlation 0.54, p = 0.0558, t-statistic = 2.1377, 11 degrees of freedom), especially when removing T1D and RA (Pearson correlation 0.85, p = 0.00094, t-statistic = 4.83, 9 degrees of freedom), i.e., we found a linear relationship between the differences between methods and overall effect size, especially in the set of non-autoimmune traits.

τ_biobank_ was less correlated with the meta-analyzed average effect size (Pearson correlation 0.367, p = 0.219, t-statistic = 1.30, 11 degrees of freedom), i.e., large PGS effect sizes weren’t necessarily associated with higher variability between biobanks.

For SAS ancestry target data, we did not find significant heterogeneity of PGS effect sizes in CKD, stroke, prostate cancer, or breast cancer (FWER<=0.05, Cochran’s *Q*-test, accounting for 11 tests) and τ_biobank_ could never reliably be estimated (Supplementary Figure 14).

Likelihood-based 95% confidence intervals for τ_biobank_ included 0 for 9/11 replicated traits (all but T2D and eGFR).

### High variability between PGS methods for autoimmune diseases

Effect sizes were most variable between methods for autoimmune diseases T1D and RA (τ_method_) (Supplementary Figure 15), even when accounting for the average effect size in those traits (τ_method_/μ_β_), or relative to the total variation of effect sizes (I^2^_method_). The scores for T1D and RA also had the largest fraction of PGS variance originating in the HLA region (mean 0.7 ± 0.12 sd and 0.54 ± 0.21 sd, respectively) (Supplementary Figure 16). For T1D, the method with the largest effect size appeared to be biobank-specific, with FinnGen favoring PRScs, while the UKBB and EBB had significantly larger effect sizes for LDpred2 and MegaPRS (forest plots for all 3-level meta-analytic models are available in the Supplementary Data). In contrast, effect sizes for BMI and HDL varied the least between methods.

Regarding SAS-matched target data, RA was only available in GNH with a limited number of cases (60) but displayed the highest heterogeneity of effect sizes due to method (τ_method_ = 0.137, 95% CI: 0.046-0.379), consistent with the findings in EUR ancestry. T1D scores were not predictive in GNH (Supplementary Figure 1), and not evaluated in the UKBB due to small sample size, therefore, we couldn’t replicate the related findings from the EUR subset.

## Discussion

With this study, we have provided to our knowledge the most comprehensive systematic PGS method comparison to date. By publishing our workflow, we aim to increase access to PGS methods and facilitate future research. We believe that PGS method software could be greatly improved by support for standard formats (e.g., those maintained by the GWAS Catalog and PGS Catalogs^28^) alongside software containerization (containers were supported in all the research environments that contributed to this study).

Our analysis was based on a previously published framework^17^ which we automated, and expanded application and evaluation to multiple biobanks. Methods explicitly tailored for diverse target populations or source GWAS were missing in this framework, and diverse ancestries were not well represented in our target data, which provides a limitation of this study. PGS tuning was performed in one biobank (UKBB-EUR) relying largely on author-provided LD reference panels. This approach more closely resembles real-world PGS application and allowed us to harness the full sample sizes in other biobanks/ancestries to maximize statistical power, and test transferability.

Importantly, we were unable to identify a single method that consistently outperformed all others (not counting the ensemble PGS), and the two highest performing methods (CV-tuned MegaPRS and LDpred2) were virtually tied. The best automatically tuned method was MegaPRS, albeit like other automatic methods it suffered sporadic cases of comparatively lower performance. Which method performs best may vary based on the specifics of the GWAS summary statistics, trait, and target sample. Given that the best methods performed so similarly, other modelling choices not investigated here (such as the set of included variants and their availability in the target sample) may well tip the balance in favor of one or the other when starting from the same GWAS summary statistics. Based on our results, we recommend tuning with cross-validation (with sufficiently large ranges of hyperparameters) instead of using methods’ automatic settings, primarily to prevent cases of comparatively lower performance, rather than providing large improvements across traits.

One reason for the lower performance of automatic tuning could be model misspecification, e.g., mismatched LD-references, or misreported fields in the input summary statistics. These inconsistencies may not be considered when tools are developed. The variable performance of LDpred2-auto stood out particularly against the high performance of CV-tuned PGS from same method. We note that LDpred2-auto has been updated at the time of writing including an optional new parameterization, which could affect its performance^29^.

This highlights a challenge faced by any method comparison: The frequent emergence of new tools and methods means that comparisons risk becoming outdated shortly after execution. Method evaluation across multiple biobanks can hardly match the pace of new developments. We therefore caution against using the results of this study to make definitive claims about relative method performance of actively developed methods. A more sustainable approach to method comparisons would be decentralized, with researchers individually submitting performance estimates for published scores (starting from the same summary statistics and variants) to a central repository and receiving credit by having such submissions be referenceable.

Using meta-analytic mixed models, we found that the performances of well-tuned PGS varied more between biobanks than within biobanks. This likely reflects heterogeneity in phenotyping (e.g., disease diagnosis practices) rather than differences in population structure or genotyping. Effect sizes for BMI, which presumably is consistently measured, varied the least between biobanks, supporting this hypothesis. The variability between biobanks for some traits implies that scores need to be re-evaluated when switching between different target data even when comparing ancestry-matched populations.

We note that the parameters by which we quantified variability are sensitive to which biobanks and PGS are included. The setting we chose mimics the case in which multiple UKBB-EUR-validated PGS are available. The variability between methods could increase if poorly performing (non-validated) scores are included in the analysis. On the other hand, the variability between biobanks could decrease if, e.g., phenotype definitions were further refined.

We found particularly large differences between methods for autoimmune diseases T1D and RA. This could be driven by the way methods handle the HLA-region, as well as genotyping differences in the target biobanks. Our analyses highlight modelling of the HLA-region as an area in which methods could potentially be improved.

One of the most useful insights from this study is that ensemble PGS tuned in the UKBB-EUR sample provided consistently strong performance, albeit at the cost of higher computational demand during training. We see this method as complimentary to cross-trait prediction strategies (MultiPGS)^30–32^ that use PGS constructed from multiple sets of GWAS summary statistics (from different traits). Considering the small differences in performance we observe for well-tuned scores from single methods, we see ensemble PGS and MultiPGS as promising avenues to further improve PGS performances beyond what is currently possible with single methods. Future research needs to assess how well EUR-trained ensemble PGS transfer to other genetic ancestries. It is possible that training needs to be performed in a population similar to the target population to ensure optimal performance and avoid exacerbating already existing issues with current PGS^7^.

In Summary, while no single method outperformed all others, method ensembles provided consistently strong performance (with few exceptions). PGS effect size heterogeneity between biobanks was larger than between methods within biobanks, likely pointing to challenges with phenotyping. Large heterogeneity between methods was observed for autoimmune diseases, indicating that special care should be taken for PGS which rely heavily on the HLA region. Our open-source workflow, analyses framework and online results provide a rich ground for future method benchmarking and development.

## Methods

### Participating Studies

Data from five biobanks were considered: The UK Biobank, FinnGen, Estonian Biobank, Trøndelag Health Study (HUNT) and Genes & Health. All biobanks independently performed genotyping, imputation, and variant quality control.

### GWAS summary statistics selection and processing

We selected summary statistics from the GWAS catalog for eight binary traits and for five continuous traits. Table 1 shows GWAS catalog study identifiers and traits. Where available, we directly used the pre-harmonized summary statistics provided by the GWAS catalog. For GWAS catalog studies GCST90013445^33^ (T1D), GCST008972^34^ (urate), GCST007954^35^

(HbA1c) and GCST004773^36^ (T2D) we used the MungeSumstats R package^37^ (version 1.0.1) to retrieve missing fields (e.g., variant positions). GWAS variants were matched to the HapMap3-1KG variants based on positions and allele codes and renamed accordingly. Other quality control steps are flipping of variants to match the HapMap3-1KG reference, variant frequency filtering (>1%), removal of variants with invalid p-values (>1 or <0), ambiguous variants, variants with missing data, duplicate variants, or variants with sample size more than three standard deviations away from the median per-variant sample size (if available), as previously described^17^.

We selected GWAS studies with predominantly European ancestry discovery samples, because the evaluated biobanks primarily contain individuals of European ancestry. Because we use the UKBB for evaluation and score selection, we selected for studies with large sample sizes that preferably did not include the UKBB in the discovery sample. Yet, the selected summary statistics for AD and height included the UKBB-EUR data, which was then excluded from performance estimation for those phenotypes, from hyperparameter tuning with cross validation, and ensemble PGS.

### Reference genotype harmonization

We constructed our own definition of the HapMap3-variants^38^ to avoid favoring one of the definitions used by the PGS methods. We retrieved HapMap3 variant rsids, and downloaded genotypes for the 1000 Genomes reference from ftp://ftp.1000genomes.ebi.ac.uk/vol1/ftp/technical/working/20140708_previous_phase3/v5_vcfs/ (v5). We retrieved updated rsids for all 1000 Genomes variants using the Bioconductor SNPlocs.Hsapiens.dbSNP144.GRCh37 R-package^39^ (the latest version for the GRCh37 genome-build at the time) based on GRCh37 variant positions and allele codes, and intersected them with the HapMap3 variants based on rsids. We then mapped these variants from GRCh37 to GRCh38 using liftOver^40^ and retrieved rsids in that genome build too, based on location and allele codes, using the SNPlocs.Hsapiens.dbSNP151.GRCh38 R-package^39^ (the latest version for the GRCh38 genome-build at the time). We retained variants with an allele frequency of at least 1% in any of the 1000 Genomes superpopulations. These variants (HapMap3-1KG, N = 1,330,821) form the basis for subsequent analyses.

The list of variants including GRCh37 (hg19) and GRCh38 (hg38) coordinates, rsids, and allele frequencies in the 1000 Genomes superpopulations is available on https://github.com/intervene-EU-H2020/prspipe/blob/main/resources/1kg/1KGPhase3_hm3_hg19_hg38_mapping_cached.tsv.gz. Scripts to reproduce these steps are available as part of the prspipe workflow (https://github.com/intervene-EU-H2020/prspipe/blob/main/workflow/rules/1kg_hm3_processing.smk).The filtered and intersected 1000 Genomes genotypes are provided as a separate resource. Variants are further filtered when constructing polygenic scores, as described below.

### Target genotype harmonization

Target genotype data were intersected with the HapMap3-1KG variants based on positions and allele codes, renamed, and converted to plink1 format. The harmonized data served as input to all subsequent analyses involving target genetic data, i.e., ancestry reference matching and polygenic scoring. Target harmonization is part of the prspipe workflow and corresponding steps are defined in https://github.com/intervene-EU-H2020/prspipe/blob/main/workflow/rules/genotype_harmonization.smk.

### Binary disease phenotype harmonization

We used expert curated ICD-code based definitions^20,41^ developed at FinnGen to define binary disease traits (referred to as endpoints). Individuals were counted as cases for a specific endpoint if they matched ICD-9-or ICD-10-code-based inclusion/exclusion criteria (Supplementary Table 13). The remaining (non-matching) individuals for that endpoint were counted as controls. All data used to define binary disease endpoints were registry based. breast cancer was only evaluated when the reported sex was female, and prostate cancer only when the reported sex was male.

For the UKBB, we considered both main (data-fields 41202 and 41203) and secondary (data-fields 41204 and 41205) ICD-9 and ICD-10 diagnosis codes derived from hospital inpatient admissions.

### Continuous trait definitions

For the UKBB, we used the following data-fields to define continuous traits: 50 for height, 21001 for BMI, 30700 for creatinine, 30750 for HbA1c and 30880 for urate.

For GNH, we considered all instances where a continuous trait was measured per individual through their primary and secondary health records. We removed outliers based on a 6SD deviation per trait and calculated the mean value per trait per individual to use in the analysis. For HUNT, the latest value was chosen when continuous traits were measured at more than one baseline enrollment or sub-study screening over three recruitment waves since 1984.

Standard quality assessment measures were taken across variables and are described at the HUNT Databank (https://hunt-db.medisin.ntnu.no/hunt-db/variablelist). BMI was defined by height and weight measured at screening. HDL and creatinine were measured from serum in non-fasting individuals. HbA1C was measured in mmol/mol according to the international federation of clinical chemistry (IFCC) standard.

For Estonian Biobank, the earliest value available were chosen for BMI and height, as some individuals are repeatedly measured. Height values larger than 260 or smaller than 100 were omitted. Similarly, BMI values less than 10 or larger than 200 were discarded. Metabolic profiles for HDL and creatinine were obtained with NMR for a random subset of Estonian biobank (n=10681).

For each biobank in which creatinine measurements were available, we calculated the eGFR based according to the diet in renal disease study equation^42^, as follows:

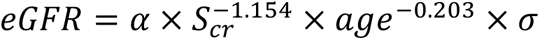

Where α is 30849 if creatinine was measured in μmol/l, or 175 if measured in mg/dl, S_cr_ is the serum creatinine measurement and σ is 0.742 if the reported sex is female, or 1 if sex is male. We did not include the multiplier for “ethnicity” as we only perform comparisons within ancestry-matched populations. During evaluation, all continuous traits are standardized to mean 0 and unit standard deviation.

### Polygenic score weight derivation

We derived polygenic scoring weights with pT+clump, lassosum, PRScs, SBayesR (robust parameterization), LDpred2 and DBSLMM using the settings described previously^17^. For MegaPRS, we used the author-recommended BLD-LDAK heritability model and specified “--model mega” to fit many different scores with different tools (lasso, bolt, ridge, bayesr) and included the HLA region, as recommended. Software versions and sources for each tool are listed in Supplementary Table 14.

Besides letting methods determine suitable hyperparameters based on the summary statistics alone (automatic tuning), we generated scores over grids of hyperparameters for target-data-based tuning with 10-fold cross-validation (see below).

We used European ancestry LD reference panels for all analyses, as the selected GWAS were performed in majority European ancestry samples. In contrast to Pain et al., we use PGS method author-provided LD-references for DBSLMM, lassosum, LDpred2, SBayesR and PRScs. DBSLMM and lassosum LD-references are based on the 1000 Genomes data. For LDpred2, SBayesR, and PRScs they are based on UKBB data. We use the 1000 Genomes EUR-subset to calculate LD when running pT+clump and MegaPRS. Scripts to download PGS method software and data are part of the prspipe workflow. The workflow uses GenoPred scripts to generate plink2-compatible scoring files.

### Ancestry matching and genetic outlier removal

Rather than directly inferring genetic ancestry, we score individuals according to their similarity with groups defined in the 1000 Genomes reference^25^. We use GenoPred Ancestry_identifier.R to project target genetic data into the 1000 Genomes genetic principal component space, and match individuals to one of the five 1000 Genomes superpopulations (AFR, AMR, EAS, EUR, SAS). Both the target and 1000 Genomes genotype data are filtered to variants available in both samples (subset of HapMap3-1KG) with allele frequencies above 5%, missingness below 2%, and variants that do not violate Hardy Weinberg equilibrium (p < 1e-6). Regions of long LD are excluded^43^ and variants are LD-pruned based on the 1000 Genomes reference (using plink “--indep-pairwise 1000 5 0.2”). Genetic PCs are derived in the 1000 Genomes based on the filtered variants, and an elastic net classifier is fit with 5-fold cross validation to place individuals into one of the five groups based on 100 PCs. Target genotype data are projected into the same PC space using plink, and the classifier is used to predict the most matching superpopulation for all individuals.

Additionally, we used GenoPred Population_outlier.R to remove extreme outliers within the assigned ancestry-matched groups in the target data based on the first eight genetic principal components constructed within those groups. We used the same variant filters described above (except that LD-pruning was now performed within the target data), calculate genetic PCs within the assigned groups, and define up to ten centroids in the PC space using R NbClust^44^ (distance = ‘euclidian’, method = ‘kmeans’). For each centroid, the Euclidian distance of individuals to the center is calculated and those with distances that are larger than the 75^th^ percentile + 30 IQR are removed (i.e., extreme outliers). The UKBB was the only biobank that had more than one ancestry group well-represented. Our analyses focus on the replicated groups EUR (UKBB, EBB, HUNT, FinnGen) and SAS (UKBB, GNH).

### Polygenic scoring

We performed polygenic scoring for scores derived by single methods with plink2 using GenoPred Scaled_polygenic_scorer_plink2.R. Polygenic scoring is part of the prspipe workflow. For the evaluation of the ensemble PGS, we performed scoring with plink2. In both cases, missing genotypes are imputed using the 1000 Genomes matched superpopulation allele frequencies as previously described^17^.

### Hyperparameter tuning and ensemble PGS

For the methods that generate scores over a range of hyperparameters (pT+clump, lassosum, PRScs, LDpred2) we used 10-fold cross validation to select the score with the largest correlation with the trait, as described previously^17^. Where available, we included scores produced by methods’ automatic settings in the selection process. We perform cross-validation using 80% of the UKBB EUR data and retain 20% for evaluation (we used different subsamples for each trait in order to perform stratified sampling).

For pT+clump, we define the score given by the p-value threshold of 1e-8 as the automatically tuned score. SBayesR and DBSLMM only use automatic settings, i.e., they produce just a single set of weights and are not tuned with CV.

To fit the ensemble PGS, we include all scores from all methods across hyperparameters, and use 10-fold CV to determine suitable shrinkage parameters for an elastic net model combining the different scores with the caret R-package^45^ (which relies on glmnet^46^). For tuning of the ensemble PGS, we used non-nested scores for pT+clump, i.e., scores with disjoint variant sets corresponding to 10 p-value bins. These steps were performed with GenoPred Model_builder_V2.R and are part of the prspipe workflow.

To score other biobanks with the UKBB ensemble PGS, we generated plink2-compatible scoring files by multiplying the PGS weights of every variant with their corresponding weights in the ensemble PGS model and adding them (yielding a single weight for each variant).

### Performance evaluation within biobanks

All PGS were standardized to mean zero and unit standard deviation within biobanks and ancestries for performance evaluation. We calculated the following metrics for binary disease traits: β coefficients, i.e., the change in log-odds ratios per PGS standard deviation, the change in odds ratio per PGS standard deviation (OR = exp(β)), fraction of variance explained on the observed scale (r^2^_obs_) and the area under the receiver operating characteristic curve (AUROC). The variance explained on the liability scale (r^2^_liab_) was calculated from r^2^_obs_ ^47^ using the median prevalence within ancestries as the population prevalence estimate (Supplementary Table 1). We retrieved DeLong 95% confidence intervals^48^ for the AUROC using the ci.auc-function in the pROC R-package. Confidence intervals for r^2^_obs_ were derived from 1000 bootstrap samples of r_obs_ (the Pearson correlation on the observed scale) for binary traits.

For continuous traits, we calculated β coefficients, i.e., the change in standard deviations of the trait per standard deviation of the PGS and the fraction of variance explained (r^2^_obs_).

When comparing two effect sizes of scores β_a,i_ and β_b,i_ within biobank “i”, we use the two-sided z-test and adjust for the correlation between scores, with test statistic:

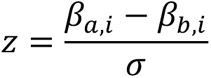

where

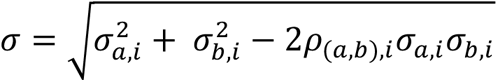

With σ_a,i_ and σ_b,i_ denoting the standard deviations of β_a,i_ and β_b,i_, respectively and ρ_(a,b),i_ denoting the correlation between scores “a” and “b” measured in biobank “i”.

### Data Exclusions before meta-analyses

None of the scores evaluated in GNH-SAS for T1D reached nominal significance (p < 0.05, two-sided z-test) for association with the endpoint (Supplementary Table 1), and all effect sizes were close to zero. We removed these data from further analysis. We found strongly reduced effect sizes of scores for HbA1c in GNH-SAS compared to the UKBB-SAS (Supplementary Table 2, Supplementary Figure 1) and decided not to include these data for meta-analyses (it was unclear if reduced performance was due to a phenotyping issue). We found low effect sizes compared to other biobanks for T1D in HUNT (Supplementary Figure 1), determined it was likely due to a phenotyping issue, and excluded those data from meta-analyses.

### Meta-analysis for methods comparisons

All Meta-analyses were performed in R (version 4.1.1) with the metafor package (rma.mv function, version 3.8-1), using the V-argument to account for the dependence of effect-sizes within biobanks (see below), and models were fit with REML.

We meta-analyzed the β coefficients of scores across biobanks within ancestries and traits using meta-analytic mixed effects models. The observed β coefficients are modelled as follows:

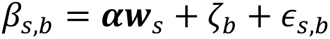

Where β_s,b_ is the observed coefficient for PGS “s” in biobank “b” for a specific trait. β_s,b_ is modelled as a combination of fixed effects (moderators) with realizations **w**_s_ and parameters **α** (bold characters indicate vectors) and two error terms: the sampling error ɛ_b,s_ and a biobank-specific random intercept ζ_b_ (shared by all observed coefficients in that biobank). τ^2^_biobank_ = var(**ζ**) is the random effect, where var(**ζ**) denotes the variance of the biobank-specific random intercepts **ζ**.

For every trait, we meta-analyzed up to 13 PGS in the same model. PGS-choice is modelled with the fixed effects, i.e., **w**_s_ only contains a single non-zero entry of 1 indicating which PGS “s” produced β_s,b_. With this parameterization, parameters in **α** directly correspond to the meta-analyzed effect sizes for the different PGS after inverse variance weighting, and the formula above can equivalently be written as:

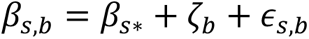

Where β_s*_ is the average effect size for score “s” across all biobanks “*”. To test whether two meta-analyzed effect sizes are significantly different, we compare parameters α_a_ and α_b_ with H_0_: α_a_-α_b_= 0 using the z-test. We also report results for the t-test (produced by the anova function applied to metafor rma.mv objects) in Supplementary Table 5-6.

We retrieved 95% likelihood-based confidence intervals for τ^2^_biobank_ using the confint function (all values are reported in Supplementary Table 4). We further report meta-analyzed AUROC, r^2^_obs_, and r^2^_liab_ values produced by weighting studies by their effective sample size^49^ in Supplementary Table 5-6.

### Method ranking

To rank methods across traits, we considered just the traits for which CV-tuning and ensemble PGS were available (i.e., all except height and AD), and ranked scores based on their meta-analyzed effect sizes β_s*_ (see definition above). To avoid counting scores produced by the same summary statistics twice for eGFR/CKD and urate/gout (e.g., for the ranks shown in Figure 3), we applied the following rule: If the continuous phenotype was available in the same number of biobanks as its binary counterpart, we used the continuous phenotype (higher power), otherwise we used the binary phenotype (larger target diversity). This led to consideration of eGFR for SAS, CKD in EUR, urate for SAS, and gout in EUR. We applied the same reasoning when calculating mean and median values of method performances across all traits.

### 3-level meta-analytic random effects models

For the 3-level meta-analysis, the observed effect-sizes are modelled as follows:

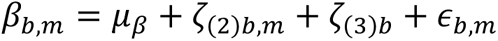

Where β_b,m_ is the observed β-coefficient for method “m” in biobank “b”, μ_β_ is the mean of the distribution of true effect sizes across biobanks and methods, ζ_(2)b,m_ is the within-biobank random intercept due to the choice of method (level 2) and ζ_(3)b_ is the random intercept due to target biobank (level 3, shared by all observations in biobank). The estimated parameter τ^2^_biobank_ = τ^2^_(3)_ = var(**ζ_(3)_**) quantifies the heterogeneity of effect sizes due to target biobank, and τ^2^_method_ = τ^2^_(2)_ = var(**ζ_(2)_**) quantifies the heterogeneity of effect sizes due to choice of method within biobank^50^. In contrast to the model introduced in the previous section, method effects are considered nested within biobanks (independent between biobanks).

All models were fit using restricted maximum-likelihood with the metafor package in R (rma.mv function), using the V-argument to account for the dependence of effect-sizes measured within the same biobanks (see below). We retrieved 95% likelihood-based confidence intervals for τ^2^_biobank_ and τ^2^_method_ using the confint function. To calculate I^2^_biobank_ = I^2^_(3)_ and I^2^_method_ = I^2^_(2)_, we used the implementation provided in dmetar^50^ (var.comp/mlm.variance.distribution function, https://github.com/MathiasHarrer/dmetar/blob/master/R/mlm.variance.distribution.R, commit 21bde652cbae5677b56b0ff848eb96c9bea877d8) based on the three-level extension of the I^2^ metric^51^. I^2^_biobank_ captures the fraction of the overall variance in effect sizes (including sampling error) attributable to biobank (level 3) and I^2^_method_ captures the fraction of the overall variance in effect sizes attributable to methods within biobank (level 2).

### Accounting for dependent effect sizes in meta-analytic models

Within each biobank, ancestry, and trait, we calculated pairwise correlations between polygenic scores based on up to 50,000 randomly sampled individuals. We use the resulting score-score correlation matrix R_b_ (where “b” indicates the biobank) to estimate V_b_, the variance-covariance matrix capturing the dependency of errors of effect size estimates for biobank “b”:

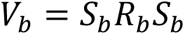

Where S_b_ is a diagonal matrix containing the standard errors of the estimated effect sizes corresponding to the rows/columns of R_b_. The effect sizes measured in different biobanks are considered independent, therefore, the full matrix V supplied to the rma.mv function is a block diagonal matrix containing all V_b_ for the different biobanks b from 1 to n on the diagonal:

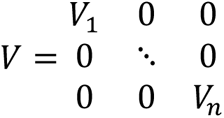

Where 0 denotes a matrix of zeros with the same shape the different V_b_.

### Calculating PGS variance in the HLA-region

For the phenotypes T1D and RA, we scored individuals in the 1000 Genomes EUR subset using either all PGS variants, or only variants contained in the HLA region (defined as the interval 28,000,000-34,000,000 on chromosome 6). The fraction of variance was then computed by dividing the variance of HLA-only PGS by that of the full PGS.

### Data availability and interactive data visualizations

The prspipe workflow used to generate polygenic score weights, perform polygenic scoring and ancestry matching is available on GitHub (https://github.com/intervene-EU-H2020/prspipe).

Genotypes and linked healthcare records held in biobanks are controlled access data and are not publicly available. An application must be made to each biobank to gain access to the data. The 1000 genomes processed genotype data (HapMap3-1KG) are available on figshare (10.6084/m9.figshare.20802700). Non-sensitive experimental data exported from the biobanks are permissively licensed and deposited in an open data repository (https://zenodo.org/doi/10.5281/zenodo.10012995). Processed summary statistics are permissively licensed and hosted on GitHub and accessible through in an R data package (https://github.com/intervene-EU-H2020/pgsCompaR). A website containing an interactive results browser is permissively licensed and available on GitHub (https://github.com/intervene-EU-H2020/pgs-method-compare), hosted at https://methodscomparison.intervenegeneticscores.org/.

Polygenic score weights for scores that were at least nominally significantly associated with the phenotype (p<0.05) in all EUR target data samples are made publicly available through the GWAS catalog (https://www.ebi.ac.uk/gwas/) with publication ID PGP000517. All evaluated scores except the one produced by LDpred2-auto for Arthritis met this threshold. A list of PGS catalog score IDs is provided in Supplementary Table 15.

## Supporting information

forest plots (three level meta-analysis, forest_plots_eur.pdf)

## Data Availability

The prspipe workflow used to generate polygenic score weights, perform polygenic scoring and ancestry matching is available on GitHub (https://github.com/intervene-EU-H2020/prspipe).
Genotypes and linked healthcare records held in biobanks are controlled access data and are not publicly available. An application must be made to each biobank to gain access to the data. The 1000 genomes processed genotype data (HapMap3-1KG) are available on figshare (10.6084/m9.figshare.20802700). Non-sensitive experimental data exported from the biobanks are permissively licensed and deposited in an open data repository (https://zenodo.org/doi/10.5281/zenodo.10012995). Processed summary statistics are permissively licensed and hosted on GitHub and accessible through in an R data package (https://github.com/intervene-EU-H2020/pgsCompaR). A website containing an interactive results browser is permissively licensed and available on GitHub (https://github.com/intervene-EU-H2020/pgs-method-compare) hosted at https://methodscomparison.intervenegeneticscores.org/. Polygenic score weight files have been deposited in the PGS catalog under publication ID PGP000517 (https://www.pgscatalog.org/publication/PGP000517/).

https://github.com/intervene-EU-H2020/prspipe

https://github.com/intervene-EU-H2020/pgsCompaR

https://methodscomparison.intervenegeneticscores.org/

https://www.pgscatalog.org/publication/PGP000517/

## Acknowledgements

This project has received funding from the European Union’s Horizon 2020 research and innovation programme under grant agreement No 101016775, the Hasso Plattner Foundation (HPF) and EMBL-EBI Core Funds. M.I. is supported by core funding from the British Heart Foundation (RG/18/13/33946) and NIHR Cambridge Biomedical Research Centre (BRC-1215-20014; NIHR203312).

This research has been conducted using the UK Biobank Resource under Application Number 78537. Ethics approval for the UK Biobank study was obtained from the North West Centre for Research Ethics Committee (11/NW/0382).

The genotyping in Trøndelag Health Study and work presented here was approved by the Regional Committee for Ethics in Medical Research, Central Norway (2014/144, 2018/1622, 2018/411492). All participants signed informed consent for participation and the use of data in research.

The activities of the Estonian Biobank are regulated by the Human Genes Research Act, which was adopted in 2000 specifically for the operations of the Estonian Biobank. Individual level data analysis in the Estonia Biobank was carried out under ethical approval 1.1-12/624 from the Estonian Committee on Bioethics and Human Research (Estonian Ministry of Social Affairs), using data according to release application S22, document number 6-7/GI/16259 from the Estonian Biobank.

Patients and control subjects in FinnGen provided informed consent for biobank research, based on the Finnish Biobank Act. Alternatively, separate research cohorts, collected prior the Finnish Biobank Act came into effect (in September 2013) and start of FinnGen (August 2017), were collected based on study-specific consents and later transferred to the Finnish biobanks after approval by Fimea (Finnish Medicines Agency), the National Supervisory Authority for Welfare and Health. Recruitment protocols followed the biobank protocols approved by Fimea. The Coordinating Ethics Committee of the Hospital District of Helsinki and Uusimaa (HUS) statement number for the FinnGen study is Nr HUS/990/2017.

The FinnGen study is approved by Finnish Institute for Health and Welfare (permit numbers: THL/2031/6.02.00/2017, THL/1101/5.05.00/2017, THL/341/6.02.00/2018, THL/2222/6.02.00/2018, THL/283/6.02.00/2019, THL/1721/5.05.00/2019 and THL/1524/5.05.00/2020), Digital and population data service agency (permit numbers: VRK43431/2017-3, VRK/6909/2018-3, VRK/4415/2019-3), the Social Insurance Institution (permit numbers: KELA 58/522/2017, KELA 131/522/2018, KELA 70/522/2019, KELA 98/522/2019, KELA 134/522/2019, KELA 138/522/2019, KELA 2/522/2020, KELA 16/522/2020), Findata permit numbers THL/2364/14.02/2020, THL/4055/14.06.00/2020,,THL/3433/14.06.00/2020, THL/4432/14.06/2020, THL/5189/14.06/2020, THL/5894/14.06.00/2020, THL/6619/14.06.00/2020, THL/209/14.06.00/2021, THL/688/14.06.00/2021, THL/1284/14.06.00/2021, THL/1965/14.06.00/2021, THL/5546/14.02.00/2020, THL/2658/14.06.00/2021, THL/4235/14.06.00/202, Statistics Finland (permit numbers: TK-53-1041-17 and TK/143/07.03.00/2020 (earlier TK-53-90-20) TK/1735/07.03.00/2021, TK/3112/07.03.00/2021) and Finnish Registry for Kidney Diseases permission/extract from the meeting minutes on 4th July 2019.

The Biobank Access Decisions for FinnGen samples and data utilized in FinnGen Data Freeze 9 include: THL Biobank BB2017_55, BB2017_111, BB2018_19, BB_2018_34, BB_2018_67, BB2018_71, BB2019_7, BB2019_8, BB2019_26, BB2020_1, Finnish Red Cross Blood Service Biobank 7.12.2017, Helsinki Biobank HUS/359/2017, HUS/248/2020, Auria Biobank AB17-5154 and amendment #1 (August 17 2020), AB20-5926 and amendment #1 (April 23 2020) and it’s modification (Sep 22 2021), Biobank Borealis of Northern Finland_2017_1013, Biobank of Eastern Finland 1186/2018 and amendment 22 §/2020, Finnish Clinical Biobank Tampere MH0004 and amendments (21.02.2020 & 06.10.2020), Central Finland Biobank 1-2017, and Terveystalo Biobank STB 2018001 and amendment 25th Aug 2020.

Genes & Health is/has recently been core-funded by Wellcome (WT102627, WT210561), the Medical Research Council (UK) (M009017, MR/X009777/1, MR/X009920/1), Higher Education Funding Council for England Catalyst, Barts Charity (845/1796), Health Data Research UK (for London substantive site), and research delivery support from the NHS National Institute for Health Research Clinical Research Network (North Thames). Genes & Health is/has recently been funded by Alnylam Pharmaceuticals, Genomics PLC; and a Life Sciences Industry Consortium of Astra Zeneca PLC, Bristol-Myers Squibb Company, GlaxoSmithKline Research and Development Limited, Maze Therapeutics Inc, Merck Sharp & Dohme LLC, Novo Nordisk A/S, Pfizer Inc, Takeda Development Centre Americas Inc.

Ethics approval for Genes & Health was obtained from the London South East Research Ethics Committee (IRAS 146051). We thank Social Action for Health, Centre of The Cell, members of our Community Advisory Group, and staff who have recruited and collected data from volunteers. We thank the NIHR National Biosample Centre (UK Biocentre), the Social Genetic & Developmental Psychiatry Centre (King’s College London), Wellcome Sanger Institute, and Broad Institute for sample processing, genotyping, sequencing and variant annotation. We thank: Barts Health NHS Trust, NHS Clinical Commissioning Groups (City and Hackney, Waltham Forest, Tower Hamlets, Newham, Redbridge, Havering, Barking and Dagenham), East London NHS Foundation Trust, Bradford Teaching Hospitals NHS Foundation Trust, Public Health England (especially David Wyllie), Discovery Data Service/Endeavour Health Charitable Trust (especially David Stables), Voror Health Technologies Ltd (especially Sophie Don), NHS England (for what was NHS Digital) - for GDPR-compliant data sharing backed by individual written informed consent. Most of all we thank all of the volunteers participating in Genes & Health.

We would like to thank Vishnu Raj, Giulia Brunelli, Zhiyu Wang, Ying Wang, Kimmo Mattila, Anne Carson, Julius Anckar, Uwe Ohler and all members of INTERVENE who gave feedback, answered questions, or otherwise facilitated the development of this project.

## Supplementary Results

### CV versus automatic tuning

Out of 14 traits, the score generated by automatic tuning was selected via CV six times for PRScs, four times for lassosum and one time for LDpred2 (of note, PRScs explores only a small grid of hyperparameters). Consequentially, we could only test for significant differences (H_0_: β_CV*_-β_auto*_=0) in a subset of traits (and these cases were not considered when calculating the medians reported in the main text. We use β_CV*_ and β_auto*_ to denote the meta-analyzed effect sizes for CV-tuned or automatically tuned scores of the same method, respectively. For EUR target data, hyperparameter tuning often significantly increased PGS effect sizes (for 10/13 traits for LDpred2, 9/10 for lassosum, 10/14 for MegaPRS and 2/8 for PRScs) and rarely significantly decreased effect sizes, specifically for RA (PRScs and MegaPRS, driven by FinnGen), and CKD (PRScs) (Supplementary Figure 12, FWER <= 0.05, two-sided z-test). For SAS target data, we observed fewer significant performance increases through CV-hyperparameter tuning: 6/13 for LDpred2, 2/9 for lassosum, 2/13 for pT+clump and 1/13 for MegaPRS. Effect-sizes never significantly increased for PRScs, and it was the only method that saw a significant reduction in effect size for this ancestry group (BMI) (Supplementary Figure 13, FWER <= 0.05, two-sided z-test).

## Supplementary Tables

Detailed column descriptions are provided in the supplementary table file worksheets.

**Supplementary Table 1: PGS effect sizes for binary traits within biobanks and ancestries (EUR, SAS)**

**Supplementary Table 2: PGS effect sizes for continuous traits within biobanks and ancestries (EUR, SAS)**

**Supplementary Table 3: Pairwise comparisons of all scores within biobanks and ancestries (EUR, SAS)**

**Supplementary Table 4: Meta-analyzed effect size estimates for all methods and tuning types (CV, auto) across traits and ancestries (EUR, SAS)**

**Supplementary Table 5: Pairwise comparisons between meta-analyzed effect size estimates for binary traits for all methods and tuning types (CV, auto) across traits and ancestries (EUR, SAS)**

**Supplementary Table 6: Pairwise comparisons between meta-analyzed effect size estimates for continuous traits for all methods and tuning types (CV, auto) across traits and ancestries (EUR, SAS)**

**Supplementary Table 7: Relative r-squared of methods compared to the ensemble PGS, stratified by tuning type (CV, auto), trait (binary, continuous) and ancestry (EUR, SAS)**

**Supplementary Table 8: Average and median relative effect sizes of scores tuned with cross-validation against those tuned using automatic settings across methods and ancestries (EUR/SAS)**

**Supplementary Table 9: 3-level meta-analytical random effects model results (EUR)**

**Supplementary Table 10: 3-level meta-analytical random effects model results (EUR) without excluding SBayesR-auto for arthritis, BMI, gout and T1D**

**Supplementary Table 11: 3-level meta-analytical random effects model results (SAS)**

**Supplementary Table 12: 3-level meta-analytical random effects model results (SAS) without excluding SBayesR-auto for BMI, gout and urate**

**Supplementary Table 13: Binary Endpoint definitions based on ICD-10/9 codes**

**Supplementary Table 14: PGS method software versions**

**Supplementary Table 15: PGS catalog score identifiers**

## Supplementary Figures

Supplementary Figures 1-5 are provided as separate files.

**Supplementary Figure 1:**
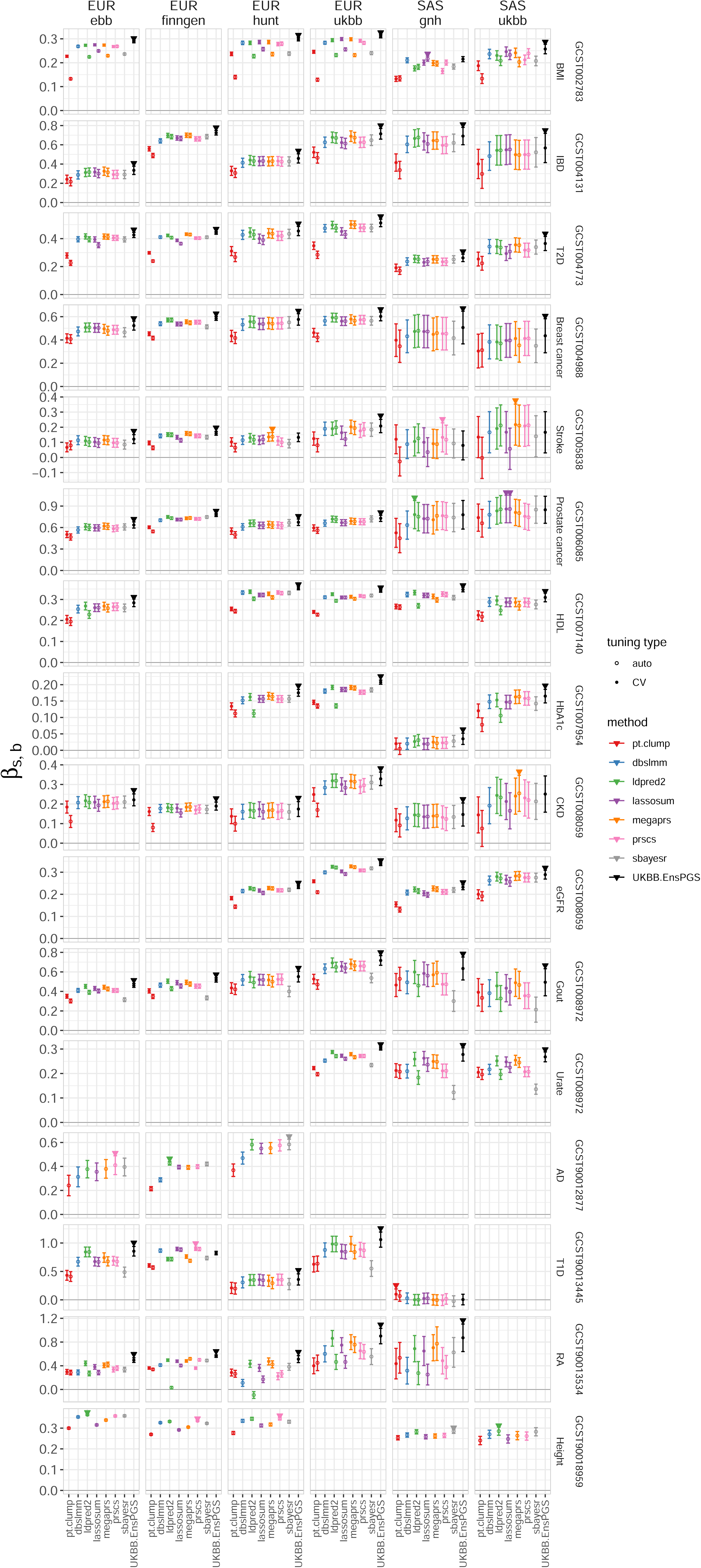
PGS β coefficients within biobanks and ancestries (EUR, SAS) for binary and continuous traits. β coefficients, i.e, the change in log odds ratio (binary traits) or trait standard deviations (continuous traits) per standard deviation of the PGS, across biobanks, ancestries, and phenotypes is shown (β_s,b_). Colors represent PGS methods, and method tuning types (CV, auto) are indicated by different markers. The score with the largest effect size within each biobank-ancestry group is marked with a triangle. Error bars denote 95% confidence intervals. Disease labels and GWAS catalog identifiers are given to the right. The data in this plot are browsable online and available in Supplementary Table 1-2.

**Supplementary Figure 2:**
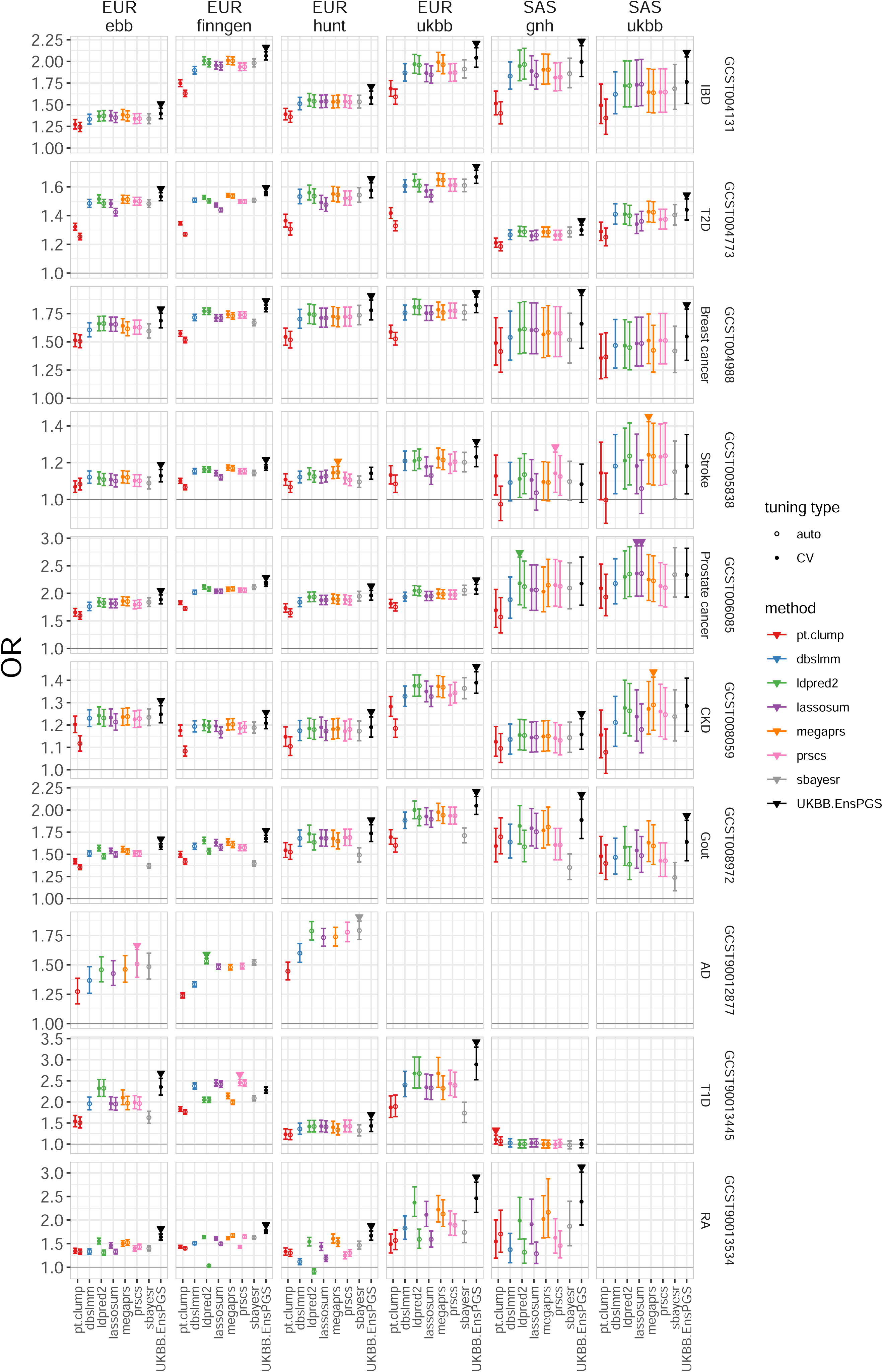
PGS odds ratios per PGS standard deviation within biobanks and ancestries (EUR, SAS) (binary traits) The change in odds ratio per standard deviation of the PGS (OR), across biobanks, ancestries, and phenotypes is shown. Colors represent PGS methods, and method tuning types (CV, auto) are indicated by different markers. The score with the largest effect size within each biobank-ancestry group is marked with a triangle. Error bars denote 95% confidence intervals. Disease labels and GWAS catalog identifiers are given to the right. The data in this plot are browsable online and available in Supplementary Table 1.

**Supplementary Figure 3:**
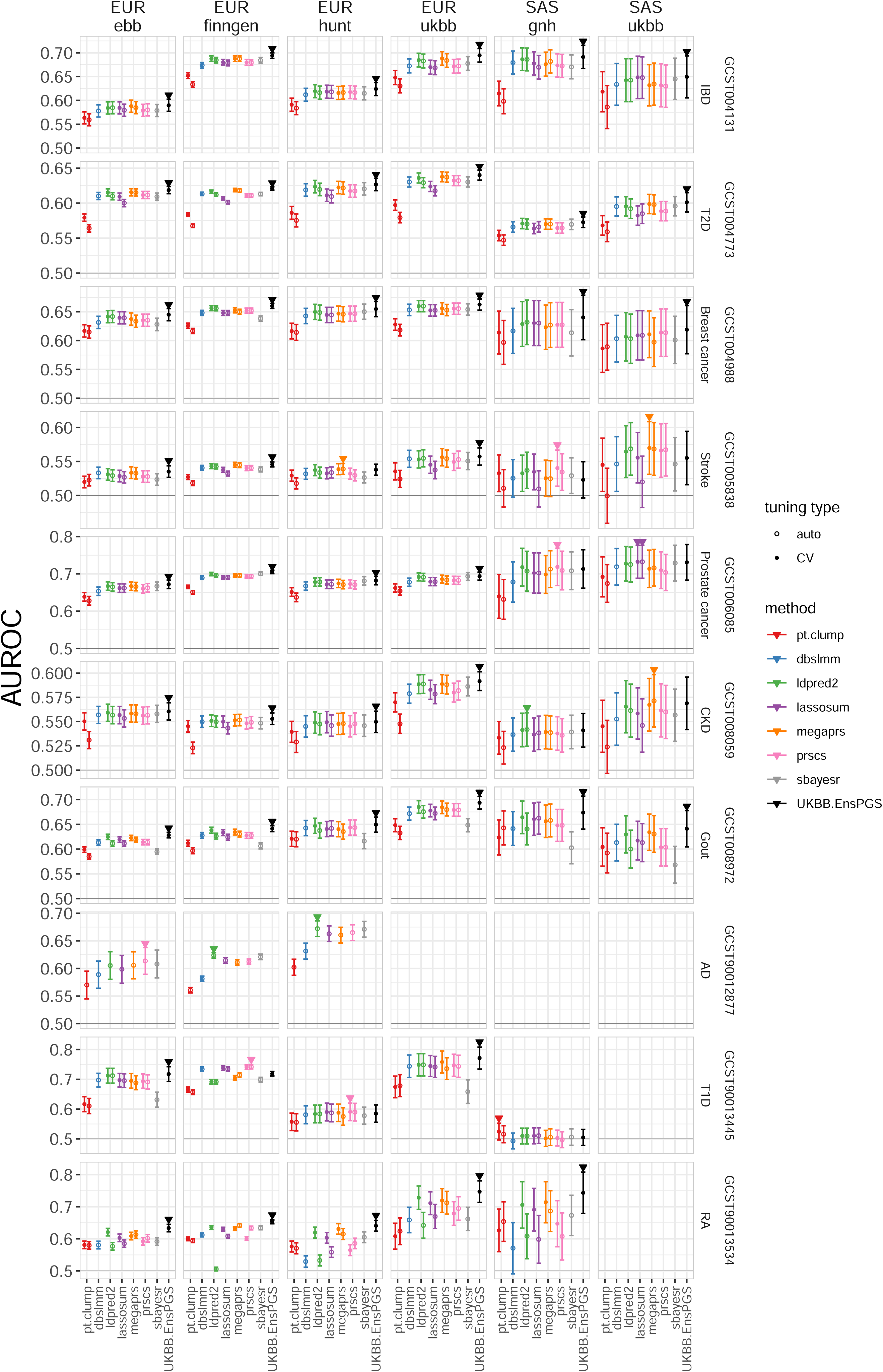
PGS area under the receiver operator characteristic curve within biobanks and ancestries (EUR, SAS) (binary traits) The area under the receiver operator characteristic curve (AUROC) of the PGS, across biobanks, ancestries, and phenotypes is shown. Colors represent PGS methods, and method tuning types (CV, auto) are indicated by different markers. The score with the largest effect size within each biobank-ancestry group is marked with a triangle. Error bars denote 95% DeLong confidence intervals (Methods). Disease labels and GWAS catalog identifiers are given to the right. The data in this plot are browsable online and available in Supplementary Table 1.

**Supplementary Figure 4:**
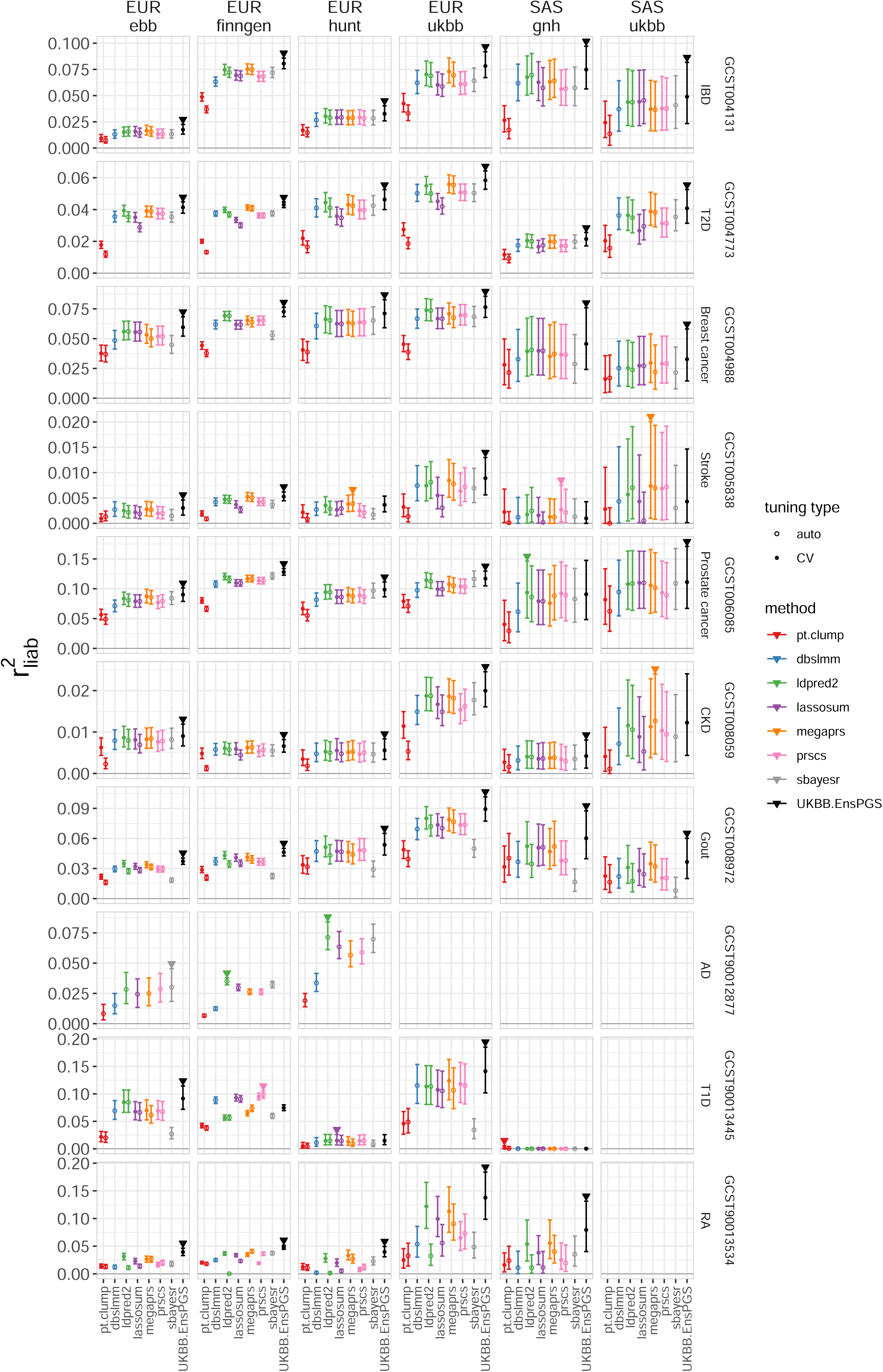
PGS variance explained on the liability scale within biobanks and ancestries (EUR, SAS) (binary traits) The variance explained by the PGS on the liability scale (r^2^_liab_, Methods), across biobanks, ancestries, and phenotypes is shown. Colors represent PGS methods, and method tuning types (CV, auto) are indicated by different markers. The score with the largest effect size within each biobank-ancestry group is marked with a triangle. Error bars denote 95% bootstrap-based confidence intervals (Methods). Disease labels and GWAS catalog identifiers are given to the right. The data in this plot are browsable online and available in Supplementary Table 1.

**Supplementary Figure 5:**
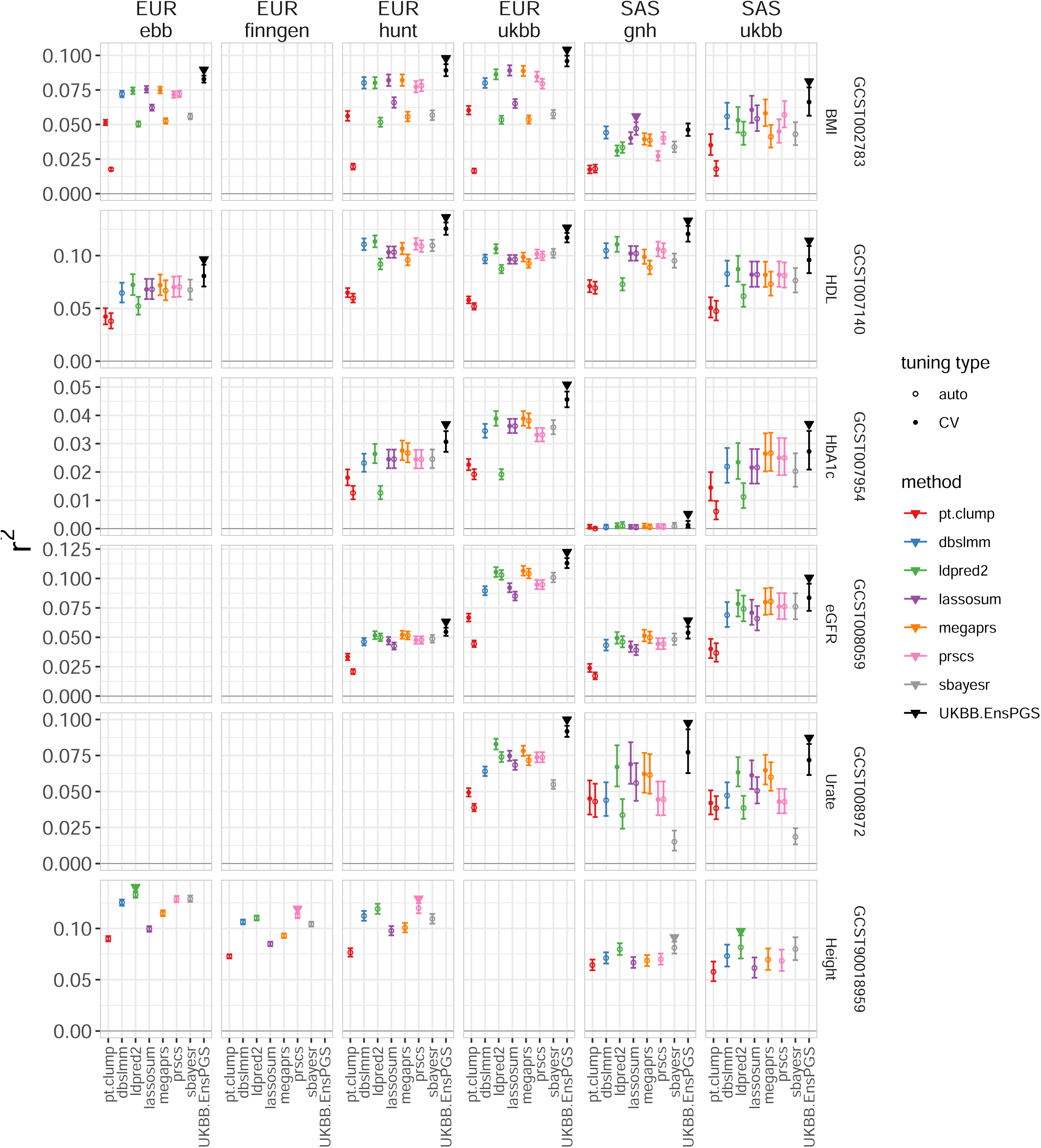
PGS variance explained on the observed scale within biobanks and ancestries (EUR, SAS) (continuous traits) The variance explained by the PGS on the observed scale (r^2^), across biobanks, ancestries, and phenotypes is shown. Colors represent PGS methods, and method tuning types (CV, auto) are indicated by different markers. The score with the largest effect size within each biobank-ancestry group is marked with a triangle. Error bars denote 95% confidence intervals. Disease labels and GWAS catalog identifiers are given to the right. The data in this plot are browsable online and available in Supplementary Table 2.

**Supplementary Figure 6:**
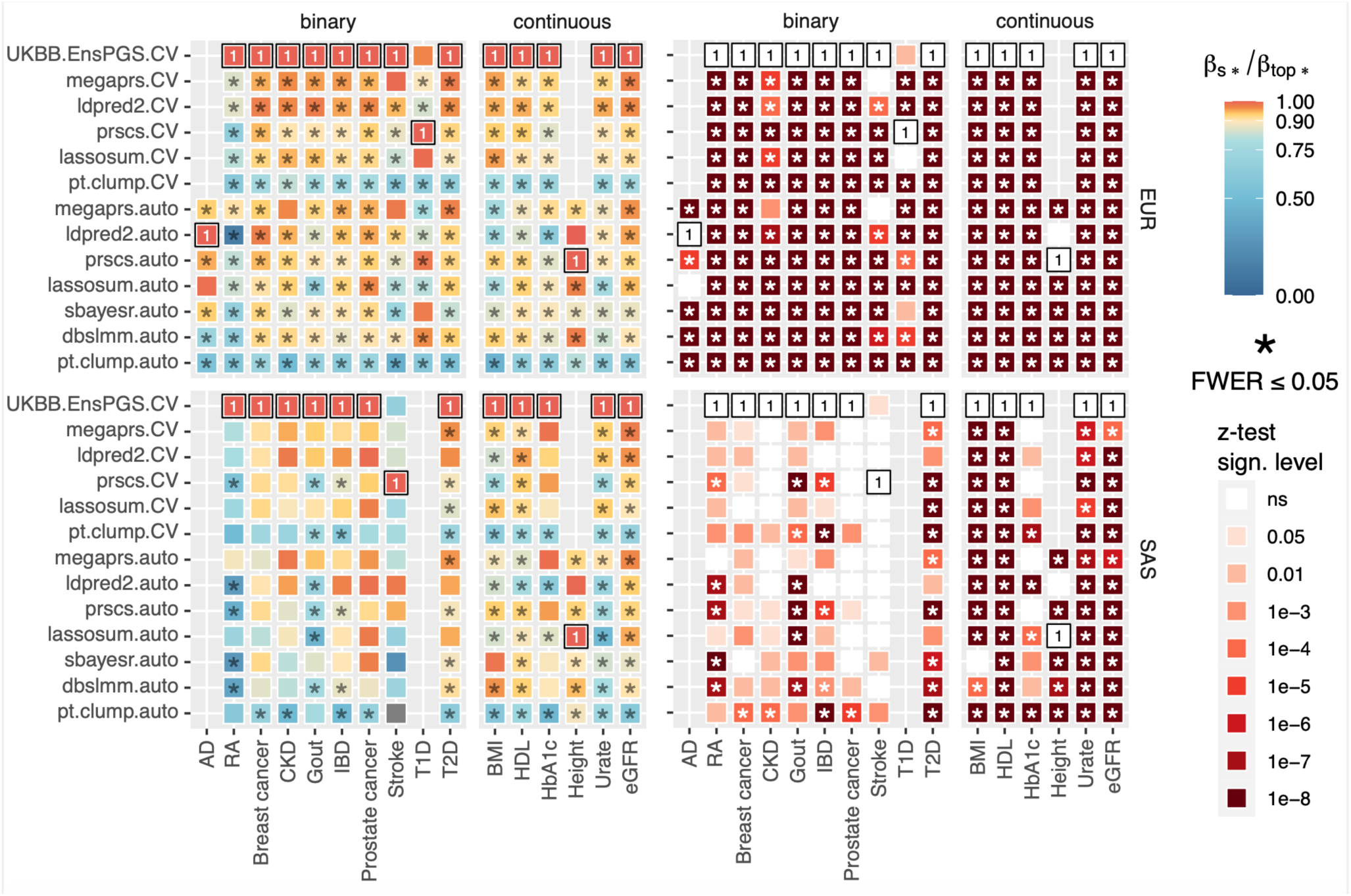
Relative meta-analyzed PGS effect sizes across 16 traits. For the all traits (x-axis) we show heatmaps of meta-analyzed β-coefficients relative to the highest (β_s*_/β_top*_, left) as well as the corresponding significance thresholds for the two-sided z-test (H_0_: β_s*_-β_top*_= 0, right) stratified by ancestry (EUR, SAS), method, and tuning type (CV, auto). The score with the largest effect size for each trait (β_top*_) is marked with a “1” and black box. Differences significant at FWER <= 0.05 are marked with asterisks (*), accounting for all 351 tests. Pairwise comparisons between all PGS are reported in Supplementary Table 5-6.

**Supplementary Figure 7:**
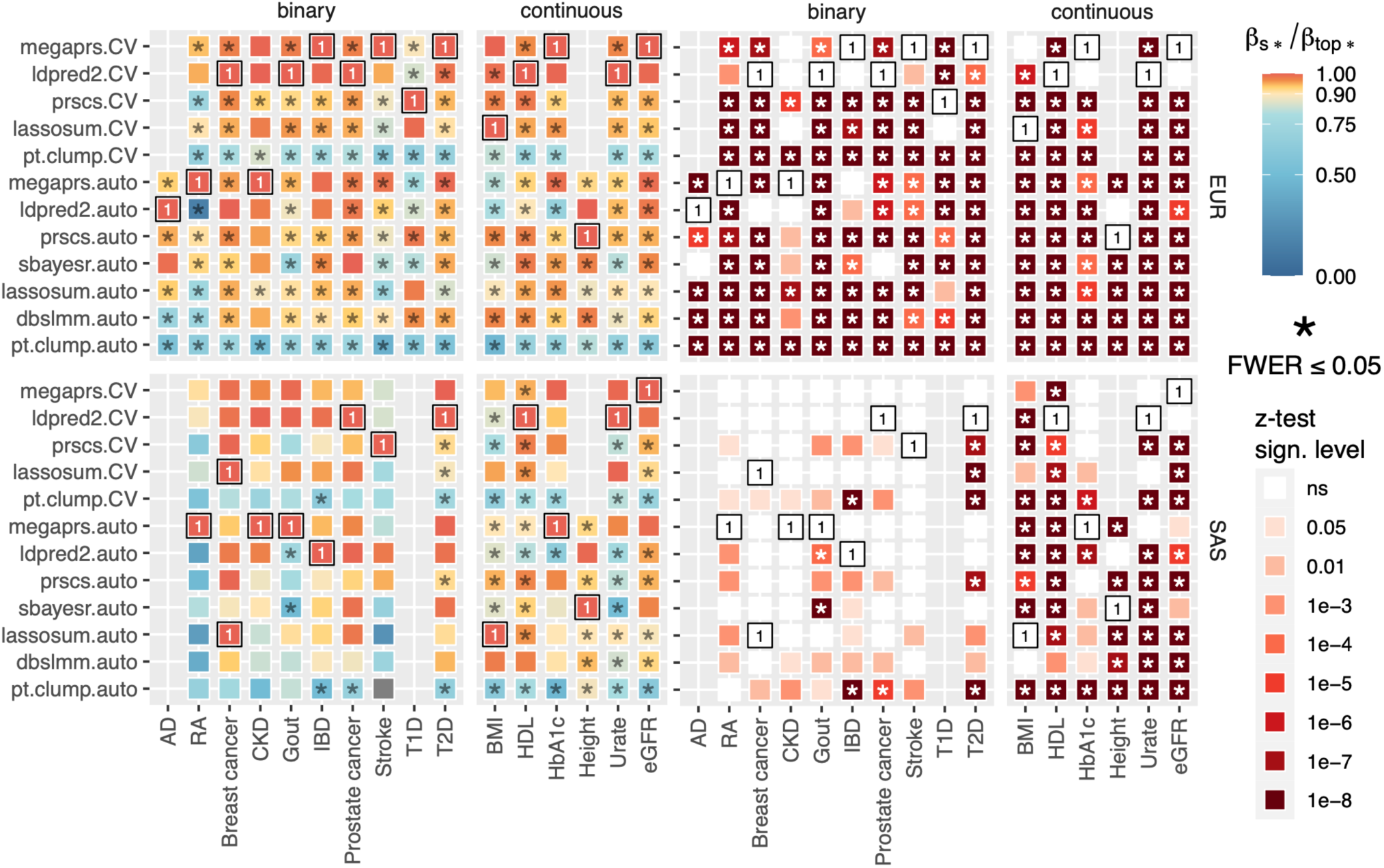
Relative meta-analyzed PGS effect sizes across 16 traits excluding the ensemble PGS. For the all traits (x-axis) we show heatmaps of meta-analyzed β-coefficients relative to the highest (excluding the ensemble PGS, β_s*_/β_top*_, left) as well as the corresponding significance thresholds for the two-sided z-test (H_0_: β_s*_-β_top*_= 0, right) stratified by ancestry (EUR, SAS), method, and tuning type (CV, auto). The score with the largest effect size for each trait (β_top*_) is marked with a “1” and black box. Differences significant at FWER <= 0.05 are marked with asterisks (*), accounting for all 351 tests. Pairwise comparisons between all PGS are reported in Supplementary Table 5-6.

**Supplementary Figure 8:**
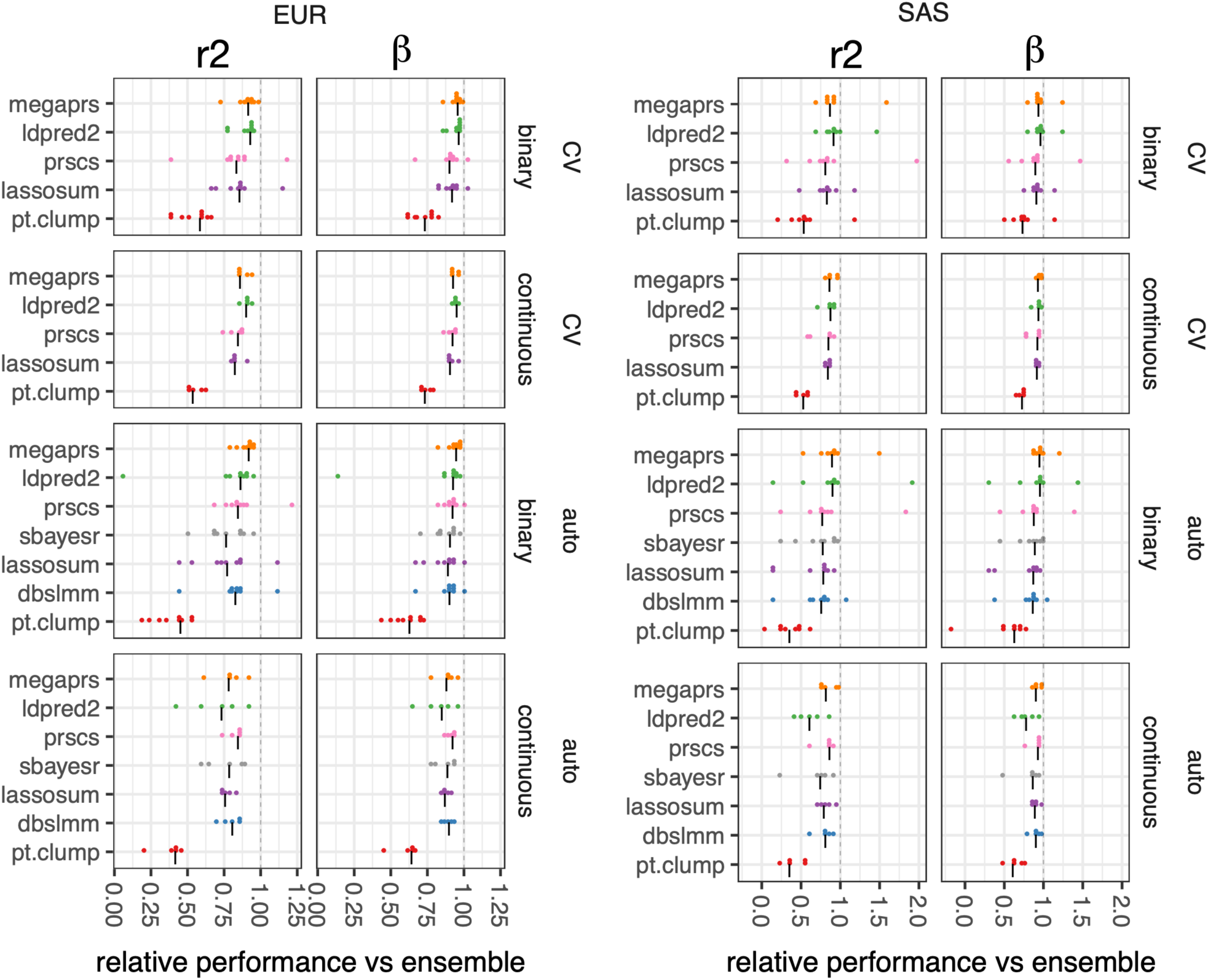
Meta-analyzed performance of methods relative to the ensemble PGS stratified by type of tuning (CV/auto), trait (binary/continuous) and ancestry (EUR, SAS). Dot plots showing, for each method (y-axis), the meta-analyzed variance explained (r2, on the liability scale for binary straits) or the meta-analyzed β-coefficients relative to the ensemble PGS across traits (every dot represents a single trait), stratified by tuning type (auto/CV) and type of trait (binary/continuous). Horizontal black lines “|” denote the median values. This Figure corresponds to the medians and averages reported in Table 3.

**Supplementary Figure 9:**
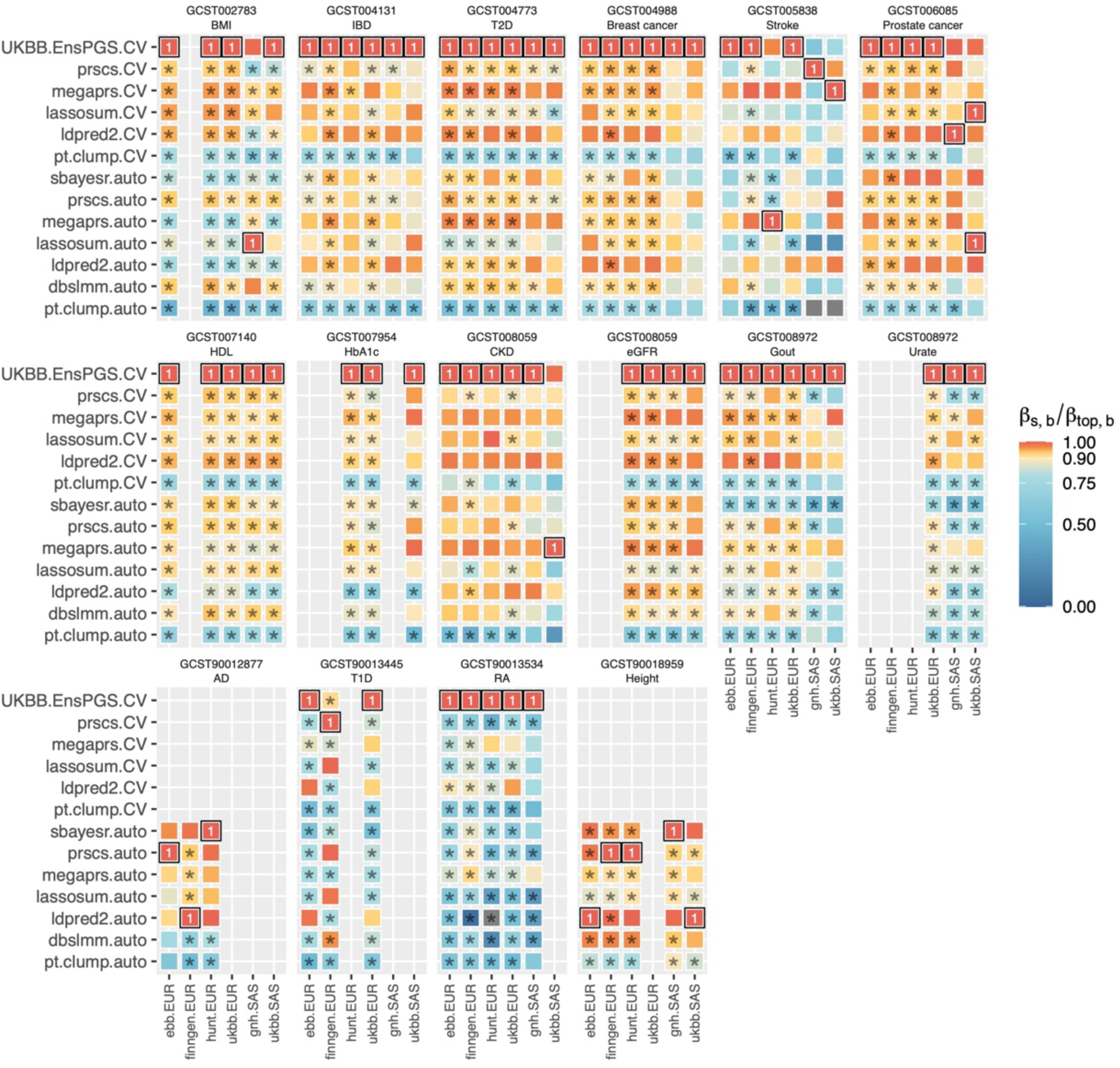
Relative PGS effect sizes (β_s,b_/β_top,b_) across 16 traits within biobanks and ancestries. We show heatmaps of PGS effect sizes (β-coefficients) relative to the highest (β_s,b_/β_top,b_) within biobanks (ebb, finngen, hunt, ukbb, gnh) and ancestries (EUR, SAS) across traits (subpanels). The score with the largest effect size for each trait within each ancestry and biobank (β_top,b_) is marked with a “1” and black box. Differences significant at FWER <= 0.05 are marked with asterisks (*), accounting for all 915 tests (H_0_: β_s,b_-β_top,b_= 0, two-sided z-test). All pairwise comparisons are reported in Supplementary Table 3.

**Supplementary Figure 10:**
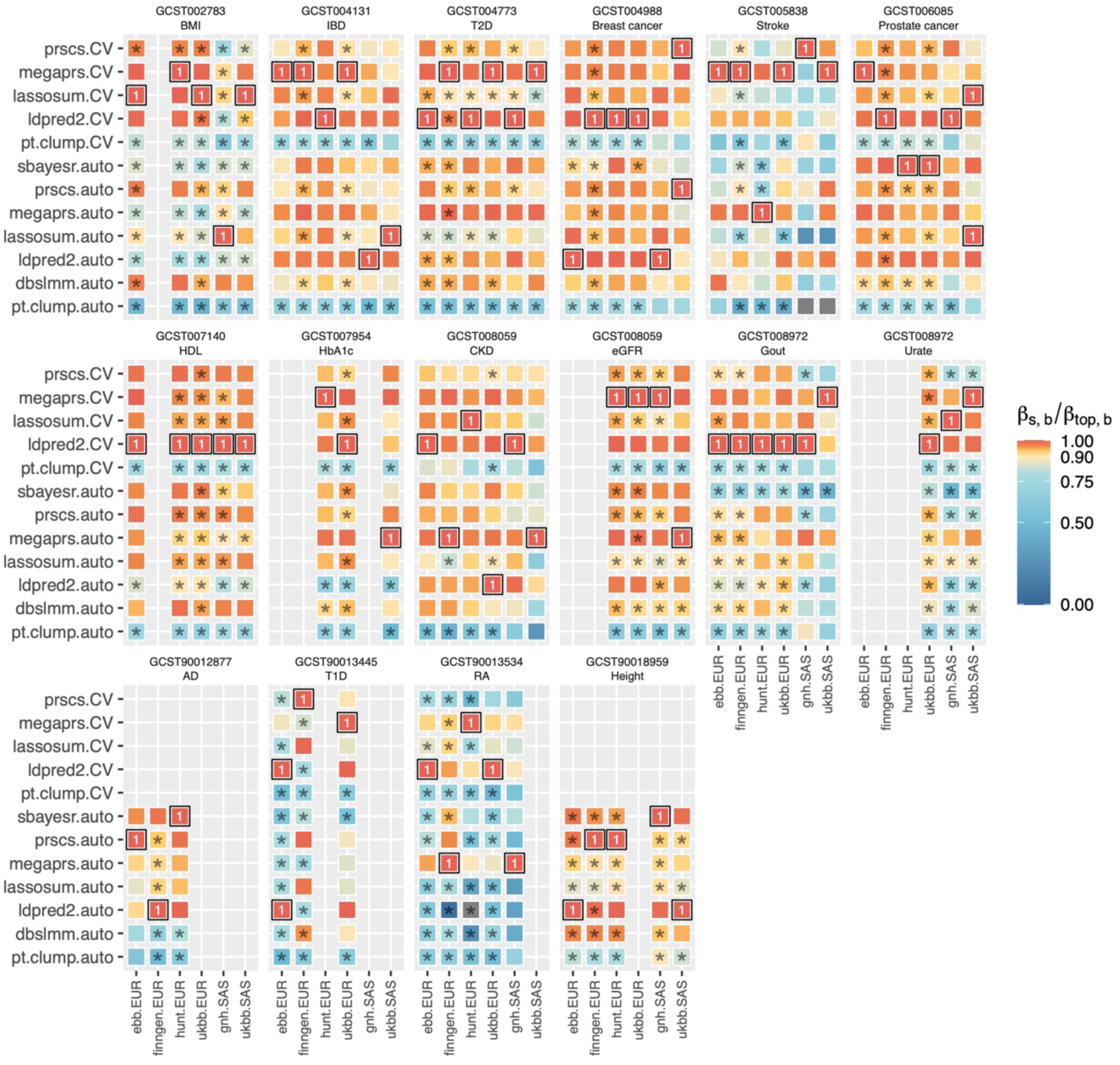
Relative PGS effect sizes (β_s,b_/β_top,b_) across 16 traits within biobanks and ancestries excluding the ensemble PGS. For all traits (x-axis) we show heatmaps of PGS effect sizes (β-coefficients) relative to the highest (β_s,b_/β_top,b_) within biobanks (ebb, finngen, hunt, ukbb, gnh) and ancestries (EUR, SAS) across traits (subpanels). The score with the largest effect size for each trait within each ancestry and biobank (β_top,b_) is marked with a “1” and black box (excluding the ensemble PGS). Differences significant at FWER <= 0.05 are marked with asterisks (*), accounting for all 915 tests (H_0_: β_s,b_-β_top,b_= 0= 0, two-sided z-test). All pairwise comparisons are reported in Supplementary Table 3.

**Supplementary Figure 11:**
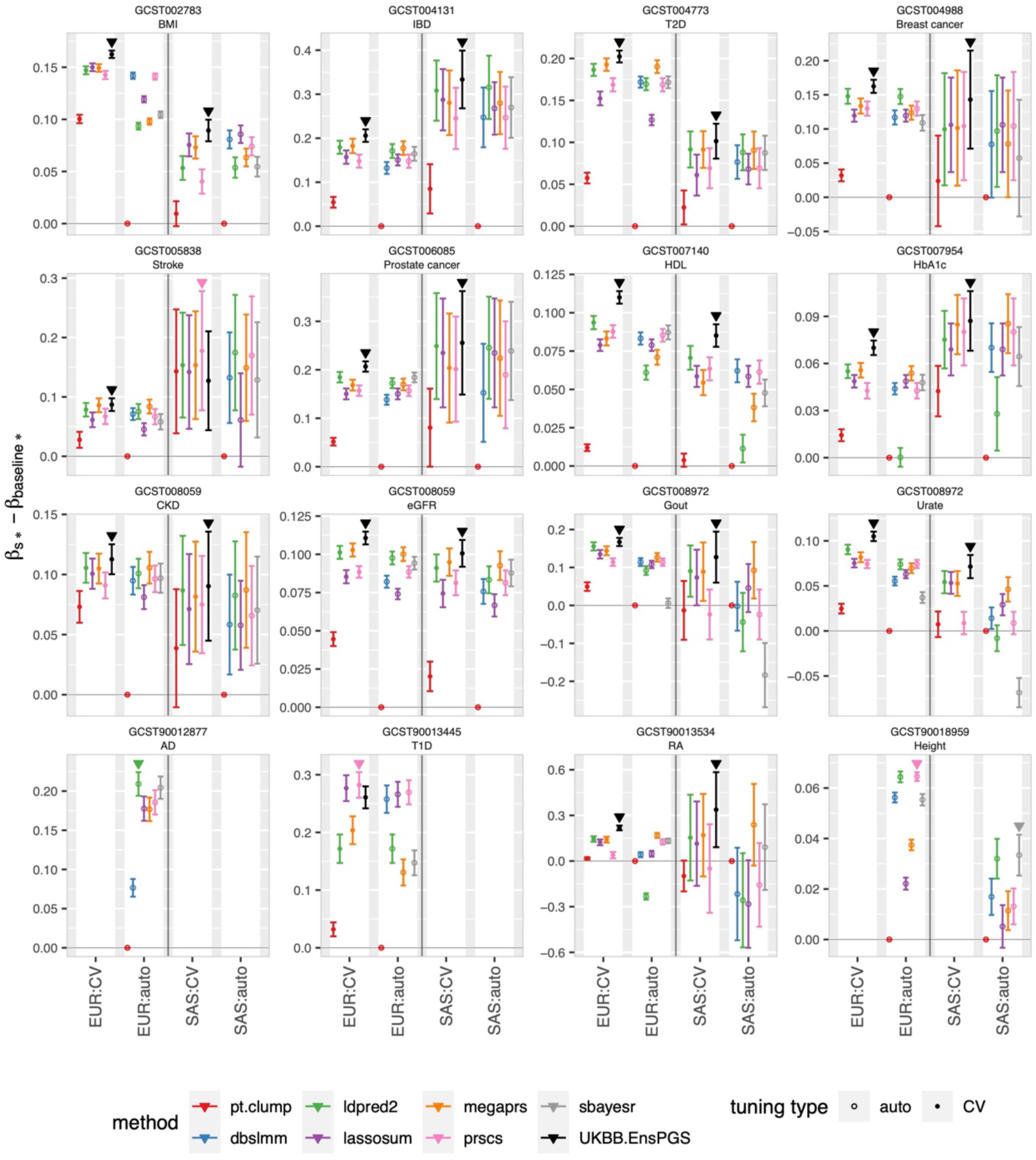
Meta-analyzed PGS performance relative to the baseline method pT+clump that uses highly significant variants only. Across all traits, we show meta-analyzed β-coefficient-differences compared to the baseline pT+clump-auto (β_s*_- β_basline*_) that uses only highly significant (p<1e-8) SNPs (y-axis). Effect sizes are stratified by method, tuning type (CV/auto) and ancestry (EUR/SAS). The score with the highest effect size within ancestries and traits is marked with a triangle. Error bars denote 95% confidence intervals. CV-tuning could not be performed for AD and height due to GWAS sample overlap with the UKBB EUR-ancestry data. All pairwise comparisons are reported in Supplementary Table 5-6.

**Supplementary Figure 12:**
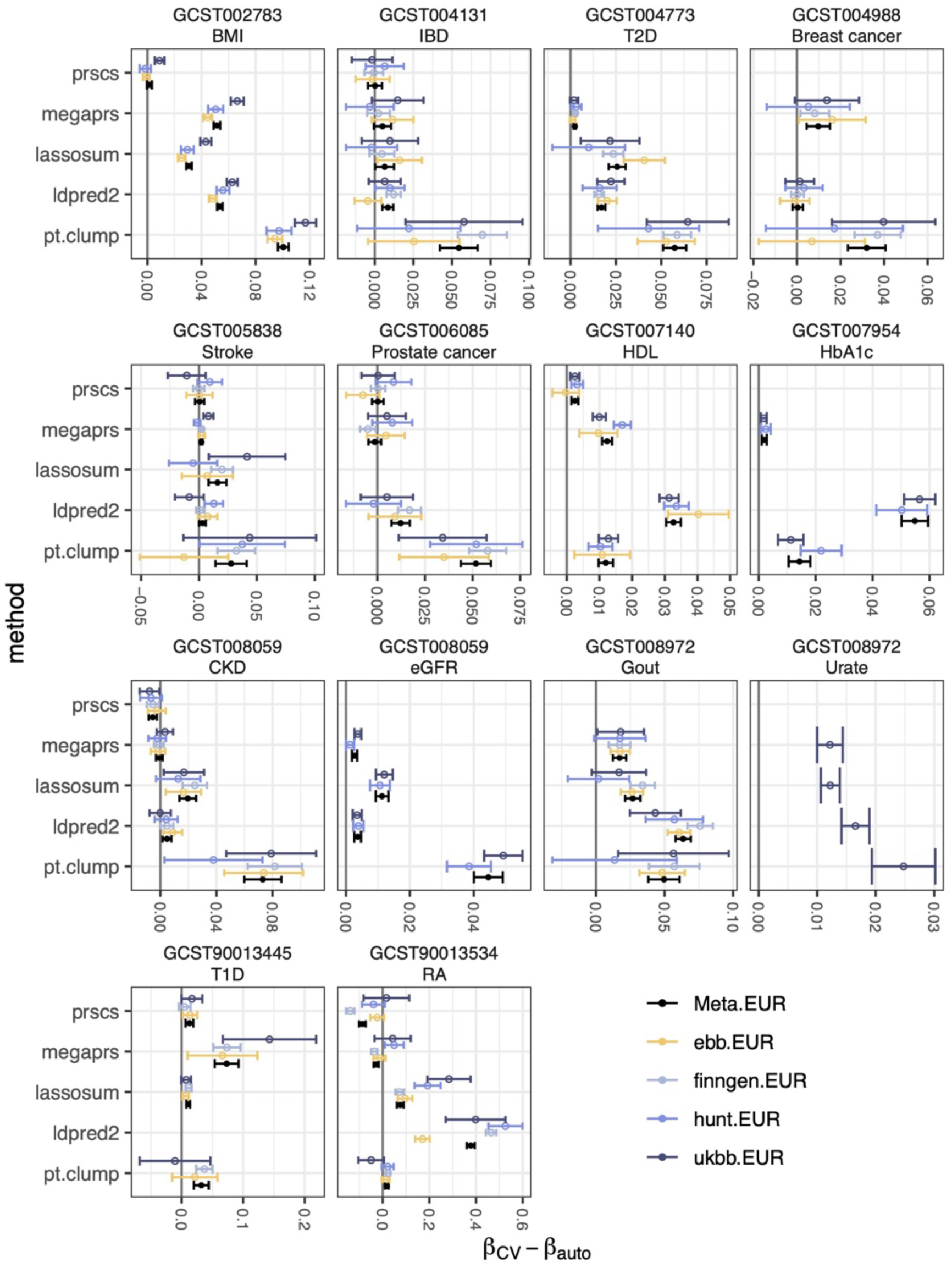
Comparison of CV- and automatic tuning in EUR target data. Effect-size differences between CV- and automatically tuned PGS for five methods (β_CV_- β_auto_) across traits, including 95% confidence intervals. Both values within biobanks (ebb, finngen, hunt, ukbb) and the meta-analyzed estimates (Meta) are shown. If the auto-score was selected via CV, no differences are shown.

**Supplementary Figure 13:**
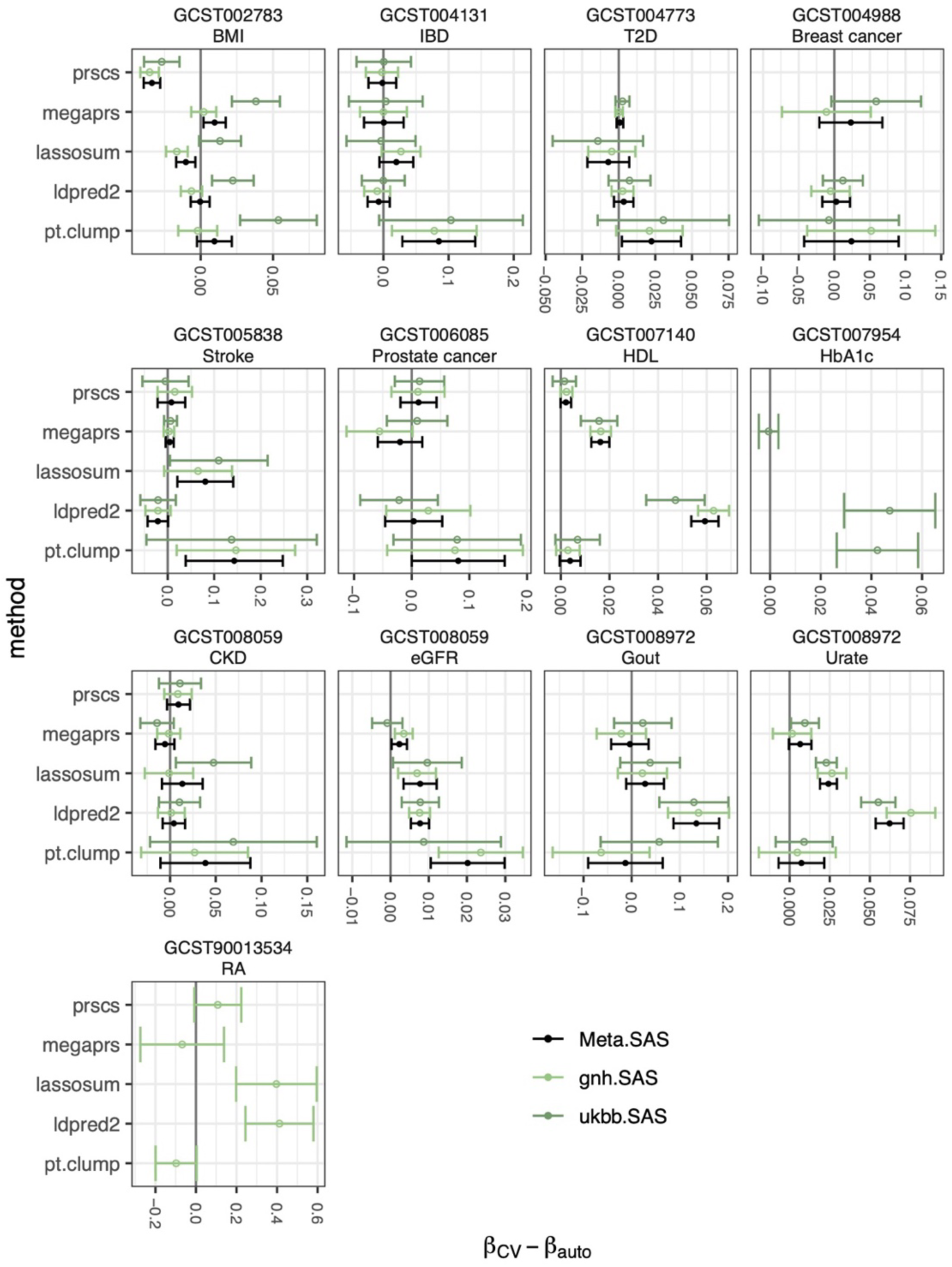
Comparison of CV- and automatic tuning in SAS target data. Effect-size differences between CV- and automatically tuned PGS for five methods (β_CV-_ β_auto_) across traits, including 95% confidence intervals. Both values within biobanks (gnh, ukbb) and the meta-analyzed estimates (Meta) are shown. If the auto-score was selected via CV, no differences are shown.

**Supplementary Figure 14:**
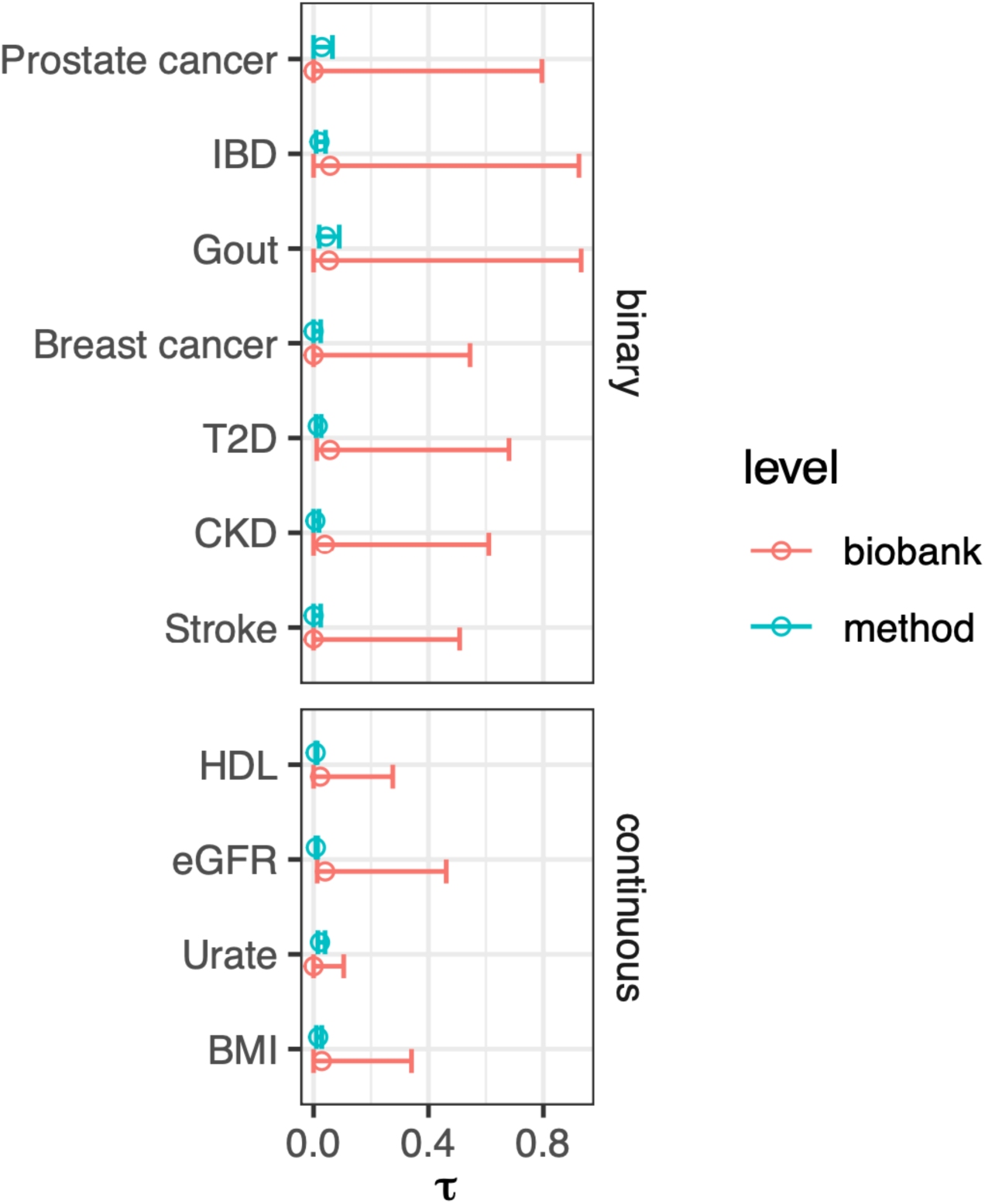
τ_biobank_ and τ_method_ estimated in SAS ancestry target data. Estimates for τ_biobank_ and τ_method_ (x-axis, Methods) for traits available in both the UKBB and GNH SAS ancestry matched target data, including 95% likelihood-based confidence intervals. Endpoints are ordered by the average effect size of scores in that ancestry, from largest (top) to lowest (bottom). τ_biobank_ was not well identifiable in SAS ancestry target data (just two biobanks represented).

**Supplementary Figure 15:**
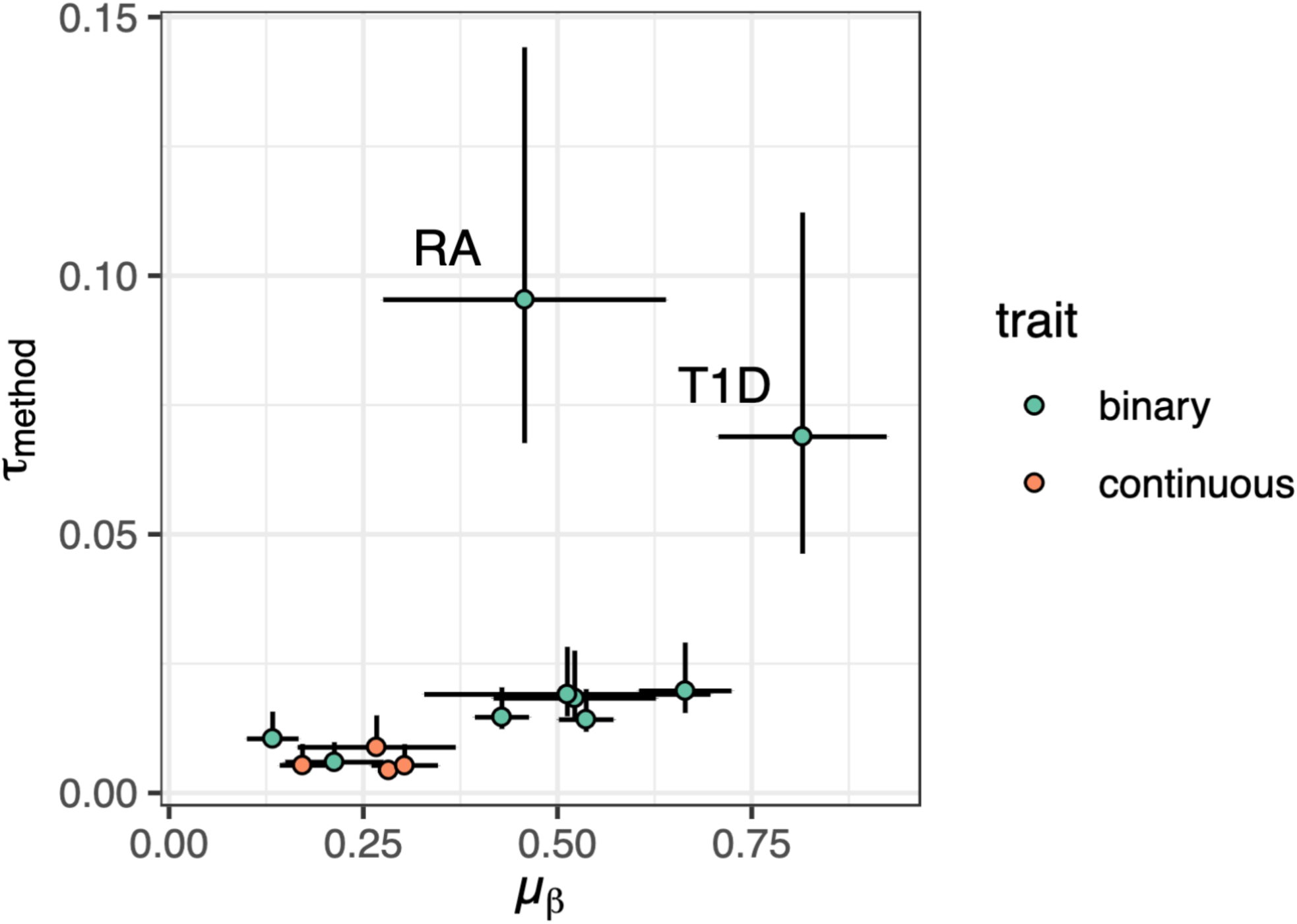
Scatterplot of μ_β_ vs τ_method_ in the 3-level meta-analytic random effects model (EUR) Scatterplot of the average meta-analyzed PGS β coefficients across methods and biobanks (μ_β_, x-axis) vs τ_method_ (y-axis) for the 13 traits available in at least two biobanks and EUR target data, colored by the type of trait (continuous/binary). Lines denote 95% confidence intervals. The two autoimmune diseases RA and T1D displayed the largest differences in effect size between methods within biobanks, as quantified by τ_method._ These scores also had the highest fraction of variance originating in the HLA region, as shown in Supplementary Figure 16.

**Supplementary Figure 16:**
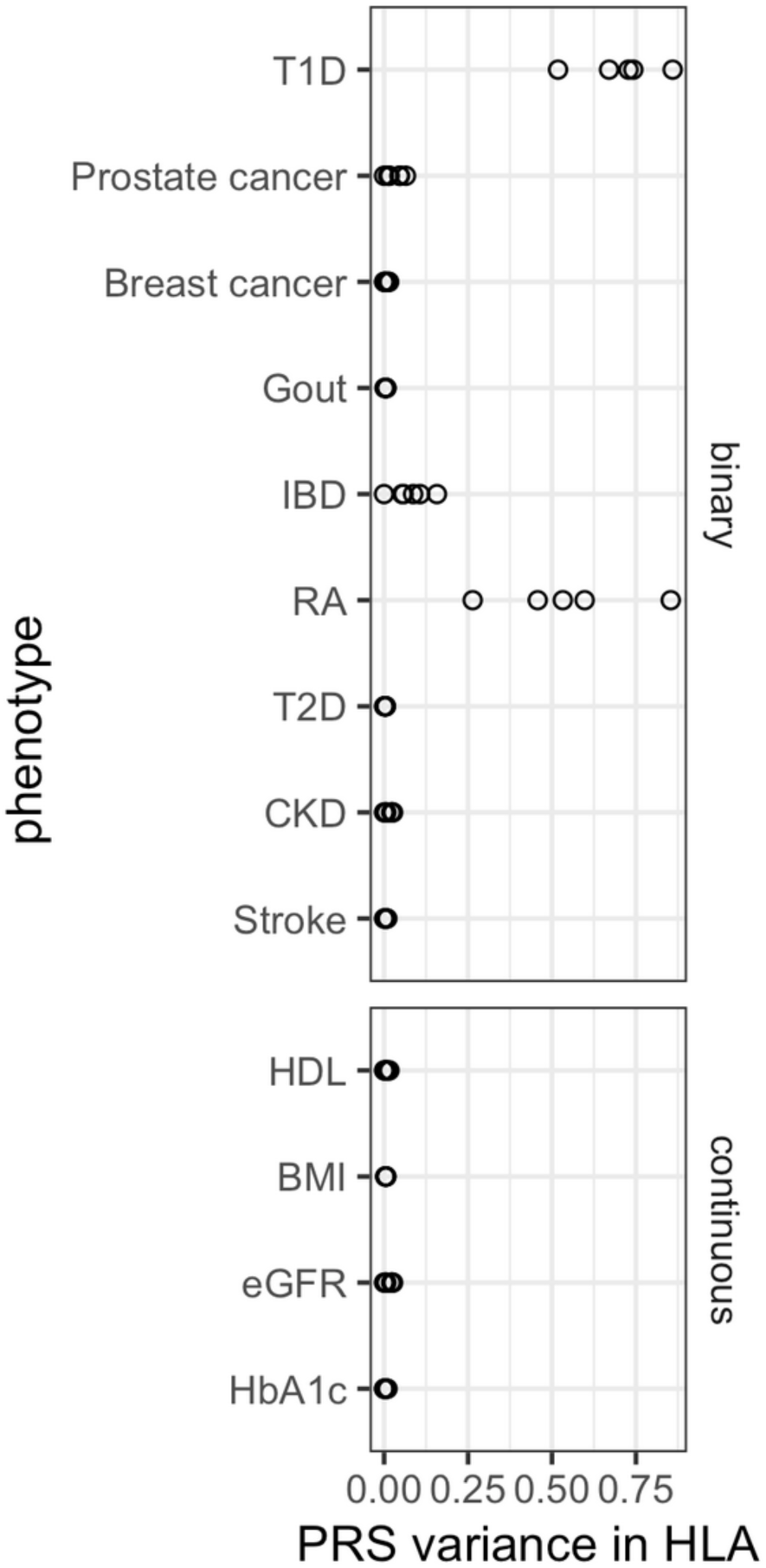
PGS variance originating in the HLA region for the scores used to estimate τ_biobank and_ τ_method_ in EUR ancestry target data. PGS variance originating in the HLA region divided by the total PGS variance (x-axis) calculated on the 1000 Genomes EUR reference data across traits (y-axis, Methods). Values were calculated for the 13 traits available in at least two biobanks and EUR target data for which we estimated τ_biobank_ and τ_method_. T1D and RA showed the highest fraction of PGS variance originating in the HLA region and had the most variable performance depending on the PGS method, as estimated by τ_method_ (Supplementary Figure 15).

## Supplementary Data

**forest_plots_eur.pdf**

Forest plots generated with the metafor package related to the 3-level meta-analysis.

