## Supplementary material for "Evaluation of polygenic scoring methods in five biobanks reveals greater variability between biobanks than between methods and highlights benefits of ensemble learning": forest plots (three level meta-analysis, forest_plots_eur.pdf)

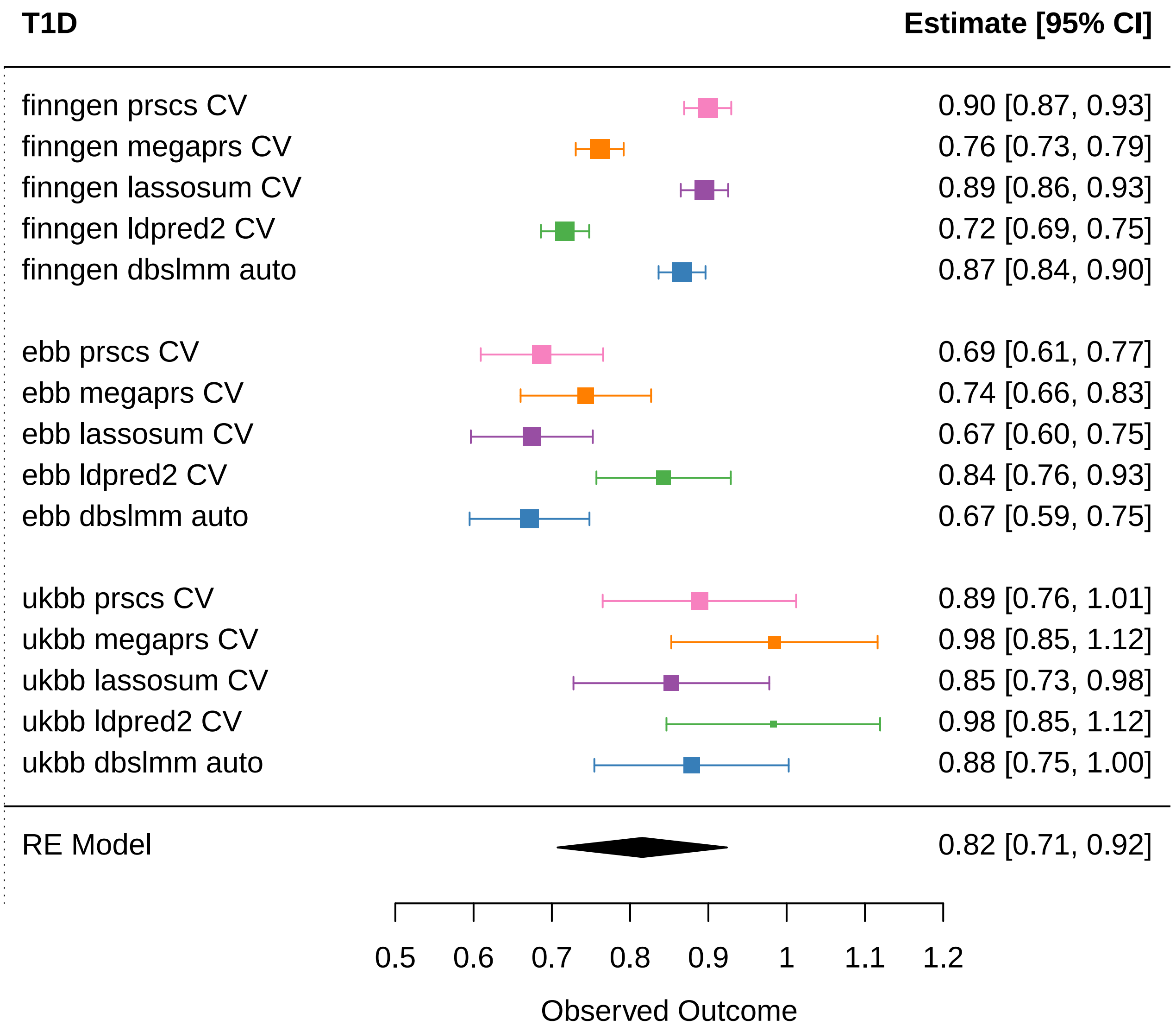
T1D
Estimate [95% CI]
finngen prscs CV
0.90 [0.87, 0.93]
finngen megaprs CV
0.76 [0.73, 0.79]
finngen lassosum CV
0.89 [0.86, 0.93]
finngen ldpred2 CV
0.72 [0.69, 0.75]
finngen dbslmm auto
0.87 [0.84, 0.90]
ebb prscs CV
0.69 [0.61, 0.77]
ebb megaprs CV
0.74 [0.66, 0.83]
ebb lassosum CV
0.67 [0.60, 0.75]
ebb ldpred2 CV
0.84 [0.76, 0.93]
ebb dbslmm auto
0.67 [0.59, 0.75]
ukbb prscs CV
0.89 [0.76, 1.01]
ukbb megaprs CV
0.98 [0.85, 1.12]
ukbb lassosum CV
0.85 [0.73, 0.98]
ukbb ldpred2 CV
0.98 [0.85, 1.12]
ukbb dbslmm auto
0.88 [0.75, 1.00]
RE Model
0.82 [0.71, 0.92]
0.5
0.6
0.7
0.8
0.9
1
1.1
1.2
Observed Outcome

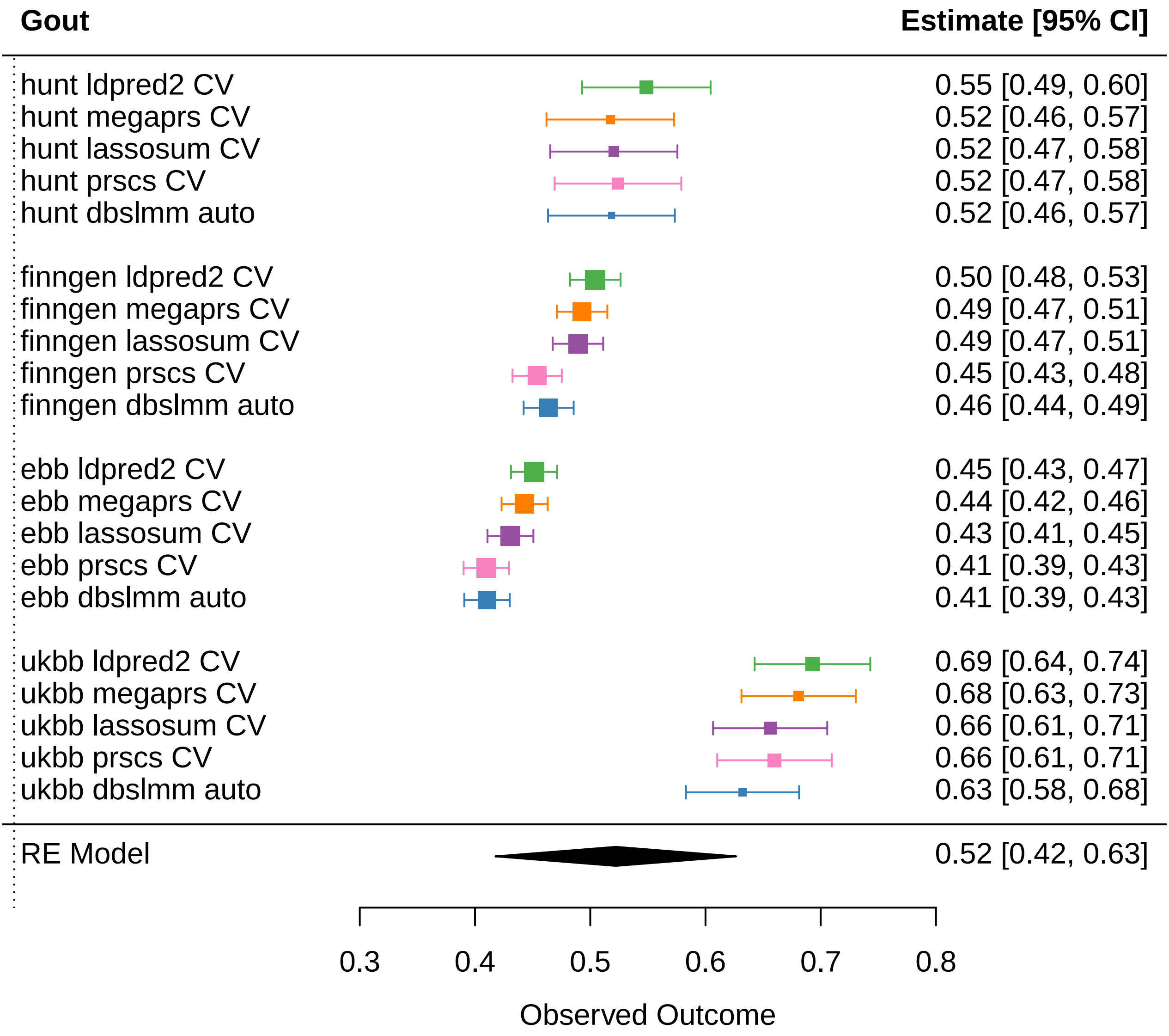
Gout
Estimate [95% CI]
hunt ldpred2 CV
hunt megaprs CV
hunt lassosum CV
hunt prscs CV
hunt dbslmm auto
finngen ldpred2 CV
finngen megaprs CV
finngen lassosum CV
finngen prscs CV
finngen dbslmm auto
ebb ldpred2 CV
ebb megaprs CV
ebb lassosum CV
ebb prscs CV
ebb dbslmm auto
ukbb ldpred2 CV
ukbb megaprs CV
ukbb lassosum CV
ukbb prscs CV
ukbb dbslmm auto
RE Model
0.55 [0.49, 0.60]
0.52 [0.46, 0.57]
0.52 [0.47, 0.58]
0.52 [0.47, 0.58]
0.52 [0.46, 0.57]
0.50 [0.48, 0.53]
0.49 [0.47, 0.51]
0.49 [0.47, 0.51]
0.45 [0.43, 0.48]
0.46 [0.44, 0.49]
0.45 [0.43, 0.47]
0.44 [0.42, 0.46]
0.43 [0.41, 0.45]
0.41 [0.39, 0.43]
0.41 [0.39, 0.43]
0.69 [0.64, 0.74]
0.68 [0.63, 0.73]
0.66 [0.61, 0.71]
0.66 [0.61, 0.71]
0.63 [0.58, 0.68]
0.52 [0.42, 0.63]
0.3
0.4
0.5
0.6
0.7
0.8
Observed Outcome

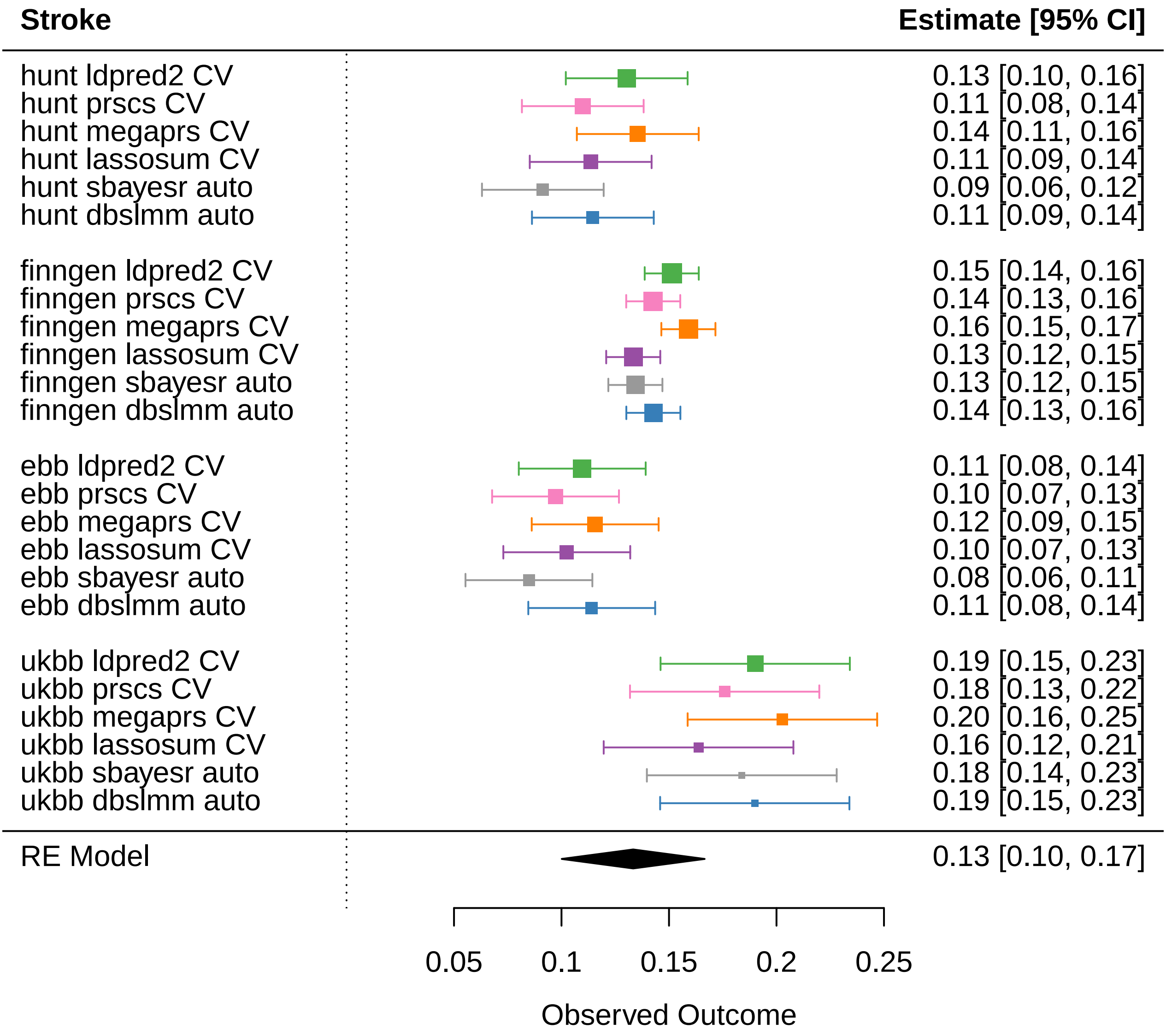
Stroke
Estimate [95% CI]
hunt ldpred2 CV 0.13 [0.10, 0.16]
hunt prscs CV 0.11 [0.08, 0.14]
hunt megaprs CV 0.14 [0.11, 0.16]
hunt lassosum CV 0.11 [0.09, 0.14]
hunt sbayesr auto 0.09 [0.06, 0.12]
hunt dbslmm auto 0.11 [0.09, 0.14]
finngen ldpred2 CV 0.15 [0.14, 0.16]
finngen prscs CV 0.14 [0.13, 0.16]
finngen megaprs CV 0.16 [0.15, 0.17]
finngen lassosum CV 0.13 [0.12, 0.15]
finngen sbayesr auto 0.13 [0.12, 0.15]
finngen dbslmm auto 0.14 [0.13, 0.16]
ebb ldpred2 CV 0.11 [0.08, 0.14]
ebb prscs CV 0.10 [0.07, 0.13]
ebb megaprs CV 0.12 [0.09, 0.15]
ebb lassosum CV 0.10 [0.07, 0.13]
ebb sbayesr auto 0.08 [0.06, 0.11]
ebb dbslmm auto 0.11 [0.08, 0.14]
ukbb ldpred2 CV 0.19 [0.15, 0.23]
ukbb prscs CV 0.18 [0.13, 0.22]
ukbb megaprs CV 0.20 [0.16, 0.25]
ukbb lassosum CV 0.16 [0.12, 0.21]
ukbb sbayesr auto 0.18 [0.14, 0.23]
ukbb dbslmm auto 0.19 [0.15, 0.23]
RE Model 0.13 [0.10, 0.17]
0.05
0.1
0.15
0.2
0.25
Observed Outcome

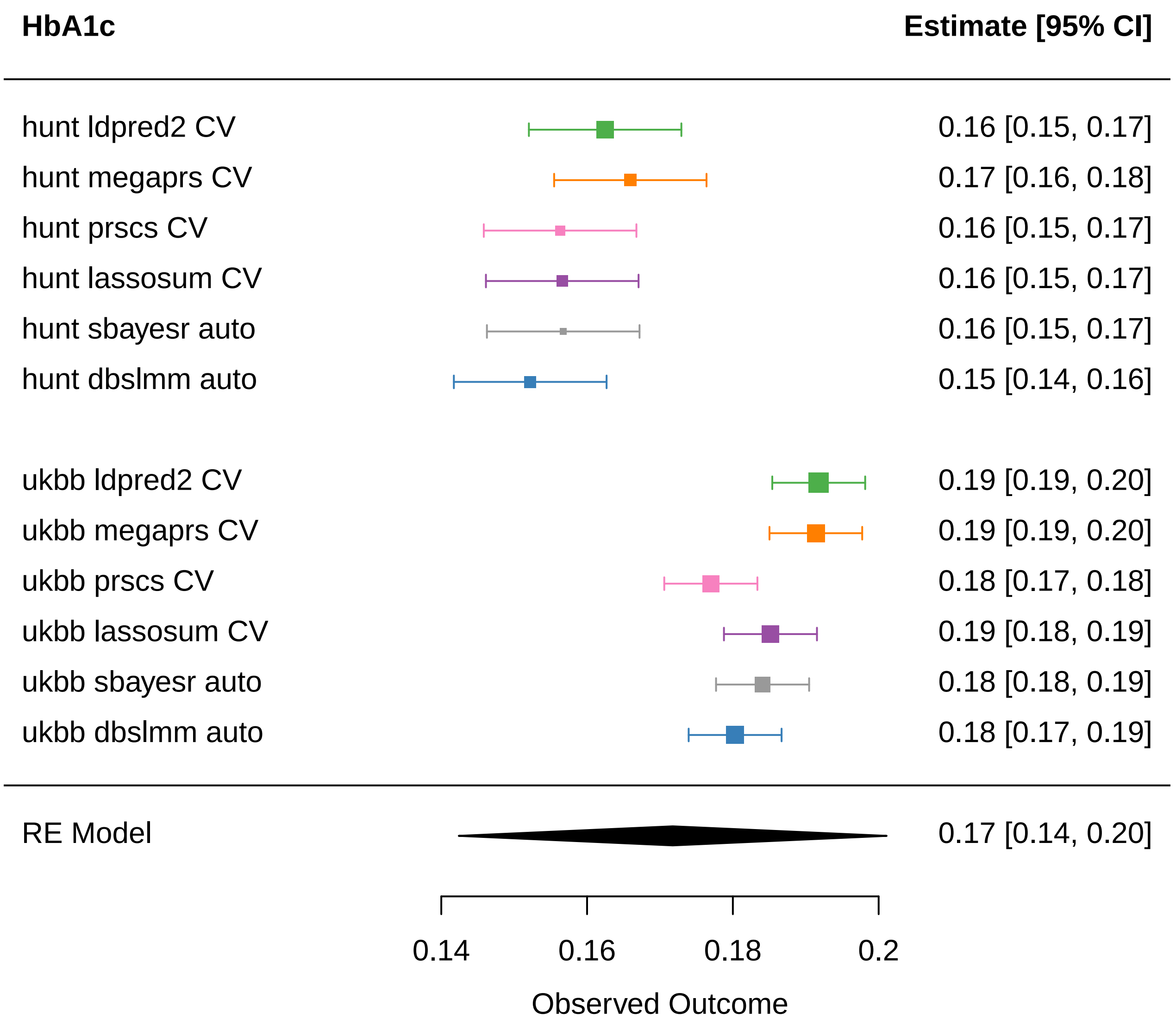
HbA1c
Estimate [95% CI]
hunt ldpred2 CV
0.16 [0.15, 0.17]
hunt megaprs CV
0.17 [0.16, 0.18]
hunt prscs CV
0.16 [0.15, 0.17]
hunt lassosum CV
0.16 [0.15, 0.17]
hunt sbayesr auto
0.16 [0.15, 0.17]
hunt dbslmm auto
0.15 [0.14, 0.16]
ukbb ldpred2 CV
0.19 [0.19, 0.20]
ukbb megaprs CV
0.19 [0.19, 0.20]
ukbb prscs CV
0.18 [0.17, 0.18]
ukbb lassosum CV
0.19 [0.18, 0.19]
ukbb sbayesr auto
0.18 [0.18, 0.19]
ukbb dbslmm auto
0.18 [0.17, 0.19]
RE Model
0.17 [0.14, 0.20]
0.14
0.16
0.18
0.2
Observed Outcome

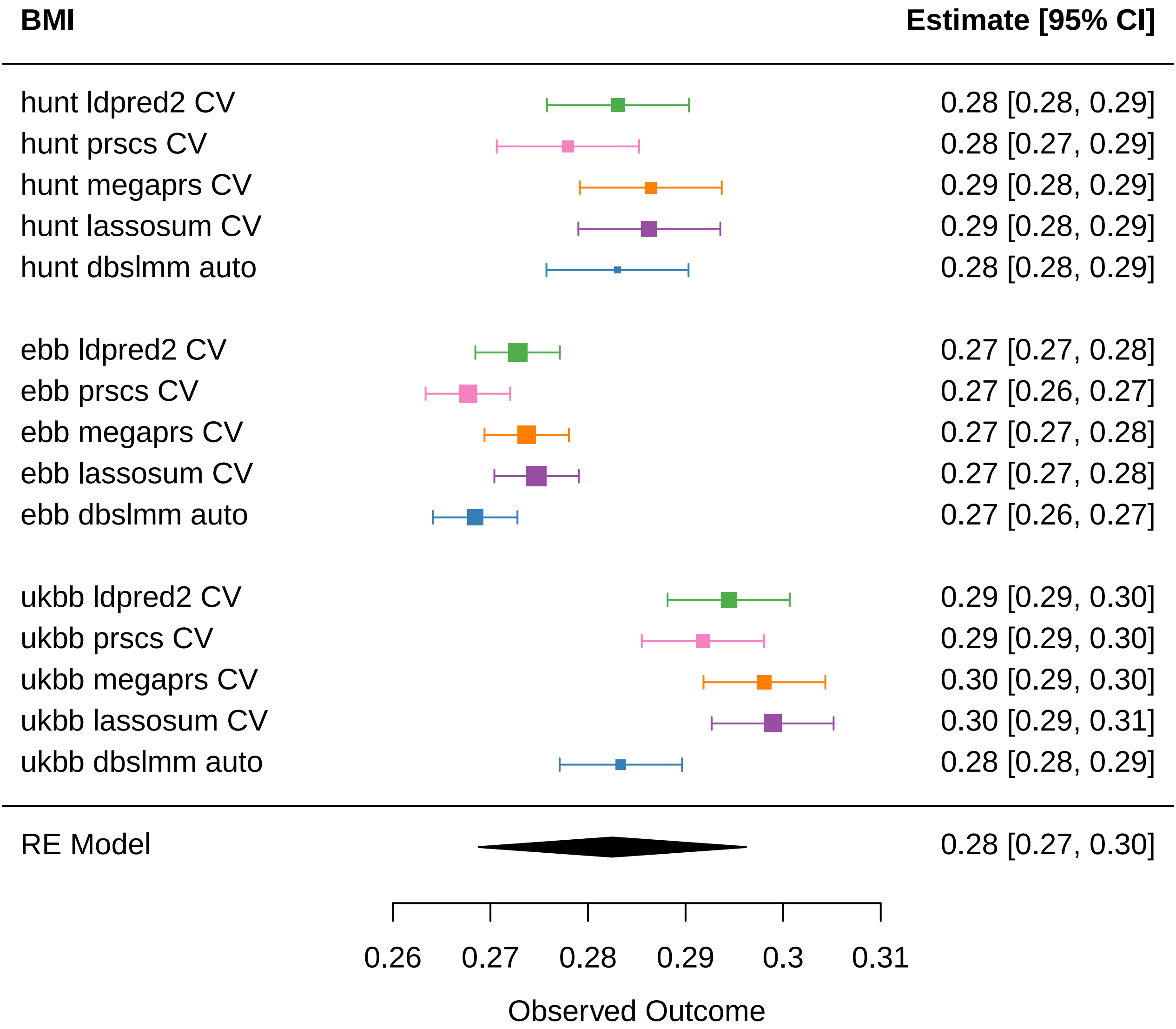
BMI
Estimate [95% CI]
hunt ldpred2 CV
0.28 [0.28, 0.29]
hunt prscs CV
0.28 [0.27, 0.29]
hunt megaprs CV
0.29 [0.28, 0.29]
hunt lassosum CV
0.29 [0.28, 0.29]
hunt dbslmm auto
0.28 [0.28, 0.29]
ebb ldpred2 CV
0.27 [0.27, 0.28]
ebb prscs CV
0.27 [0.26, 0.27]
ebb megaprs CV
0.27 [0.27, 0.28]
ebb lassosum CV
0.27 [0.27, 0.28]
ebb dbslmm auto
0.27 [0.26, 0.27]
ukbb ldpred2 CV
0.29 [0.29, 0.30]
ukbb prscs CV
0.29 [0.29, 0.30]
ukbb megaprs CV
0.30 [0.29, 0.30]
ukbb lassosum CV
0.30 [0.29, 0.31]
ukbb dbslmm auto
0.28 [0.28, 0.29]
RE Model
0.28 [0.27, 0.30]
0.26
0.27
0.28
0.29
0.3
0.31
Observed Outcome

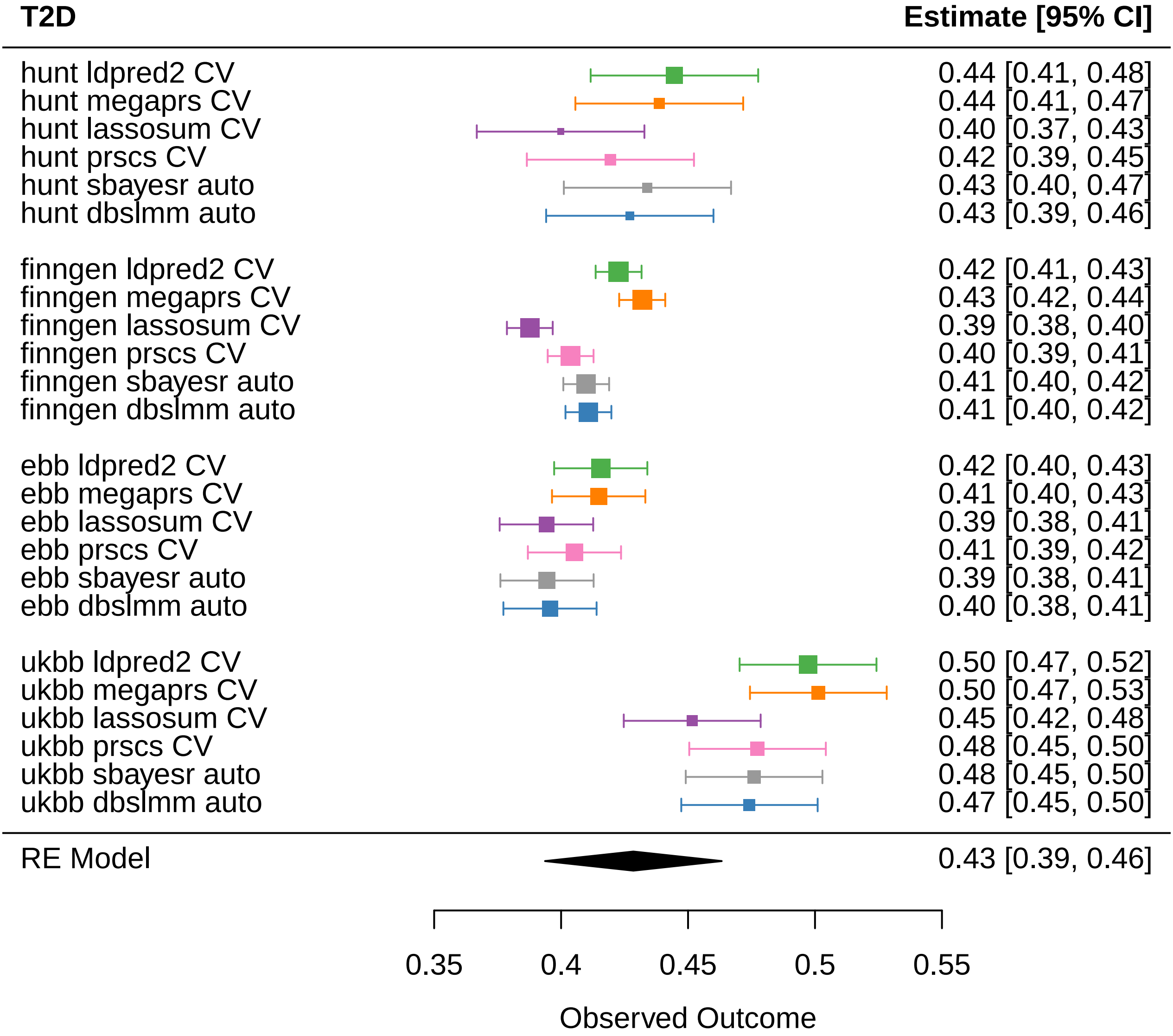
T2D
Estimate [95% CI]
hunt ldpred2 CV 0.44 [0.41, 0.48]
hunt megaprs CV 0.44 [0.41, 0.47]
hunt lassosum CV 0.40 [0.37, 0.43]
hunt prscs CV 0.42 [0.39, 0.45]
hunt sbayesr auto 0.43 [0.40, 0.47]
hunt dbslmm auto 0.43 [0.39, 0.46]
finngen ldpred2 CV 0.42 [0.41, 0.43]
finngen megaprs CV 0.43 [0.42, 0.44]
finngen lassosum CV 0.39 [0.38, 0.40]
finngen prscs CV 0.40 [0.39, 0.41]
finngen sbayesr auto 0.41 [0.40, 0.42]
finngen dbslmm auto 0.41 [0.40, 0.42]
ebb ldpred2 CV 0.42 [0.40, 0.43]
ebb megaprs CV 0.41 [0.40, 0.43]
ebb lassosum CV 0.39 [0.38, 0.41]
ebb prscs CV 0.41 [0.39, 0.42]
ebb sbayesr auto 0.39 [0.38, 0.41]
ebb dbslmm auto 0.40 [0.38, 0.41]
ukbb ldpred2 CV 0.50 [0.47, 0.52]
ukbb megaprs CV 0.50 [0.47, 0.53]
ukbb lassosum CV 0.45 [0.42, 0.48]
ukbb prscs CV 0.48 [0.45, 0.50]
ukbb sbayesr auto 0.48 [0.45, 0.50]
ukbb dbslmm auto 0.47 [0.45, 0.50]
RE Model 0.43 [0.39, 0.46]
0.35
0.4
0.45
0.5
0.55
Observed Outcome

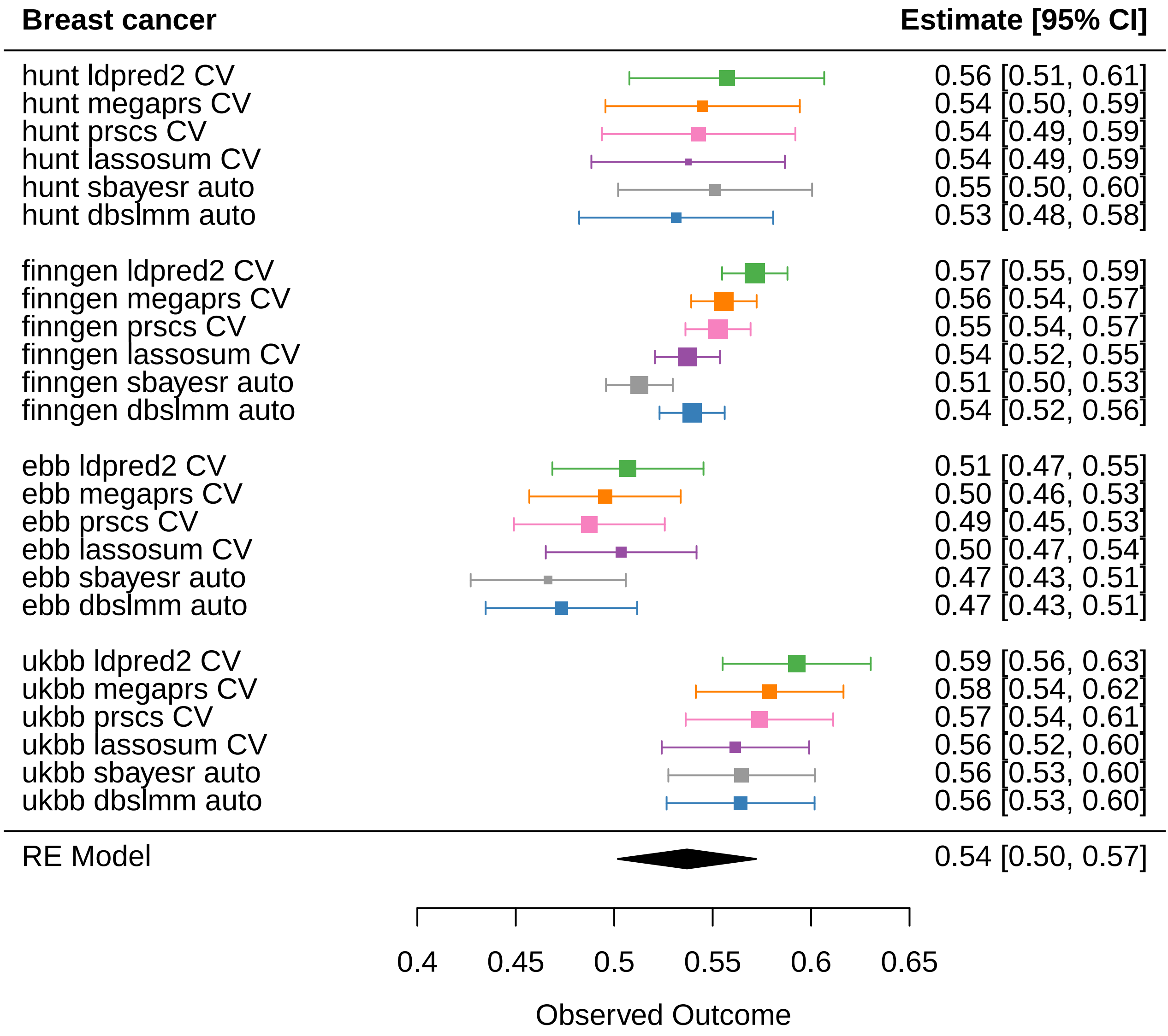
Breast cancer
Estimate [95% CI]
hunt ldpred2 CV 0.56 [0.51, 0.61]
hunt megaprs CV 0.54 [0.50, 0.59]
hunt prscs CV 0.54 [0.49, 0.59]
hunt lassosum CV 0.54 [0.49, 0.59]
hunt sbayesr auto 0.55 [0.50, 0.60]
hunt dbslmm auto 0.53 [0.48, 0.58]
finngen ldpred2 CV 0.57 [0.55, 0.59]
finngen megaprs CV 0.56 [0.54, 0.57]
finngen prscs CV 0.55 [0.54, 0.57]
finngen lassosum CV 0.54 [0.52, 0.55]
finngen sbayesr auto 0.51 [0.50, 0.53]
finngen dbslmm auto 0.54 [0.52, 0.56]
ebb ldpred2 CV 0.51 [0.47, 0.55]
ebb megaprs CV 0.50 [0.46, 0.53]
ebb prscs CV 0.49 [0.45, 0.53]
ebb lassosum CV 0.50 [0.47, 0.54]
ebb sbayesr auto 0.47 [0.43, 0.51]
ebb dbslmm auto 0.47 [0.43, 0.51]
ukbb ldpred2 CV 0.59 [0.56, 0.63]
ukbb megaprs CV 0.58 [0.54, 0.62]
ukbb prscs CV 0.57 [0.54, 0.61]
ukbb lassosum CV 0.56 [0.52, 0.60]
ukbb sbayesr auto 0.56 [0.53, 0.60]
ukbb dbslmm auto 0.56 [0.53, 0.60]
RE Model 0.54 [0.50, 0.57]
0.4
0.45
0.5
0.55
0.6
0.65
Observed Outcome

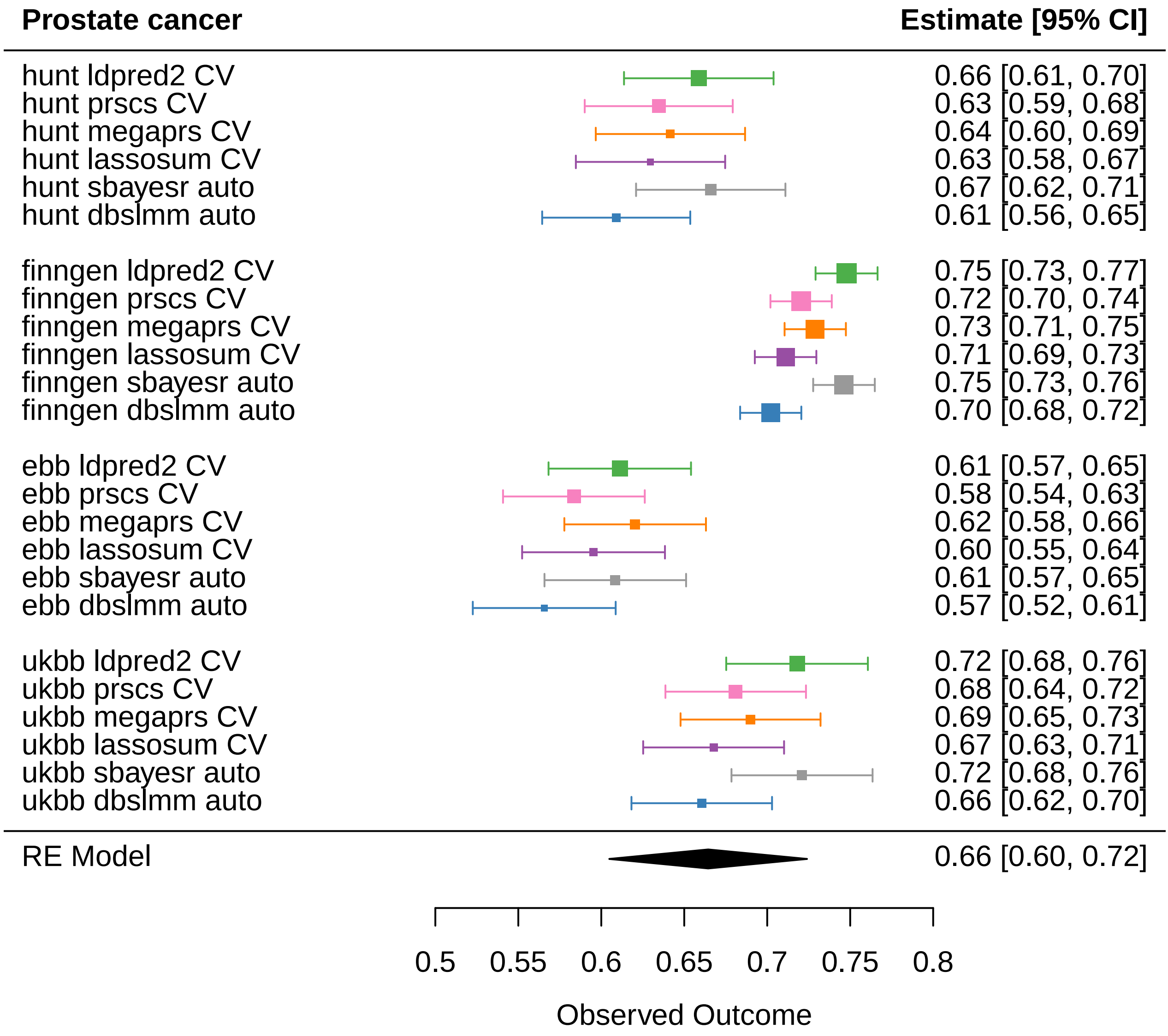

Prostate cancer
Estimate [95% CI]
hunt ldpred2 CV 0.66 [0.61, 0.70]
hunt prscs CV 0.63 [0.59, 0.68]
hunt megaprs CV 0.64 [0.60, 0.69]
hunt lassosum CV 0.63 [0.58, 0.67]
hunt sbayesr auto 0.67 [0.62, 0.71]
hunt dbslmm auto 0.61 [0.56, 0.65]
finngen ldpred2 CV 0.75 [0.73, 0.77]
finngen prscs CV 0.72 [0.70, 0.74]
finngen megaprs CV 0.73 [0.71, 0.75]
finngen lassosum CV 0.71 [0.69, 0.73]
finngen sbayesr auto 0.75 [0.73, 0.76]
finngen dbslmm auto 0.70 [0.68, 0.72]
ebb ldpred2 CV 0.61 [0.57, 0.65]
ebb prscs CV 0.58 [0.54, 0.63]
ebb megaprs CV 0.62 [0.58, 0.66]
ebb lassosum CV 0.60 [0.55, 0.64]
ebb sbayesr auto 0.61 [0.57, 0.65]
ebb dbslmm auto 0.57 [0.52, 0.61]
ukbb ldpred2 CV 0.72 [0.68, 0.76]
ukbb prscs CV 0.68 [0.64, 0.72]
ukbb megaprs CV 0.69 [0.65, 0.73]
ukbb lassosum CV 0.67 [0.63, 0.71]
ukbb sbayesr auto 0.72 [0.68, 0.76]
ukbb dbslmm auto 0.66 [0.62, 0.70]
RE Model 0.66 [0.60, 0.72]
0.5
0.55
0.6
0.65
0.7
0.75
0.8
Observed Outcome

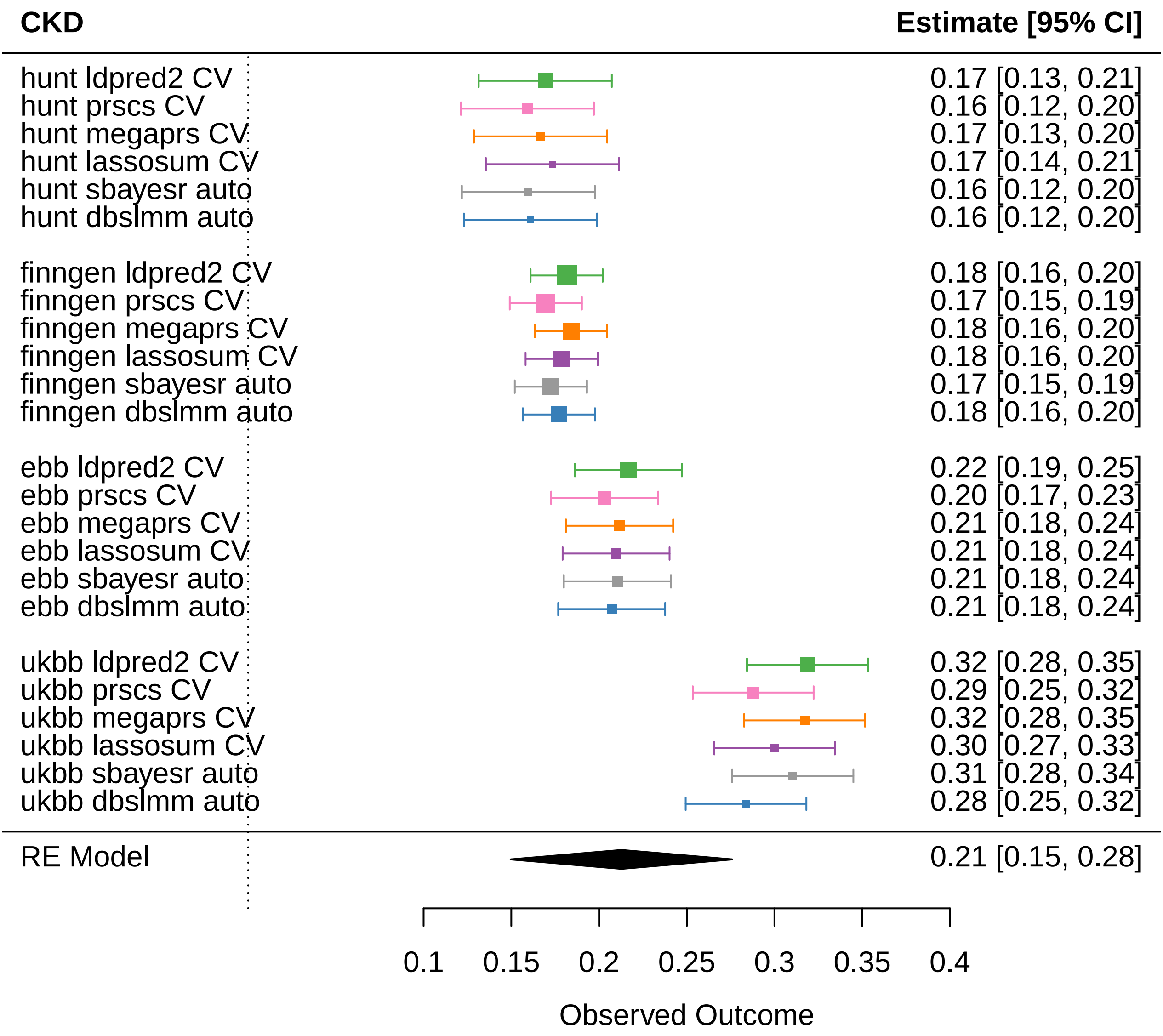
CKD
Estimate [95% CI]
hunt ldpred2 CV 0.17 [0.13, 0.21]
hunt prscs CV 0.16 [0.12, 0.20]
hunt megaprs CV 0.17 [0.13, 0.20]
hunt lassosum CV 0.17 [0.14, 0.21]
hunt sbayesr auto 0.16 [0.12, 0.20]
hunt dbslmm auto 0.16 [0.12, 0.20]
finngen ldpred2 CV 0.18 [0.16, 0.20]
finngen prscs CV 0.17 [0.15, 0.19]
finngen megaprs CV 0.18 [0.16, 0.20]
finngen lassosum CV 0.18 [0.16, 0.20]
finngen sbayesr auto 0.17 [0.15, 0.19]
finngen dbslmm auto 0.18 [0.16, 0.20]
ebb ldpred2 CV 0.22 [0.19, 0.25]
ebb prscs CV 0.20 [0.17, 0.23]
ebb megaprs CV 0.21 [0.18, 0.24]
ebb lassosum CV 0.21 [0.18, 0.24]
ebb sbayesr auto 0.21 [0.18, 0.24]
ebb dbslmm auto 0.21 [0.18, 0.24]
ukbb ldpred2 CV 0.32 [0.28, 0.35]
ukbb prscs CV 0.29 [0.25, 0.32]
ukbb megaprs CV 0.32 [0.28, 0.35]
ukbb lassosum CV 0.30 [0.27, 0.33]
ukbb sbayesr auto 0.31 [0.28, 0.34]
ukbb dbslmm auto 0.28 [0.25, 0.32]
RE Model 0.21 [0.15, 0.28]
0.1
0.15
0.2
0.25
0.3
0.35
0.4
Observed Outcome

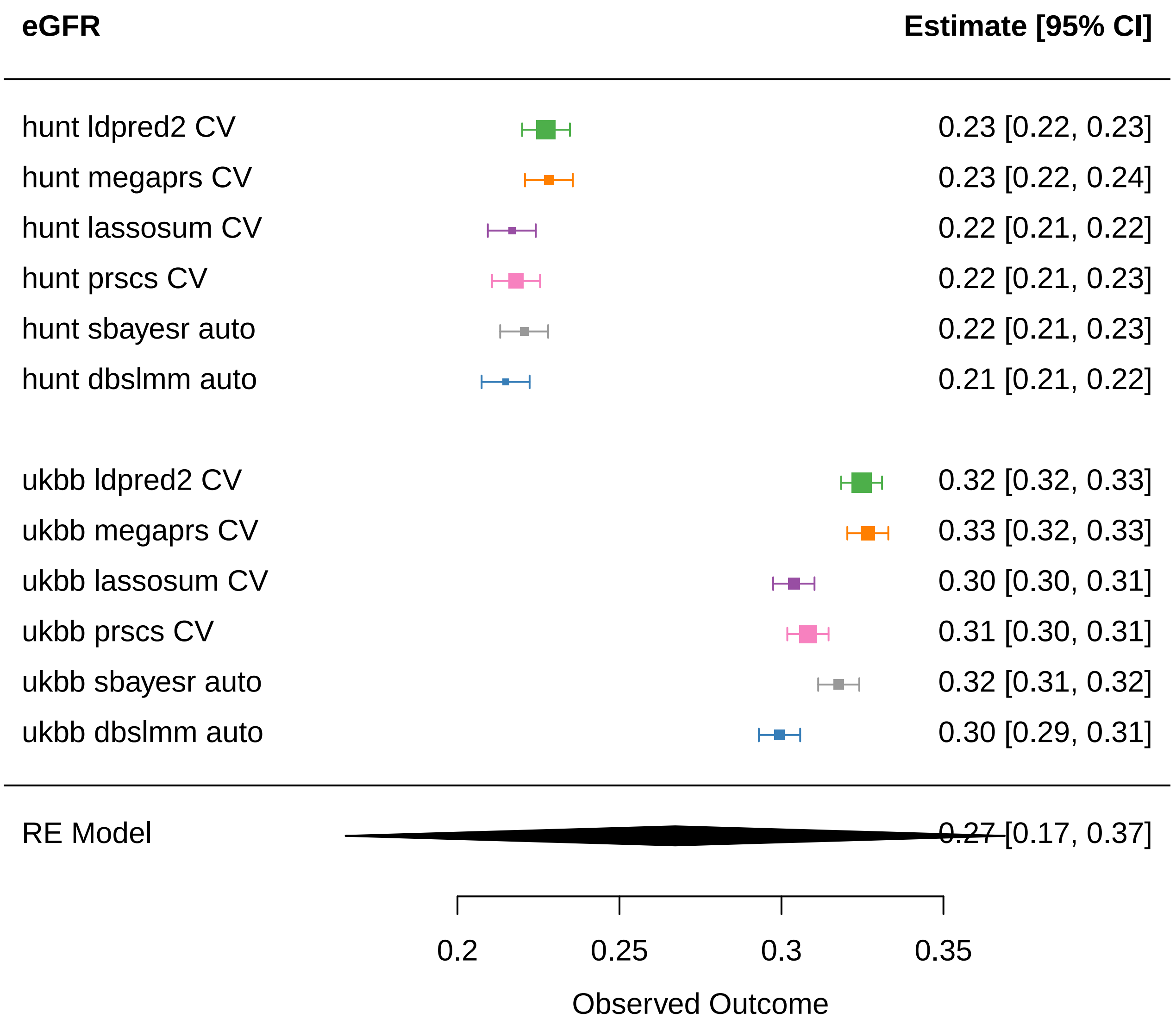

eGFR
Estimate [95% CI]
hunt ldpred2 CV 0.23 [0.22, 0.23]
hunt megaprs CV 0.23 [0.22, 0.24]
hunt lassosum CV 0.22 [0.21, 0.22]
hunt prscs CV 0.22 [0.21, 0.23]
hunt sbayesr auto 0.22 [0.21, 0.23]
hunt dbslmm auto 0.21 [0.21, 0.22]
ukbb ldpred2 CV 0.32 [0.32, 0.33]
ukbb megaprs CV 0.33 [0.32, 0.33]
ukbb lassosum CV 0.30 [0.30, 0.31]
ukbb prscs CV 0.31 [0.30, 0.31]
ukbb sbayesr auto 0.32 [0.31, 0.32]
ukbb dbslmm auto 0.30 [0.29, 0.31]
RE Model 0.27 [0.17, 0.37]
0.2
0.25
0.3
0.35
Observed Outcome

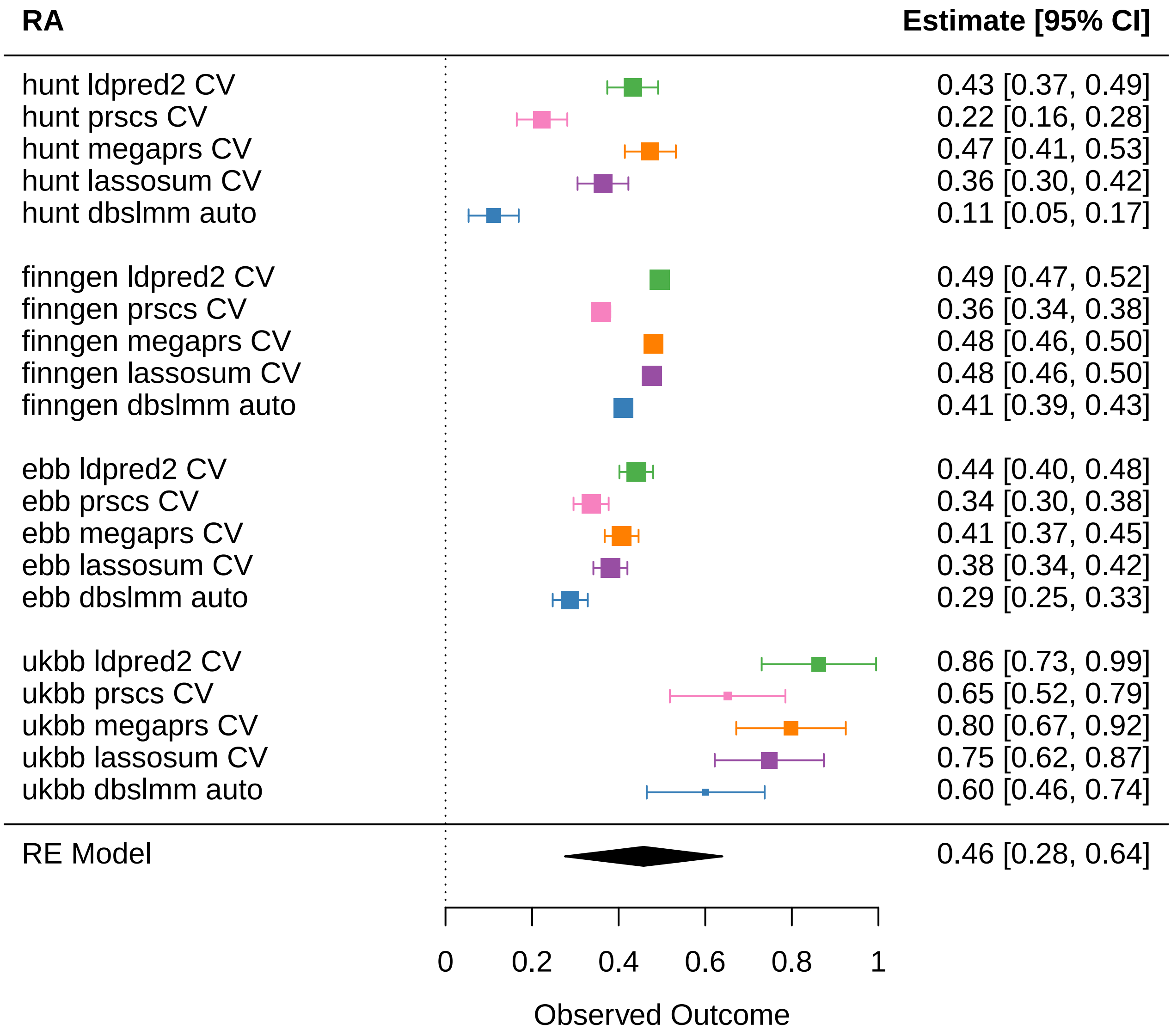
RA
Estimate [95% CI]
hunt ldpred2 CV
hunt prscs CV
hunt megaprs CV
hunt lassosum CV
hunt dbslmm auto
finngen ldpred2 CV
finngen prscs CV
finngen megaprs CV
finngen lassosum CV
finngen dbslmm auto
ebb ldpred2 CV
ebb prscs CV
ebb megaprs CV
ebb lassosum CV
ebb dbslmm auto
ukbb ldpred2 CV
ukbb prscs CV
ukbb megaprs CV
ukbb lassosum CV
ukbb dbslmm auto
RE Model
0.43 [0.37, 0.49]
0.22 [0.16, 0.28]
0.47 [0.41, 0.53]
0.36 [0.30, 0.42]
0.11 [0.05, 0.17]
0.49 [0.47, 0.52]
0.36 [0.34, 0.38]
0.48 [0.46, 0.50]
0.48 [0.46, 0.50]
0.41 [0.39, 0.43]
0.44 [0.40, 0.48]
0.34 [0.30, 0.38]
0.41 [0.37, 0.45]
0.38 [0.34, 0.42]
0.29 [0.25, 0.33]
0.86 [0.73, 0.99]
0.65 [0.52, 0.79]
0.80 [0.67, 0.92]
0.75 [0.62, 0.87]
0.60 [0.46, 0.74]
0.46 [0.28, 0.64]
0
0.2
0.4
0.6
0.8
1
Observed Outcome

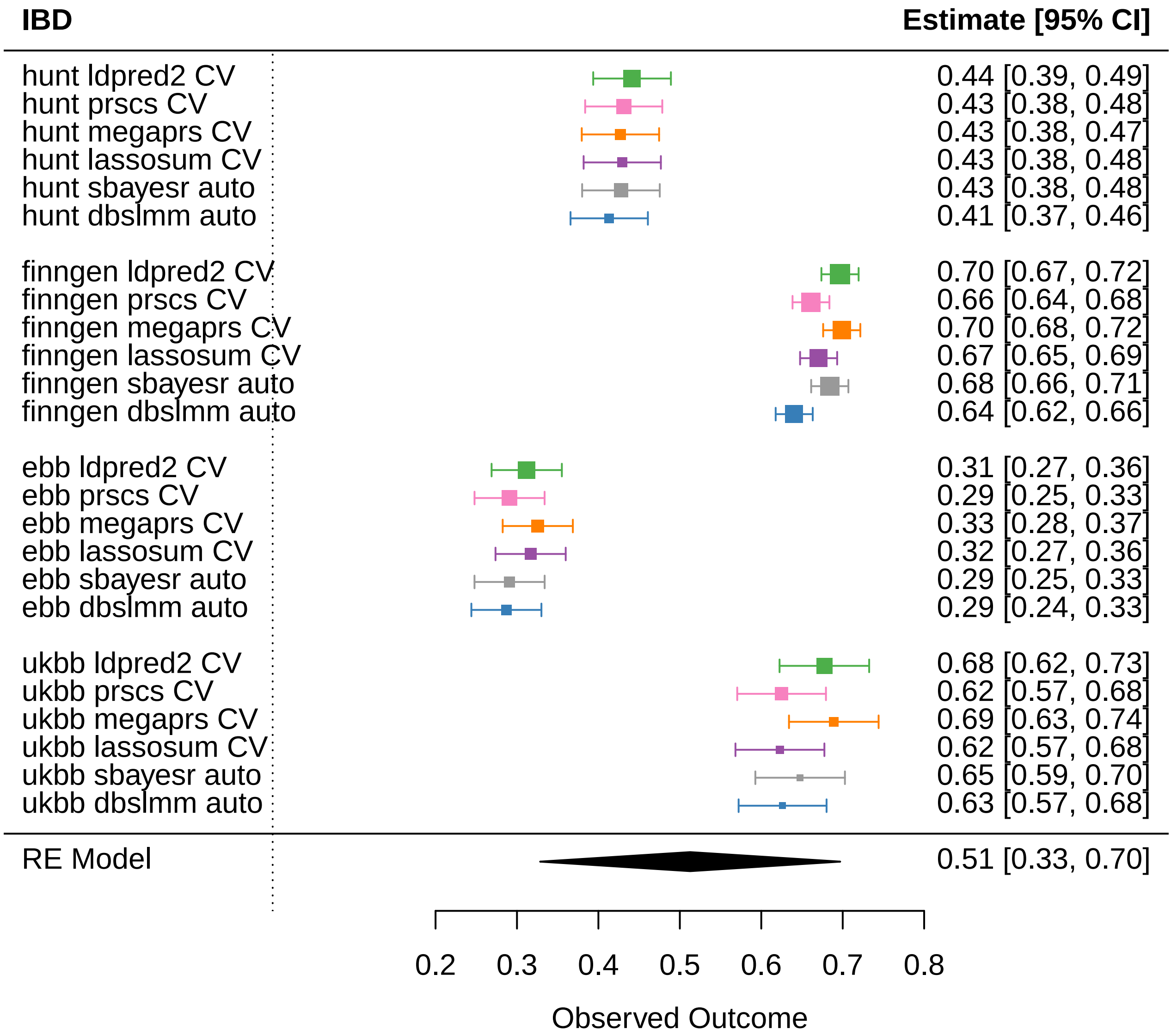
IBD
Estimate [95% CI]
hunt ldpred2 CV 0.44 [0.39, 0.49]
hunt prscs CV 0.43 [0.38, 0.48]
hunt megaprs CV 0.43 [0.38, 0.47]
hunt lassosum CV 0.43 [0.38, 0.48]
hunt sbayesr auto 0.43 [0.38, 0.48]
hunt dbslmm auto 0.41 [0.37, 0.46]
finngen ldpred2 CV 0.70 [0.67, 0.72]
finngen prscs CV 0.66 [0.64, 0.68]
finngen megaprs CV 0.70 [0.68, 0.72]
finngen lassosum CV 0.67 [0.65, 0.69]
finngen sbayesr auto 0.68 [0.66, 0.71]
finngen dbslmm auto 0.64 [0.62, 0.66]
ebb ldpred2 CV 0.31 [0.27, 0.36]
ebb prscs CV 0.29 [0.25, 0.33]
ebb megaprs CV 0.33 [0.28, 0.37]
ebb lassosum CV 0.32 [0.27, 0.36]
ebb sbayesr auto 0.29 [0.25, 0.33]
ebb dbslmm auto 0.29 [0.24, 0.33]
ukbb ldpred2 CV 0.68 [0.62, 0.73]
ukbb prscs CV 0.62 [0.57, 0.68]
ukbb megaprs CV 0.69 [0.63, 0.74]
ukbb lassosum CV 0.62 [0.57, 0.68]
ukbb sbayesr auto 0.65 [0.59, 0.70]
ukbb dbslmm auto 0.63 [0.57, 0.68]
RE Model 0.51 [0.33, 0.70]
0.2 0.3 0.4 0.5 0.6 0.7 0.8
Observed Outcome

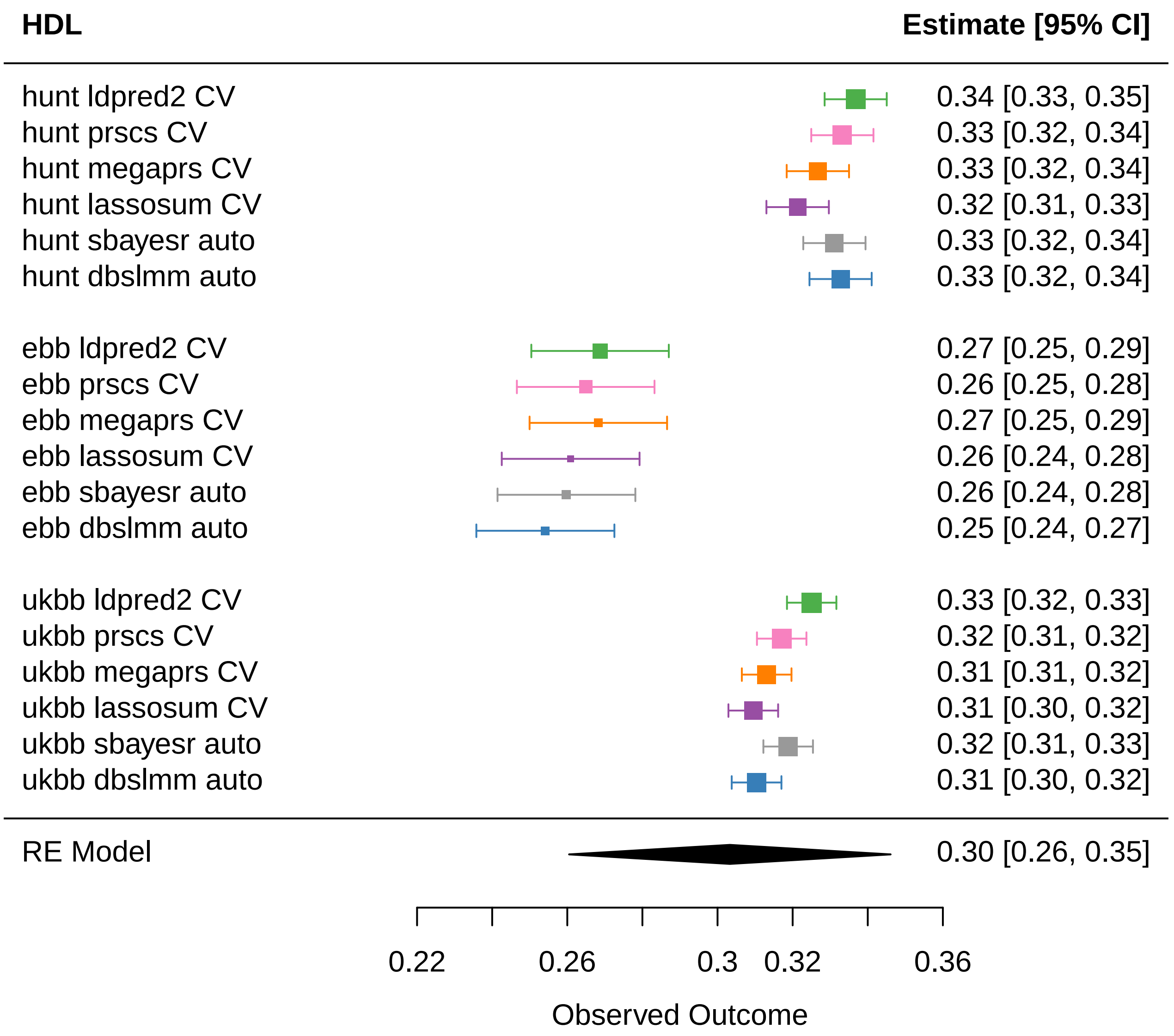

HDL
Estimate [95% CI]
hunt ldpred2 CV
hunt prscs CV
hunt megaprs CV
hunt lassosum CV
hunt sbayesr auto
hunt dbslmm auto
ebb ldpred2 CV
ebb prscs CV
ebb megaprs CV
ebb lassosum CV
ebb sbayesr auto
ebb dbslmm auto
ukbb ldpred2 CV
ukbb prscs CV
ukbb megaprs CV
ukbb lassosum CV
ukbb sbayesr auto
ukbb dbslmm auto
RE Model
0.34 [0.33, 0.35]
0.33 [0.32, 0.34]
0.33 [0.32, 0.34]
0.32 [0.31, 0.33]
0.33 [0.32, 0.34]
0.33 [0.32, 0.34]
0.27 [0.25, 0.29]
0.26 [0.25, 0.28]
0.27 [0.25, 0.29]
0.26 [0.24, 0.28]
0.26 [0.24, 0.28]
0.25 [0.24, 0.27]
0.33 [0.32, 0.33]
0.32 [0.31, 0.32]
0.31 [0.31, 0.32]
0.31 [0.30, 0.32]
0.32 [0.31, 0.33]
0.31 [0.30, 0.32]
0.30 [0.26, 0.35]
0.22
0.26
0.3
0.32
0.36
Observed Outcome
